# Spontaneous fast-ultradian dynamics of polymorphic interictal events in drug-resistant focal epilepsy

**DOI:** 10.1101/2023.04.05.23288085

**Authors:** Damián Dellavale, Francesca Bonini, Francesca Pizzo, Julia Makhalova, Fabrice Wendling, Jean-Michel Badier, Fabrice Bartolomei, Christian-George Bénar

## Abstract

**Objective:** We studied the rate dynamics of interictal events occurring over fast-ultradian time scales, as commonly examined in clinics to guide surgical planning in epilepsy.

**Methods:** Stereoelectroencephalography (SEEG) traces of 35 patients with good surgical outcome (Engel I) were analyzed. For this, we developed a general data mining method aimed at clustering the plethora of transient waveform shapes including interictal epileptiform discharges (IEDs), and assessed the temporal fluctuations in the capability to map the epileptogenic zone (EZ) of each type of event.

**Results:** We found that the fast-ultradian dynamics of the IEDs rate may effectively impair the precision of EZ identification, and appear to occur spontaneously, that is, not triggered by or exclusively associated with a particular cognitive task, wakefulness, sleep, seizure occurrence, post-ictal state or antiepileptic drug withdrawal. Propagation of IEDs from the EZ to the propagation zone (PZ) could explain the observed fast-ultradian fluctuations in a reduced fraction of the analyzed patients, suggesting that other factors like the excitability of the epileptogenic tissue could play a more relevant role. A novel link was found between the fast-ultradian dynamics of the overall rate of polymorphic events and the rate of specific IEDs subtypes. We exploited this feature to estimate in each patient the 5 min interictal epoch for near-optimal EZ and resected zone (RZ) localization, which resulted at the population level better than those obtained using either 1) the whole time series available in each patient (P = 0.084 for EZ, P < 0.001 for RZ, Wilcoxon signed rank test) and 2) 5 min epochs randomly sampled from the interictal recordings of each patient (P < 0.05 for EZ, P < 0.001 for RZ, 10^5^ random samplings).

*Significance:* Our results highlight the relevance of the fast-ultradian IEDs dynamics in mapping the EZ, and show how it can be prospectively estimated to inform surgical planning in epilepsy.

*Key Points:*

- The rate of IED observed in SEEG recordings undergoes spontaneous fluctuations over fast-ultradian time scales.
- The fast-ultradian dynamics of the IED rate may impair the EZ identification and hence are clinically relevant for surgical planning.
- Propagation does not fully explain the fast-ultradian dynamics of the IED rate constraining the precision to localize the EZ.
- Interictal epochs, as commonly examined in clinics, producing near-optimal EZ mapping can be inferred based solely on the LFP dynamics.
- Fluctuations of the AUPREC based on epileptic and non-epileptic events are linked to scale-free and scale-rich processes, respectively.

## Introduction

The success rate of the surgical treatment for drug-resistant focal epilepsies is in the order of 60%.^1,2^ This imperfect outcome can be partially explained by the fact that optimal delineation of the epileptogenic zone (EZ) remains unknown. In this context, significant efforts are being made to quantify intracranial EEG^3^ and extract biomarkers that can best predict the location of the EZ. Epileptic spikes are the classic interictal biomarker of epilepsy, used by epileptologists as part of standard practice to inform surgical planing.^4^ The relationship of the EZ with the epileptiform spikes is complex. In a previous study it was shown a variable correlation of the spike rate with EZ, but particularly important in focal cortical dysplasia.^5^ Currently, there is no gold standard to define subtypes of epileptiform discharges and to differentiate them from non-epileptic paroxysmal events.^6^ However, some characteristics of spikes have been found to be better markers of EZ, such as frequency of occurrence, association with high frequency oscillations (HFO)^7,8^ or with gamma activity.^9^ An underestimated dimension is the time-varying factors underlying the genesis of different subtypes of interictal epileptiform discharges.^10^ In this regard, there are important open issues related to the way IEDs fluctuate in spatial extent,^11^ waveform shape (see Figure 2A in Tomlinson et al.^12^) and rate of occurrence. The temporal dynamics of IEDs expand over a wide range of time scales, from milliseconds associated with the propagation across macroscopic networks^12,13^ to circadian (day) and even multidien (week/month) fluctuations in relation to sleep and ictogenesis among other factors.^14-18^ Therefore, the integration of all available information over a broad range of spatial and temporal scales is crucial to improve the EZ localization based on interictal SEEG recordings.

In this work we report on a quantitative analysis of the fluctuations of the IEDs rate over fast-ultradian time scales, as well as the impact of such temporal fluctuations on the capacity of IEDs to map the EZ. The fast-ultradian dynamics analyzed in our work refers to temporal fluctuations of the rate of interictal events, not necessarily periodic, expanding over time scales ranging from sub-minute up to half an hour (i.e. sub-hour temporal dynamics).^19^ In particular, we focus on the study of previously unexplored IEDs rate dynamics of spontaneous nature and occurring in SEEG recordings as commonly examined in clinics to inform surgical planning in epilepsy (in the range of 5 minutes to 2 hours of SEEG recordings). We developed a general data mining method aimed at clustering the plethora of transient waveform shapes emerging in interictal SEEG recordings (referred in this study as polymorphic interictal events), and assessed the fluctuation in predictive power of each type of event. Importantly, the proposed method for polymorphic events analysis paves the way to unveil a novel and counter-intuitive link between the dynamics of the overall rate of polymorphic events and the rate of specific subtypes of epileptiform spikes which is exploited here to improve the EZ localization.

## Methods

### Patients and intracerebral recordings

Patients with drug resistant focal epilepsy were selected from the database of the Epileptology Unit of La Timone Hospital based on the outcome of the resective surgery. The final study includes 35 patients (20 women and 15 men) with Engel I seizure outcome classification at least 12 months after the surgical procedure, identified in the period 2008 to 2019. A variety of pathologies and electrode coverages were represented (see Table 1). The median age at the time of the SEEG surgery was 27 years (range = 19.5 - 41 years). Written informed consent regarding the SEEG procedure was obtained from all individual participants included in the study. In all patients, indication for SEEG exploration was based on Phase I presurgical non-invasive assessments for pharmacoresistant focal epilepsy including examination of detailed clinical history, neurological evaluation, neuropsychological testing, long-term scalp video-EEG monitoring and high-resolution structural magnetic resonance imaging (MRI). SEEG exploration was performed using intracerebral electrodes (Dixi Medical or Alcis Neuro (France); 10 - 15 contacts, length: 2 mm, diameter: 0.8 mm, 1.5 mm apart), placed intracranially according to Talairach stereotactic method^20^. The anatomical targets for electrode placement were defined based on the hypotheses about EZ localization resulting from Phase I. A postoperative computed tomography (CT) scan and/or MRI was performed to verify the spatial accuracy of the implantation. CT/MRI data coregistration was performed to check the anatomical location of each contact along the electrode trajectory. Local field potentials (LFP) from the SEEG electrodes were recorded on a 128-channel system (Natus/Deltamed) sampled at 512 Hz or 1024 Hz (16 bits resolution), with a built-in hardware high-pass filter (cutoff frequency = 0.16 Hz) and an antialiasing low-pass filter (cutoff frequency = 170 Hz for 512 Hz sampling rate, or 340 Hz for 1024 Hz sampling rate). Video-SEEG recordings were performed as long as necessary (1 - 3 weeks) to record several of the patient’s habitual seizures. Long term video-SEEG recordings following withdrawal of antiepileptic drugs were judged to be necessary to delineate the localization of the epileptogenic zone (EZ) for surgical treatment. The SEEG recordings analyzed in this work were chosen at least 2 days after the electrode implantation surgical procedure, and when possible before medication tapering, to limit possible effects of general anesthesia and antiepileptic drug withdrawal. In addition, we selected interictal recordings that were temporally distant from the preceding and the following seizure by at least 2 hs. The SEEG traces were recorded during time intervals in which the patients were either awake at rest or during non-REM sleep. For the awake state at rest, the patients were instructed to remain awake and during the SEEG recordings they were laying in bed doing nothing (not engaging in any particular cognitive task). Some patients, in particular the ones corresponding to long SEEG recordings (> 30 min), could have undergo drowsy (not sleep) state during part of the recording time interval. In all the cases, the patient state (awake at rest or non-REM sleep) was confirmed by the dedicated staff who reviewed the SEEG traces and the video monitoring the patient during the intracerebral recordings. Unless otherwise indicated, the results presented in this work correspond to the 35 patients listed in Table 1 and S1 for the awake state at rest. The results corresponding to the non-REM sleep are presented and discussed in Appendix S1, Section 11. In order to compare the results between two SEEG recording sessions taken at different days and time of day for the same patient state (awake at rest), a second group of 12 patients are presented and discussed in Appendix S1, Section 12. Not all the patients included in this second group have Engel I seizure outcome (see Table S2). The epileptologists team performed a visual evaluation of the SEEG traces before the analysis to identify interictal periods with a limited amount of artifacts (see Table S1). The median time-length of the interictal SEEG traces among the 35 patients in awake state was 28.2 min, range = 23.5 - 31.6 min (see Table S1). The classification of the SEEG contacts in EZ, propagation zone (PZ) and non-involved zone (NIZ) was made according to the sites of seizure initiation (SOZ) based on visual analysis and on the Epileptogenicity Index (EI).^4,5,21^ Further details about the classification of the SEEG contacts can be found in the Appendix S1, Section 1. Across the 35 patients included in the Table 1, a total of 428 brain regions were analyzed (median per patient = 13, range = 11 - 14). There were a total of 5154 SEEG contacts, 962 in epileptogenic zone (EZ), 904 in propagation zone (PZ), 3288 in non-involved zone (NIZ) and 1486 in the resected zone (RZ). The SEEG macroelectrode contacts were converted to a bipolar referencing montage for subsequent analysis. Bipolar channels were obtained as the difference between signals recorded from spatially adjacent contacts pertaining to the same depth electrode array. For other clinical characteristics of the patients see Table 1.

**Table 1:** Patients clinical information. All the 35 patients (20 women and 15 men) have Engel I seizure outcome classification at least 12 months after the surgical procedure. The information regarding the resected zone was not available for the patient 13. The time of day (or night) when the SEEG recordings were made and the length of the interictal SEEG traces corresponding to these patients can be found in the Table S1 of the Appendix S1. To ensure the anonymity, serial numbers attributed randomly to each patient are used as patients ID. The resulting ID numbers have no correlation with any clinical information of the patients.

| Patient ID | Age at SEEG | Sex | EZ localization | Etiology |
| --- | --- | --- | --- | --- |
| 1 | 36-40 | M | Temp - Fr | FCD |
| 2 | 26-30 | F | Temp | GG |
| 3 | 41-45 | F | Temp | HS |
| 4 | 16-20 | M | Fr | G |
| 5 | 21-25 | F | SMA | FCD |
| 6 | 26-30 | M | Temp | HS |
| 7 | 26-30 | F | Temp | HS |
| 8 | 26-30 | F | Par | FCD |
| 9 | 41-45 | M | Ins | N/A |
| 10 | 26-30 | M | Fr | G |
| 11 | 56-60 | M | Temp - Fr | FCD |
| 12 | 41-45 | M | Temp - Fr | G |
| 13 | 41-45 | F | pre Mot | FCD |
| 14 | 11-15 | M | Mot - pre Mot | Stroke |
| 15 | 51-55 | F | right Op and Pos Ins | FCD |
| 16 | 1-5 | M | Occ | GG |
| 17 | 11-15 | F | Fr | TS |
| 18 | 56-60 | M | left Temp Lat and Par | HS |
| 19 | 36-40 | F | right Mes - Temp | HS |
| 20 | 6-10 | F | left Fr | Neonatal ACV |
| 21 | 46-50 | M | right Ant MesTemp and Ins | HS |
| 22 | 11-15 | M | left Temp - Mes | DNET |
| 23 | 11-15 | F | left Temp Basal and<br>left Temp - Occ | Neonatal ACV |
| 24 | 66-70 | M | left Temp - Mes | HS |
| 25 | 11-15 | M | left Hem | N/A |
| 26 | 31-35 | F | left Temp - Mes | G |
| 27 | 6-10 | M | left Temp - Mes | HS |
| 28 | 11-15 | F | right Fr | FCD |
| 29 | 21-25 | F | right Lat Temp | Non-specific |
| 30 | 46-50 | F | left Temp - Mes | Non-specific |
| 31 | 36-40 | F | Temp Lat | GG |
| 32 | 21-25 | F | Temp Lat | FCD |
| 33 | 21-25 | F | Par | DNET |
| 34 | 31-35 | F | Temp - Mes | HS + FCD |
| 35 | 21-25 | F | Fr | FCD |

Symbols and abbreviations: F, Female; M, Male; Temp, Temporal lobe; Fr, Frontal lobe; Par, Parietal lobe; Occ, Occipital lobe; SMA, Supplementary motor area; Ins, Insular; Mot, Motor cortex; Op, Opercular; Ant, Anterior; Pos, Posterior; Lat, Lateral; Mes, Mesial; Hem, Hemisphere; FCD: Focal cortical dysplasia; GG, Ganglioglioma; HS, Hippocampal sclerosis; N/A, Not available; G, Gliosis; TS, Tuberous sclerosis; DNET: Dysembryoplastic neuro-epithelial tumor; PNH, Periventricular nodular heterotopia; NO, Not operated.

### Data and statistical analysis

To systematically study the variety of transient waveform shapes emerging in the interictal SEEG dynamics, we developed the Nested Outlier Detection (NODE) algorithm as a general data mining method. The NODE algorithm uses the Local False Discovery Rate (LFDR) method^22,23^ to define the interictal events by detecting anomalies (i.e. outliers) of amplitude across the frequency bands of interest. Figure 1A schematizes the main processing steps associated with the NODE algorithm. Briefly, the NODE algorithm assigns to each event a 4-digit label. Each label digit represents the proportion of the detected anomalies that can be expected to be true outliers in the frequency band associated with that digit. The four frequency bands range from High-Delta band corresponding to the first label digit from the left, to Ripple band corresponding to the last label digit from the left. For instance, the label 0_09_09_0 groups all the interictal events with a proportion of 90% of them that can be expected to have true outliers in the two medium frequency bands [8 Hz - 32 Hz] and [30 Hz - 155 Hz]. The label 0_09_09_05 groups sharper transient waveform shapes since these paroxysmal events have, in addition to the outliers in the two medium frequency bands, a proportion of 50% of the detected anomalies associated with the Ripple band [150 Hz - 255 Hz] of these events that can be expected to be true outliers (see Figure 1C). The label 09_09_09_05 groups the spike-wave complexes with a proportion of 90% of them that can also be expected to have true outliers in the low frequency band [1 Hz - 10 Hz] corresponding to the first label digit from the left (see Figure 1B). Further details about the NODE algorithm and the semi-supervised constrained clustering method,^24^ can be found in the Appendix S1, Section 2. Time-frequency maps of the polymorphic events were computed as scalograms using Morlet wavelets (see Appendix S1, Section 3).^25^ To quantify the capability of the subtypes of events identified by the NODE algorithm in segregating the SEEG channels involved in the epileptogenic zone (EZ) from those not involved (NIZ), we implemented a precision and recall analysis which is a suitable tool for imbalanced classification problems (see Appendix S1, Section 4). To compare distributions of paired samples we used a two-tailed non-parametric Wilcoxon signed rank test with α = 0.05. A non-parametric permutation test based on random sampling without replacement was used for non-paired group analysis. Unless otherwise indicated, all the reported P values were Bonferroni-adjusted to correct for multiple comparisons. In all the violin plots, center gray boxes represent the 25th and 75th percentiles, whiskers (gray lines) extend to the most extreme data points not considered as outliers (1.5 times the interquartile range (IQR)), star markers represent outliers. The center white circle and white line indicate the median and mean, respectively. For further details about the methods used to characterize the fast-ultradian dynamics associated with the interictal events rate, the reader is referred to the Appendix S1, Sections 5 and 6.

**Figure 1:**
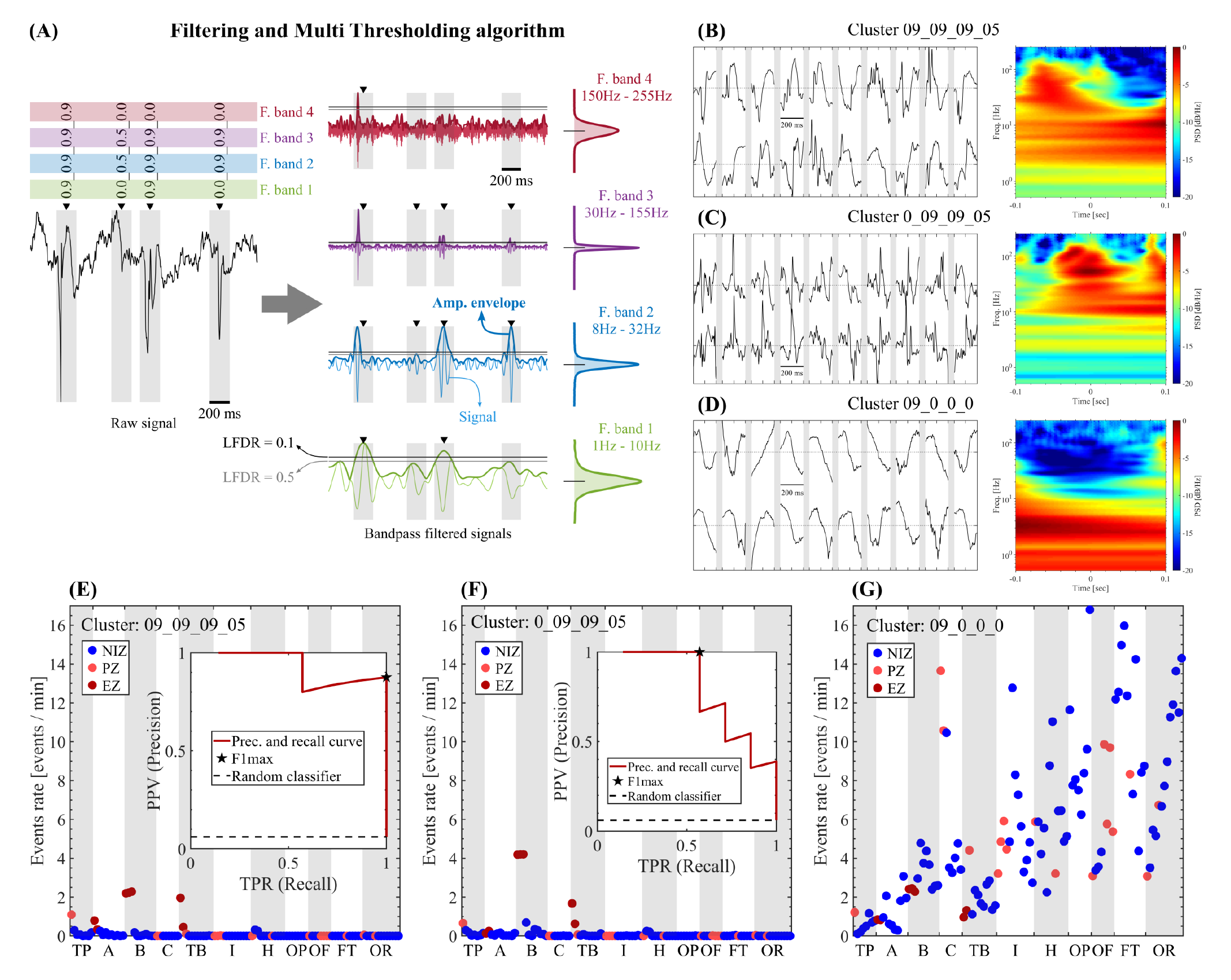
Nested Outlier Detection (NODE) algorithm. **(A)** Schematic representation of the filtering and thresholding stages of the NODE algorithm together with the resulting 4-digit labels assigned to the detected events. **(B, C, D)** Raw time series and whitened time-frequency maps (scalograms using Morlet wavelets) for three clusters of events detected in the EZ (B: Hippocampus cephalis) of the patient 7. **(E, F, G)** Mean rate of events in each bipolar channel obtained from the whole interictal SEEG recording available for the patient 7 (60 min). The insets included in panels and **(F)** show the results of the precision and recall analysis for the IEDs subtypes 09_09_09_05 (F1 max = 0.93) and 0_09_09_05 (F1 max = 0.73). The chance level corresponding to a random classifier, was computed as the ratio between the number of channels pertaining to EZ and the total number of channels in each patient (black dashed line). Symbols and abbreviations: NODE, Nested Outlier Detection; PSD, power spectral density; EZ, epileptogenic zone; PZ, propagation zone; NIZ, non-involved zone; PPV, positive predictive value; TPR, true positive rate; TP, Temporal pole; A, Amygdala complex; B, Hippocampus cephalis; C, Hippocampus caudalis; TB, Temporal basalis; I, Insula; H, Gyrus temporal transverse Heschl; OP, Opercule parietal; OF, Opercule frontal; FT, Front Triangularis; OR, Fronto Orbitaire oblique.

## Results

### Detection and clustering of interictal events

We used the NODE algorithm to detect and cluster interictal events by identifying amplitude outliers across two LFDR thresholds and four frequency bands, with a time discretization of 200 ms defining the time length of each event (see Figure 1A). The variety of the detected interictal events expanded over the maximum number of clusters corresponding to this particular set of NODE parameters (80 clusters, each one characterized by 4-digit label). In the detection and clustering stages, we followed a epileptogenicity-agnostic approach, that is, no *a priori* information about the epileptogenic or physiological nature of the events was introduced during the detection and labeling processes. Then, to assess the epileptogenicity of the detected events we implemented two quantitative strategies, 1) ordering the NODE clusters according to their power to segregate the EZ and NIZ channels across all the patients (see Figure S1), and 2) computing the fraction of epileptiform discharges as visually identified by an epileptologist (FBo) captured by each NODE cluster (see Figure S3). Importantly, consistency was found between these two approaches (see Figures S1 and S3). Figures 1B, 1C and 1D show examples of two subtypes of IEDs given by the clusters 0_09_09_05 and 09_09_09_05 which were associated by the epileptologists with interictal epileptiform spikes and spike-wave complexes respectively, and one type of non-epileptiform events associated with amplitude outliers occurring in the low frequency bands (1 Hz - 30 Hz, Cluster: 09_0_0_0). Interestingly, we found that the events corresponding to low frequency oscillations (e.g. Clusters: 09_0_0_0) were significantly less abundant in the EZ channels with respect to the NIZ channels across all the analyzed patients (see Figures S1C and S1D). Henceforth, we will refer to the events grouped in these clusters as non-epileptic events. Figure S3 shows that only a small fraction of the visually marked spikes corresponds to NODE clusters which were found to be significantly more abundant in NIZ than in EZ across the 35 patients included in the Table 1 (e.g. cluster 0_09_0_0 in Figures S1 and S3). This observation is consistent with the notion that primary spikes associated with EZ and/or abnormal activity have sharper waveform shapes, meaning their spectral decomposition shows more power in the high frequency bands (e.g. clusters 09_09_09_05, 0_09_09_05), when compared to other discharges having smoother waveform morphologies (e.g. cluster 0_09_0_0) which can be in part explained by a propagation mechanism.^5,12,28-30^ Figure 1E, 1F and 1G show the mean rate of events (average over 60 min recording) in each bipolar channel of the patient 7, together with the results of the precision and recall analysis quantifying the capacity of EZ localization associated with each type of event. Importantly, it was found that the subtypes of sharp paroxysmal events identified by the NODE algorithm present important differences in terms of their capacity for the localization of the epileptogenic zone (EZ). In particular, for the patient 7 the clusters 09_09_09_05, 0_09_09_05 and 09_09_09_0 produce F1 max = 0.93, F1 max = 0.73 and F1 max = 0.64 for EZ localization, respectively (see Figures 1E, 1F and Figure S1A). The Figure S1C shows the quantification of the capacity for EZ localization of the 80 NODE clusters across all the analyzed patients.

### Spontaneous fast-ultradian dynamics of the rate of interictal events

We then investigated the temporal dynamics of the interictal events, not necessarily periodic, expanding over time scales ranging from sub-minute up to half an hour (i.e. sub-hour temporal dynamics). For this, we analyzed SEEG recordings as commonly examined in clinics for the treatment of drug-resistant epilepsy (in the range of 5 minutes to 2 hours of SEEG recordings).^5,8,9,26,27^ Figures 2A and 2B show for two patients (7, 13) the cumulative count of all the detected events (CE) across the SEEG channels (each rectilinear segment corresponds to a single bipolar channel). In these plots, the slope of each rectilinear segment correspond to the mean rate of events in that particular bipolar channel (total number of events / whole time period shown in the figures). In order to analyze the fluctuations of the events rate around its mean value, we subtracted a fitted straight line from the CE to obtain the residuals of the CE for each bipolar channel. The resulting detrended count of events (DCE) are presented in Figures 2C and 2D showing a clear oscillatory dynamics of the DCE in particular for the bipolar channels pertaining to the EZ. In the Figures 2C and 2D, the time intervals of the DCE with positive or negative slope correspond to an increase or decrease of the instantaneous rate of events, respectively. Significantly, we found that the interictal dynamics of the rate of polymorphic events observed over fast-ultradian time scales was highly independent of the parameters used in the NODE algorithm (see Figures S4, S5 and S6) and also independent of the specificity of the events detector (compare the fluctuations disclosed by the clusters 0_09_09_05 and 09_09_09_05 with respect to the Spike-like group in the Figures 3A to 3D). These results suggest that the observed dynamics is an intrinsic feature of the interictal neural activity captured by the SEEG recordings, putatively elicited by physiological brain states and/or epileptogenic mechanisms. Then, we analyzed whether the temporal fluctuations of the DCE during the analyzed fast-ultradian time scales entrain the rate of specific epileptogenic biomarkers or not, which could ultimately affect the localization of the EZ. To investigate this, we focused on the temporal evolution of the rate of epileptiform discharges that produced a good EZ localization when averaged over long time intervals (approx. 60 min recording, see Figure 1E and 1F). Figures 2E and 2F show the DCE including all the events as gray dots and highlighting in color the epileptiform discharges pertaining to the cluster 09_09_09_05. Figures 2E and 2F show a clear oscillatory dynamics and rhythmic burst of events pertaining to the cluster 09_09_09_05, respectively. Importantly, Figures 2E and 2F show that 1) the occurrence of the epileptiform discharges (CoI: 09_09_09_05) is specific to the EZ (resulting in more dark red dots than blue dots) and 2) the rate of the epileptiform discharges in the EZ (density of the dark red dots) increases and decreases following the temporal evolution of the DCE with positive slope and negative slope, respectively. These two features can also be identified in the Figures 2G and 2H showing the mean value and standard error of the rate of epileptiform discharges (ER) computed in a sliding epoch of 5 min in length and 90% overlap. The clinically relevant consequence of these observations is exemplified for two patients in the Figures 2I and 2J showing that the performance of EZ localization as quantified by the area under the precision and recall curve (AUPREC for EZ) follows the dynamics of the mean rate of epileptiform discharges. Values fluctuate between close to perfect classification (which would correspond to an AUPREC = 1) and very small values close to chance level (gray filled circles in the Figures 2I and 2J). Importantly, we found that the observed temporal fluctuations of the IEDs rate entraining the precision to localize the EZ over fast-ultradian time scales occurs spontaneously during both the awake and the non-REM sleep states of the patients (see Appendix S1, Section 11). Moreover, the magnitude of the fluctuations of the AUPREC for EZ based on the rate of different subtypes of interictal events disclosed no significant differences at the group level between both awake vs non-REM sleep states, and also between two SEEG recording sessions taken at different days and time of day for the same patient state (awake at rest). For a detailed description of these results the reader is referred to the Appendix S1, Sections 11 and 12. We also investigated the dependence of the observed fluctuations of the AUPREC for EZ on the type of events and on the specificity of the detector of interictal discharges (see Appendix S1, Sections 6 and 7). Figures 2I, 2J, 3A and 3B show that the events pertaining to the clusters 09_09_09_05, 0_09_09_05 and the Spike-like group produce similar temporal fluctuations of the AUPREC for EZ. These results reveal that the temporal dynamics of the events rate during interictal periods is present across different subtypes of epileptiform discharges and effectively entrains the precision to localize the EZ. Figures 3A and 3B show that, as expected, non-epileptic events (blue dots) produce very low values of AUPREC for EZ when compared to the epileptiform discharges (red dots), however, fluctuations of the AUPREC for EZ values are observed in both types of events. Figures 3E and 3F show the absolute difference (AD) between the extreme values (max - min) of the AUPREC for EZ time series based on the rate of IEDs, as a function of the sliding epoch length. Importantly, we found that the magnitude of the fast-ultradian fluctuations of the AUPREC for EZ based on the rate of IEDs decay exponentially as a function of the epoch length with a characteristic time scale *Tao* (i.e. scale-rich process. See Figures 3E and 3F). This scale-rich behavior is essentially different from the temporal dynamics of the AD of AUPREC for EZ based on the non-epileptic interictal events showing a scale-free trend (see Figures 3G and 3H and Appendix S1, Section 8). Besides, the non-epileptic interictal events were found to be significantly more abundant in the NIZ (see Figures S1C and S1D). Of note, the fluctuations of the rate of IEDs in EZ present a scale-free like dependence as a function of the epoch length (linear trend in a log-log plots shown in Figures S2I and S2J), which is different from the exponential trend with a characteristic time scale *Tao* disclosed by the fluctuations of the corresponding AUPREC for EZ (see the linear trend in the log-linear plots shown in Figures S2A, 3E and S2B, 3F). The latter, suggests that the observed fast-ultradian fluctuations of the AUPREC for EZ can not be completely explained by considering it simply as a function of the IEDs rate fluctuations within the EZ (see the caption of the Figure S2). Taken together, these results suggest that overlooking the spontaneous fast-ultradian dynamics of IEDs can produce incomplete and/or misleading information, leading to a suboptimal delineation of epileptogenic targets, and this holds true regardless of the specificity of the IEDs detector included in the processing pipeline (compare the fluctuations disclosed by the clusters 0_09_09_05 and 09_09_09_05 with respect to the Spike-like group in the Figures 3A to 3D).

**Figure 2:**
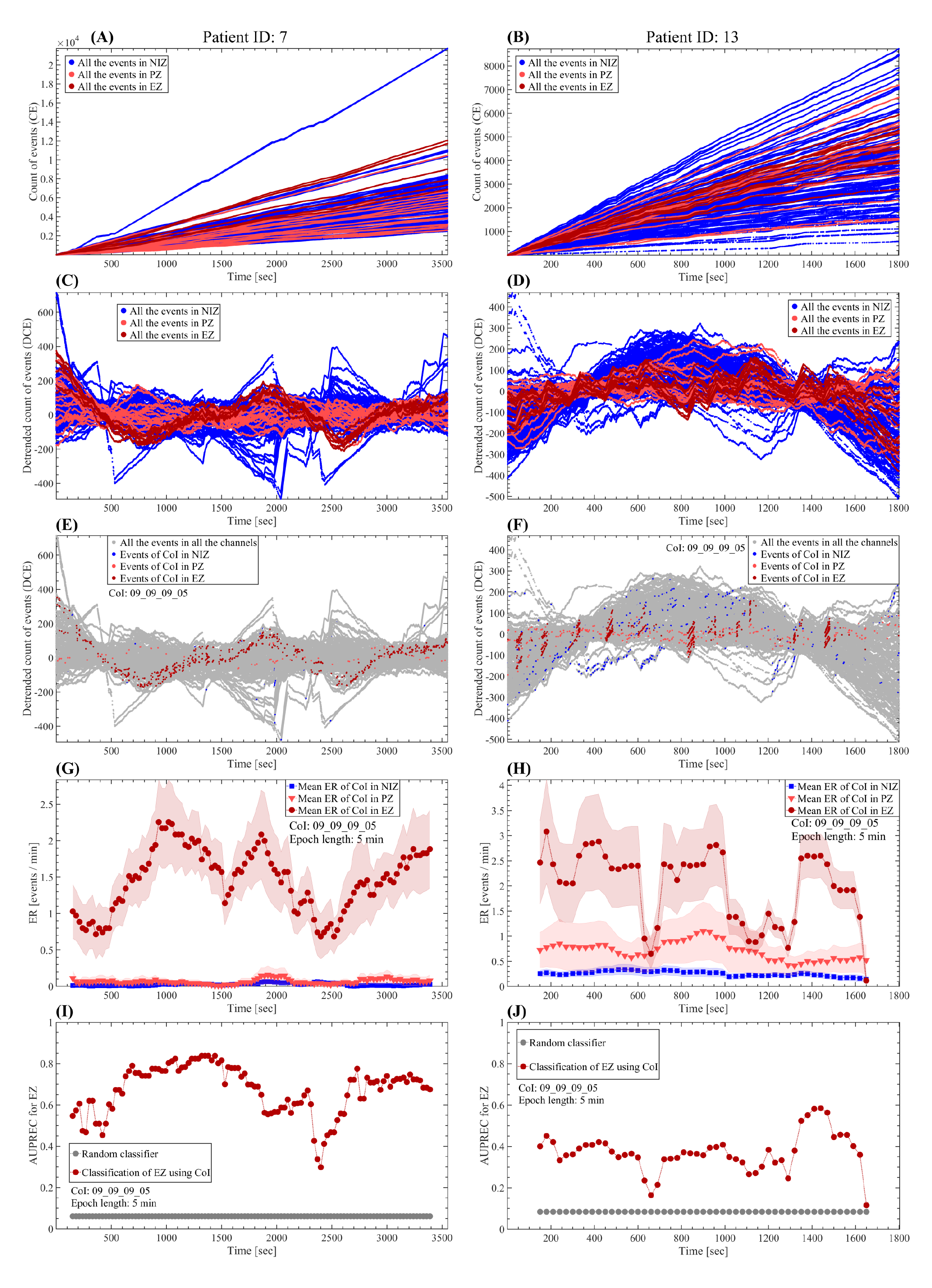
Spontaneous temporal dynamics of the rate of interictal events. **(A, B)** Cumulative count of events including all events subtypes (epileptic and non-epileptic) detected by the NODE algorithm (80 clusters). Each line correspond to a single bipolar channel and the slope of each line (number of events / time) is the mean rate of events for that particular SEEG channel. **(C, D)** Detrended cumulative count of events (cumulative residual). **(E, F)** Cumulative residual showing all the detected events (gay dots) and the discharges pertaining to the cluster 09_09_09_05 in color (NIZ: blue dots, PZ: light red dots, EZ: dark red dots). **(G, H)** Mean rate of events pertaining to the cluster 09_09_09_05. The dots correspond to the mean value of the events rate at each time position of the sliding epoch of 5 min in length and 90% overlap. The shaded error bars correspond to the standard error. **(I, J)** AUPREC for EZ localization based on the rate of events pertaining to the cluster 09_09_09_05 at each time position of the 5 min length sliding epoch. Symbols and abbreviations: CoI, cluster of interest; EZ, epileptogenic zone; PZ, propagation zone; NIZ, non-involved zone; ER, events rate; AUPREC, area under the precision and recall curve.

**Figure 3:**
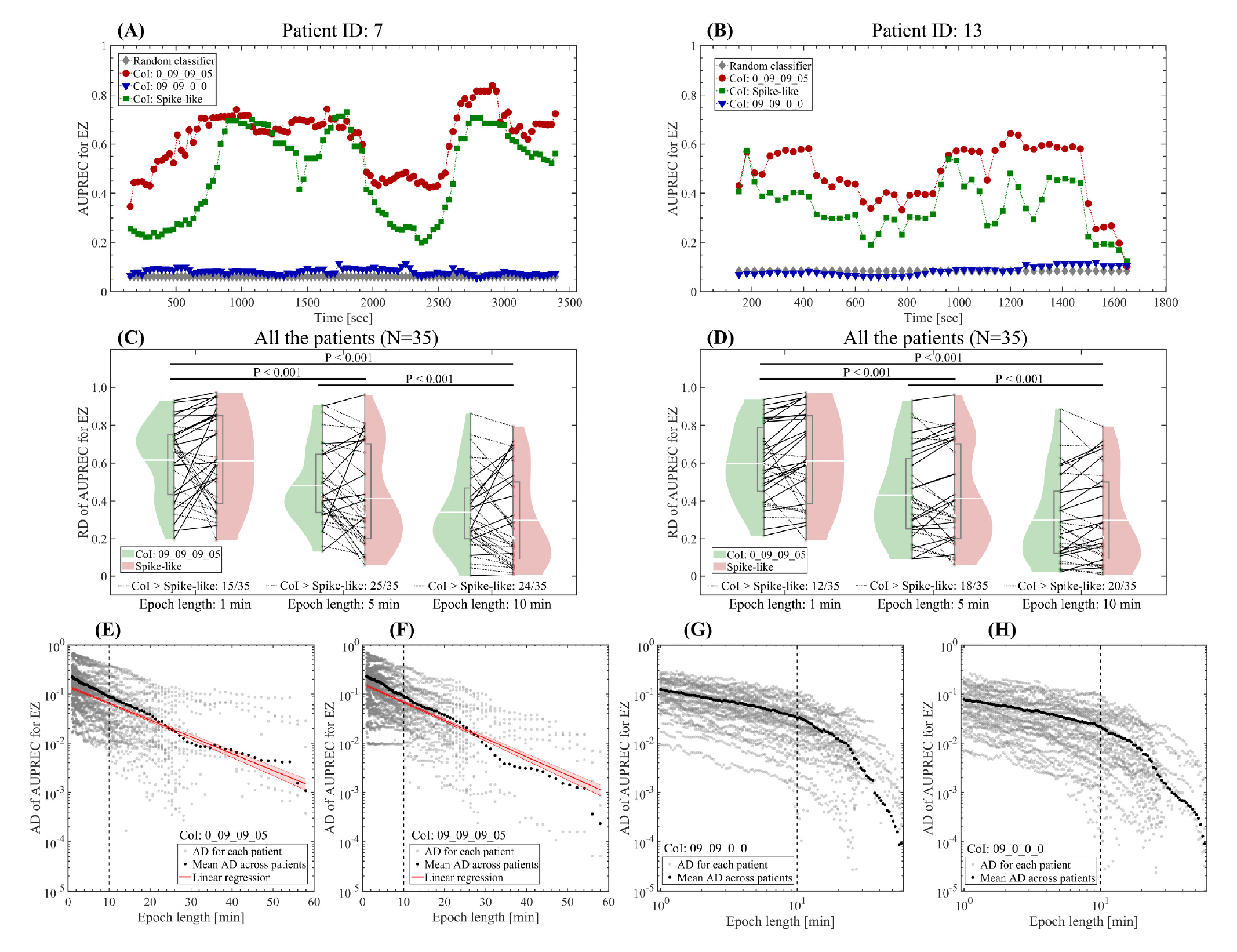
Temporal dynamics of the goodness of EZ localization as quantified by the AUPREC based on the rate of interictal events. **(A, B)** Temporal evolution of the AUPREC for EZ localization based on the rate of IEDs (CoI: 0_09_09_05, Spike-like) and non-epileptic events (CoI: 09_09_0_0) computed using a sliding epoch of 5 min in length and 90% overlap. **(C, D)** Violin plots including all the patients paired across two CoIs showing the relative difference (RD) between the extreme values ((max - min) / max) of the AUPREC for EZ time series for three lengths of the sliding epoch (1 min, 5 min, 10 min). In all the cases, the overlap of the sliding epoch was 90% and it sweeps the whole interictal period available in each patient. In an inter-group paired analysis, the reported P values indicate significant differences across the three sliding epoch lengths (Wilcoxon signed rank test with the P values Bonferroni-adjusted to correct for multiple comparisons across the 3 epoch lengths). In an intra-group paired analysis, only the 5 min epoch length in panel (C) presents significant difference between the distributions corresponding to CoI: 0_09_09_05 and Spike-like (P = 0.01, Wilcoxon signed rank test). **(E, F)** Log-linear plots showing the absolute difference (AD) between the extreme values (max - min) of the AUPREC for EZ based on the rate of IEDs as a function of the sliding epoch length. The red line and red shaded error bars represent the linear regression and the 95% confidence interval, respectively. These linear trends in the log-linear plots suggest an exponential dependence *y ∝ e^-x/Tao^* where *y* represents the AD of AUPREC for EZ being proportional to a decaying exponential function of the epoch length *x*, with a characteristic time scale *Tao*. For CoI: 0_09_09_05 (panel E) we obtain: Slope = −0.079 [1/min] (*Tao* = 12.7 [min]), SE = 0.002 [1/min], P < 0.001, t-statistic of the two-sided hypothesis test. For CoI: 09_09_09_05 (panel F) we obtain: Slope = −0.086 [1/min] (*Tao* = 11.7 [min]), SE = 0.002 [1/min], P < 0.001, t-statistic of the two-sided hypothesis test. The AD of AUPREC for EZ presents a linear dependence in a log-linear plot suggesting an exponential trend, with a characteristic time scale *Tao*. **(G, H)** Log-log plots showing the AD of AUPREC for EZ based on the rate of the non-epileptic events as a function of the sliding epoch length. The AD of AUPREC for EZ presents 2 linear segments with different behavior below and above ≈ 10 min epoch length. This linear dependence in a log-log plot suggests a scale-free dynamics. Symbols and abbreviations: CoI, cluster of interest; IEDs, interictal epileptogenic discharges; EZ, epileptogenic zone; PZ, propagation zone; NIZ, non-involved zone; AD, absolute difference; AUPREC, area under the precision and recall curve.

### Putative mechanisms linked to the fast-ultradian dynamics of the rate of interictal events

The temporal fluctuation of the rate of IEDs in a given brain region can originate from variations in excitability within this region. However, it could also be explained - in a complementary way - by fluctuations in propagation from a primary irritative zone to a secondary zone. ^28-30^ In order to test the propagation hypothesis, we correlated the event rates between the EZ and the PZ (see Appendix S1, Section 5). It is important to note that the propagation of IEDs across macroscopic networks occurs within sub-second time scales (approx. 100 ms from temporal to frontal regions).^12,13^ As a consequence, this fast propagation should not influence the measure of zero-lag correlation between event rates averaged over sliding epochs of several minutes in length (see Appendix S1, Section 5). Following this reasoning, we defined two criteria associated with the propagation mechanism in order to quantitatively test this hypothesis over the fast-ultradian time scales: 1) the dynamics of the mean rate of IEDs in EZ should positively correlate with the dynamics of the mean rate of the same subtype of IEDs in PZ and 2) the AUPREC for EZ time series should negatively correlate with the dynamics of mean rate of IEDs in PZ (i.e., the occurrence of IEDs in PZ effectively impairs the EZ localization). Figure 4 shows the quantification of the two criteria associated with the propagation mechanism for three type of events and across three sliding epoch lengths. The percentage of patients satisfying the two criteria associated with the propagation mechanism is up to approx. 23% (8/35), 14% (5/35) and 11% (4/35) for epoch lengths of 1 min, 5 min and 10 min, respectively. These results strongly suggest that the propagation mechanism could have a dominant role in explaining the interictal fluctuations of the AUPREC for EZ time series only in a limited fraction of the analyzed patients. This suggests that the excitability of the epileptogenic tissues, along with other factors such as the brain state and the probed brain region, could play a more relevant role in explaining the spontaneous dynamics of the IEDs rate constraining the precision to localize the EZ over fast-ultradian time scales. Further details related to the propagation mechanism are discussed in the Appendix S1, Section 9.

**Figure 4:**
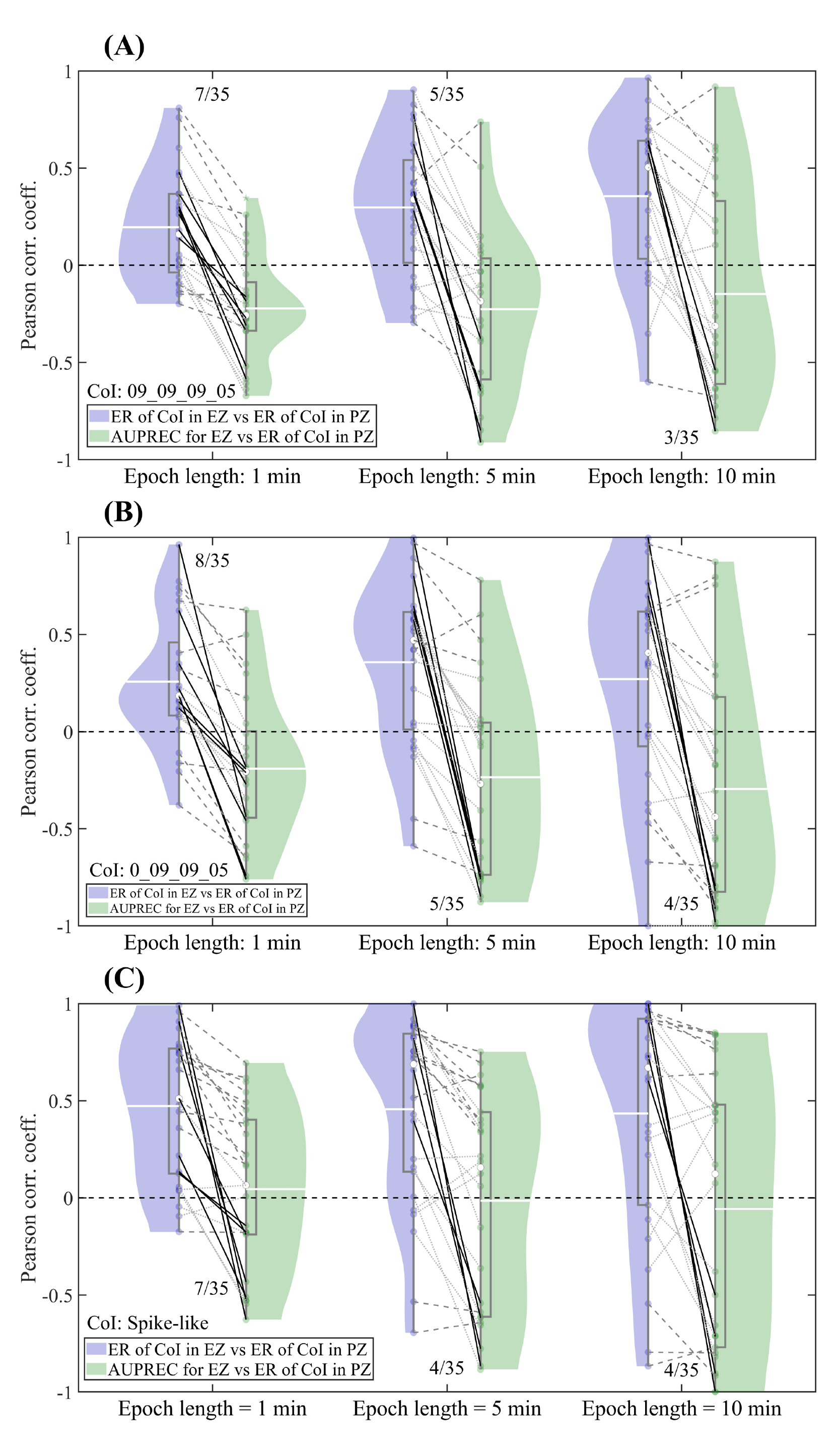
Testing the hypothesis that the temporal fluctuations of the AUPREC for EZ on fast-ultradian time scales are associated with the propagation of IEDs from EZ to PZ. Panels **(A)**, **(B)** and **(C)** correspond to the clusters 09_09_09_05, 0_09_09_05 and Spike-like, respectively. In each panel, the violin plots show all the patients paired across two correlations (ER of CoI in EZ vs ER of CoI in PZ) and (AUPREC for EZ vs ER of CoI in PZ). The three time series involved in the correlations (ER of CoI in EZ, ER of CoI in PZ and AUPREC for EZ) where computed using sliding epoch of length 1 min (left panel), 5 min (central panel) and 10 min (right panel) and 90% overlap scanning the whole interictal period available in each patient. Dotted gray lines correspond to patients in which at least one of the two correlations result no significant. Dashed gray lines correspond to patients in which the two correlations are significant, but the signs of the correlations do not satisfy the criteria associated with the propagation hypothesis (see discussion in the text). Solid black lines correspond to patients in which the two correlations are significant and satisfy the criteria associated with the propagation hypothesis (see discussion in the text). The fractional number accompanying each paired violin plot indicates the fraction of patients satisfying all the criteria associated with the propagation hypothesis. The statistical significance of the correlations (P < 0.05) was assessed by using the Student’s t distributions of the two-tailed hypothesis test under the null hypothesis that the correlation is zero. Symbols and abbreviations: CoI, cluster of interest; IEDs, interictal epileptogenic discharges; EZ, epileptogenic zone; PZ, propagation zone; NIZ, non-involved zone; ER, events rate; AUPREC, area under the precision and recall curve; ER of CoI in EZ/PZ, mean rate of events pertaining to the CoI averaged over the EZ/PZ channels.

### Predicting the spontaneous fast-ultradian dynamics to improve the epileptogenic zone localization

In this section we propose a strategy to prospectively estimate, i.e. without knowing the ground truth classification of EZ, the interictal epoch for near-optimal EZ localization based on IEDs rate. For this, we quantitatively define the optimal EZ localization for each patient as that corresponding to the maximum value of the AUPREC for EZ time series which is characterized by a temporal fluctuations over fast-ultradian time scales (see Figures 2I, 2J, 3A and 3B). Accordingly, the proposed method aims at estimating the time position of the sliding epoch corresponding to the maximum value of the AUPREC for EZ time series computed based on the rate of a given subtype of IEDs. The violin plot corresponding to the case "Best 5 min epoch (Max AUPREC)" shown in Figure 5D, summarizes the optimal EZ localization in each patient based on an epoch length of 5 min and IEDs pertaining to the cluster 09_09_09_05. We started by considering the fact that the interictal temporal dynamics of the AUPREC for EZ highly correlates with that associated with the mean rate of IEDs in EZ. Figures 2E to 2J illustrate this phenomenon for two particular patients (see also Figures S13 to S16 and S19 to S22). Moreover, Figures 5A and 5B show that the correlation between the two time series: AUPREC for EZ and "ER of IEDs (CoI: 09_09_09_05) in EZ", is positive and statistically significant in 77% (27/35) of the patients. Then, we investigated possible measures correlating with the AUPREC for EZ time series and suitable to be applied in a prospective manner. This requires the measures to be independent of the EZ, PZ, NIZ ground truth classification of the SEEG channels. We found that the time series corresponding to the mean rate of events including all the clusters and averaged across all the channels (ER of all Clust in all Chan) negatively correlates with the AUPREC for EZ time series computed using these IEDs subtypes (e.g. clusters 0_09_09_05 and 09_09_09_05 associated by the epileptologists with interictal epileptiform spikes and spike-wave complexes respectively). Note that the overall rate of interictal events "ER of all Clust in all Chan" across all the SEEG channels is a suitable measures for prospective analysis since they do not depend on the SEEG channels classification in EZ, PZ, NIZ. Figures 5B and 5C show that the correlation between the time series AUPREC for EZ (CoI: 09_09_09_05) and "ER of all Clust in all Chan", is negative and statistically significant in 63% (22/35) of the patients. A discussion regarding the putative mechanisms underlying this correlations is given in the Appendix S1, Section 10.

**Figure 5:**
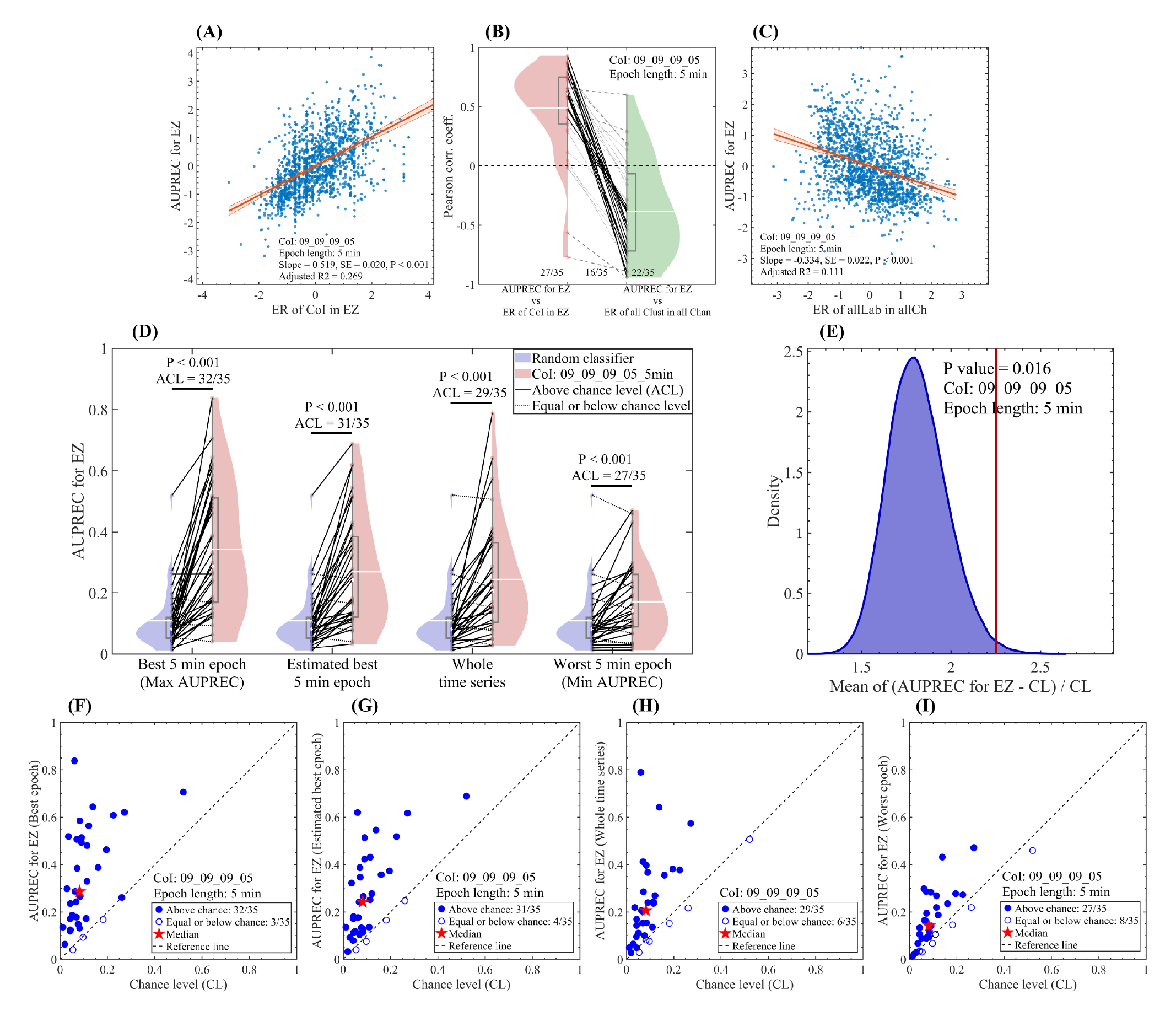
Estimating the best interictal 5 min-epoch for near-optimal EZ localization. **(A)** Scatter plot showing the correlation between the time series AUPREC for EZ and ER of CoI in EZ. **(B)** Scatter plot showing the correlation between the time series AUPREC for EZ and ER of all Clust in all Chan. In panels **(A)** and **(C)**, all the measures were computed for the CoI: 09_09_09_05 and each dot correspond to the measure value in a particular time position of the sliding epoch of 5 min in length and 90% overlap, covering the whole interictal SEEG time series available in each patient. The red line and red shaded error bars represent the linear regression and the 95% confidence interval, respectively. For panel **(A)** we obtained: Slope = +0.519 [1/min], SE = 0.020 [1/min], P < 0.001, t-statistic of the two-sided hypothesis test. For panel **(C)** we obtained: Slope = −0.334 [1/min], SE = 0.022 [1/min], P < 0.001, t-statistic of the two-sided hypothesis test. **(B)** Violin plots showing all the patients paired across the two correlations, 1) AUPREC for EZ vs ER of CoI in EZ, and 2) AUPREC for EZ vs ER of all Clust in all Chan. Dotted gray lines correspond to patients in which at least one of the two correlations is no significant. Dashed gray lines correspond to patients in which the two correlations are significant and, negative in the red violin plot and positive in the green violin plot, or have the same sign. Solid black lines correspond to patients in which the two correlations are significant and, positive in the red violin plot and negative in the green violin plot. The three fractional numbers accompanying the paired violin plots indicate the fraction of patients presenting significant value for the left-hand, both and right-hand correlations. The statistical significance of the correlations (P < 0.05) was assessed by using the Student’s t distributions of the two-tailed hypothesis test under the null hypothesis that the correlation is zero. **(D)** Violin plots showing all the patients paired across the values of AUPREC for EZ based on a random classifier, computed as the ratio between the number of channels pertaining to EZ and the total number of channels in each patient (blue violin plot), and the AUPREC for EZ based on the rate of events pertaining to the cluster 09_09_09_05 (red violin plot). In an intra-group paired analysis (Wilcoxon signed rank test), the reported P values indicate significant differences between the two distributions of AUPREC values in all the four cases shown. In an inter-group paired analysis, we found significant differences (P < 0.001) between all but one group pair: Estimated best 5 min epoch vs Whole time series (P = 0.084, Wilcoxon signed rank test with the P values Bonferroni-adjusted to correct for multiple comparisons across the 4 cases). **(E)** Histogram showing the distribution of the relative difference of the AUPREC for EZ with respect to the chance level (CL) for a 5 min epoch randomly sampled from the interictal SEEG recordings of each patient (10^5^ random samplings). The CL was computed as the ratio between the number of channels pertaining to EZ and the total number of channels in each patient. The red vertical solid line shown in the histogram indicates the relative difference value corresponding to the estimated best 5 min epoch for near-optimal EZ localization (second case from the left in panel **D**). **(F, G, H, I)** Scatter plots corresponding to the four cases shown in panel **(D)**. In panels D, E and G, the estimated best 5 min epoch for near-optimal EZ localization corresponds to the interictal epoch producing the minimum value of ER of all Clust in all Chan. Symbols and abbreviations: CoI, cluster of interest; EZ, epileptogenic zone; ER, events rate; AUPREC, area under the precision and recall curve. ACL, above chance level; ER of CoI in EZ/PZ, mean rate of events pertaining to the CoI averaged over the EZ/PZ channels; ER of all Clust in all Chan, mean rate of events including all the clusters and averaged across all the channels.

Based on these findings we estimated the epoch for near-optimal EZ localization as the interictal epoch corresponding to the minimum value of the "ER of all Clust in all Chan". For the IEDs corresponding to the cluster 09_09_09_05, Figures 5D, 5G and 5H show that the 5 min-epoch estimated in each patient using the proposed method produces at the population level an EZ localization slightly better than that obtained using the whole time series available in each patient (Bonferroni-adjusted P = 0.084, non-parameteric Wilcoxon signed rank test). Besides, these two cases (estimated best 5 min-epoch and whole time series) perform significantly worst and better than the cases for the actual best 5 min-epoch (i.e. maximum value of the AUPREC for EZ time series in each patient) and the worst 5 min-epoch (i.e, minimum value of the AUPREC for EZ time series in each patient), respectively (see Figures 5D and 5F to 5I). Figures 5E shows that the 5 min-epoch estimated in each patient using the proposed method produces an EZ localization at the population level significantly better with respect to that obtained with a 5 min epoch randomly sampled from the interictal recordings of each patient (P = 0.016, 10^5^ random samplings). Notably, we found that for the EZ and RZ localization using the cluster 09_09_09_05, the proposed method performs at the population level significantly better with respect to the approach based on a randomly sampled epoch, for epoch lengths in the range 5 - 10 min (in all the cases we obtained P < 0.05, 10^5^ random samplings, see Figures 5 and S12). The predictive performance for near-optimal EZ and RZ localization of all the NODE clusters is discussed in the Appendix S1, Section 10 in connection with the Figures S8 to S11.

## Discussion

### Spontaneous fast-ultradian dynamics of the rate of interictal events

In this study, we show that the spontaneous dynamics of different subtypes of IEDs observed over fast-ultradian time scales emerge as an intrinsic feature of these EZ biomarkers, defined by the occurrence of amplitude outliers across frequency bands of interest (see Figure 1). This approach allowed us to investigate the temporal dynamics of a massive number of interictal events (see Figures 2A to 2D), including several subtypes of epileptiform discharges predominantly occurring in EZ as well as non-epileptic events associated with amplitude outliers in low frequency bands (1 Hz - 30Hz). These latter were found to be significantly less abundant in EZ with respect to NIZ (see Figure 1, S1 and S3). The analysis of the rate of these interictal events revealed underlying temporal dynamics characterized by sub-hour time scales which includes, but is not exclusive of, the IEDs observed in the EZ (compare Figures 2C vs 2D and Figures 2E vs 2F). Crucially, we found that the observed dynamics of the rate of events is an intrinsic feature of the interictal brain activity captured by the SEEG traces. That is, the interictal dynamics of the rate of polymorphic events is not, or at least can not be explained solely by, an epiphenomenon associated with a particular parameters configuration of the NODE algorithm (see Figures S4, S5 and S6). We found that the NODE algorithm segregates the interictal epileptiform spikes visually marked by an epileptologist (FBo) into different sub-clusters (see Figure S3). Whereas most of visually marked spikes correspond to NODE clusters found to be significantly more abundant in EZ than in NIZ (e.g. 09_09_09_05, 0_09_09_05), leading to a good EZ localization (see Figure S1), they showed different characteristics regarding their temporal dynamics. Firstly, we found that the rate of different types of interictal epileptogenic discharges undergo temporal fluctuations over fast-ultradian time scales as commonly examined in clinics. Previous studies have reported that the rate of interictal epileptic spikes increase during wakefulness and sleep,^10,14,15,17,18^ and is suppressed during attention and memory tasks.^31-34^ In contrast, the fast-ultradian temporal dynamics reported in this work was observed and characterized preoperatively in the interictal SEEG traces of 35 patients with Engel I seizure outcome and appears to occur spontaneously, that is, not triggered by or exclusively associated with a particular cognitive task, wakefulness, sleep, seizure occurrence, post-ictal state or antiepileptic drug withdrawal (see subsection "Patients and intracerebral recordings" in Methods and Sections 11 and 12 of the Appendix S1). Secondly, we found that the minimization of the rate of interictal events (epileptic and non-epileptic) across all the SEEG channels is a good predictor of the interictal epoch (in the range 5 - 10 min in length) for near-optimal EZ localization based on specific subtypes of IEDs. Notably, we found that for the RZ localization based on the IEDs subtype 09_09_09_05, the proposed method performs at the population level significantly better than both 1) using the whole time series available in each patient and 2) using a short epoch (length ≈ 5 - 10 min) randomly sampled from the interictal recordings of each patient. Of note, this novel and counter-intuitive link between the dynamics of the overall rate of polymorphic events and the rate of specific subtypes epileptiform spikes which was exploited here to improve the EZ localization would be very difficult, if not impossible, to unveil by solely considering a limited subtype of events (e.g. visually marked epileptic spikes). Regarding the mechanisms underlying the observed fast-ultradian dynamics, our results suggest that the propagation mechanism could have a dominant effect only in a limited fraction of the analyzed patients (see Figure 4), suggesting that the excitability of the epileptogenic tissue along with other factors such as the brain state and the probed brain region could play a more relevant role in explaining the spontaneous temporal dynamics of the IEDs rate entraining the precision to localize the EZ over sub-hour time scales.

Taken together, our results show that the rate of different subtypes of IEDs spontaneously undergoes temporal fluctuations which effectively constraint the precision to localize the EZ over fast-ultradian time scales as commonly analyzed in clinics,^5,8^ hence, representing a clinically relevant factor for surgical planning in drug-resistant epilepsy.

### Limitations

Whereas all the patients included in the study where seizure-free after surgery, indicating that the analyzed SEEG traces effectively capture the brain activity associated with EZ, further investigation is warranted to disentangle the observed dynamics of interictal events from the heterogeneity and variable number of explored brain regions. Besides, given the heterogeneous exposure to antiepileptic drugs of the analyzed patients, further investigation is required to assess the effect of this factor on the fast-ultradian dynamics of the rate of interictal events. Importantly, the possibility that the fast-ultradian fluctuations of the rate of interictal events analyzed in this work could vary with or be hierarchically coupled to another rhythms with longer time periods (e.g. circadian rhythms), deserve further investigation. However, this does not change the conclusions of our study regarding the existence of the fast-ultradian fluctuations of the rate of interictal events entraining the presicion to localize the EZ, and its importance for surgical planning in drug-resistant epilepsy.

## Conclusion

A major limitation of previous works assessing the performance of epileptogenic biomarkers like interictal spike and HFOs, refers to the underestimation (or total neglect) of the time-varying factors underlying the genesis of these biomarkers.^5,8,9,26,27^ This limitation can effectively produce incomplete and/or misleading information, leading to a suboptimal delineation of epileptogenic targets. In this study, we provided for the first time a method to investigate and predict the spontaneous fast-ultradian dynamics of interictal transient event-like biomarkers. Based on our results, the recommendation for clinical applications is that an interictal time interval of at least 30 min in length is required to stabilize the findings, i.e. in order to produce a relative attenuation of 90% of the AUPREC for EZ fluctuations with respect to that observed in the case of a 5 min interictal time window (see the calculations in Appendix S1, Section 8). The proposed method for polymorphic events analysis paves the way to prospectively estimate the interictal time interval for near-optimal EZ localization in a substantial fraction of the analyzed patients based solely on the dynamics of the intracranial recordings (see Figure 5B). This fact reveals a novel and counter-intuitive link between the fast-ultradian dynamics of the overall rate of polymorphic events (epileptic and non-epileptic) and the rate of specific subtypes of epileptiform spikes as an intrinsic feature of the interictal brain activity (see Figures 5 and S7 to S12). This result is clinically relevant as we showed how it can be exploited to improve the EZ localization and also offer a novel approach to investigate the time-varying factors underlying the genesis of different subtypes of IEDs.

## Data Availability

The data and code that support the findings of this study are available from the corresponding author, upon reasonable request.

https://github.com/damian-dellavale/node/

## Acknowledgments

This work was supported by the ANR Neurosense ANR-18-CE19-0013 grant, and by the RHU EPINOV [A*MIDEX project (ANR-17-RHUS-0004) funded by the ’Investissements d’Avenir’ French Government].

## Authors contribution

DD contributed to the conceptualization, methodology, formal analysis, writing the original draft and figures preparation. FBo, FP, CGB and FB contributed to the data acquisition and visual analysis of the recordings, review and editing the manuscript. JM contributed to the acquisition and curation of the dataset. FW, JMB, FB, CGB contributed to the funding acquisition.

## Ethical Publication Statement

We confirm that we have read the Journal’s position on issues involved in ethical publication and affirm that this report is consistent with those guidelines.

## Data availability

The data and code that support the findings of this study are available from the corresponding author, upon reasonable request. In particular, the source code for the computation of the NODE algorithm together with test script examples are freely available at https://github.com/damian-dellavale/node/

We are willing to provide technical support to investigators who express an interest in implementing the NODE algorithm in other programming languages, integrate it in open-source software toolboxes, or use it for non-profit research activities.

## Supporting information

Additional Supporting Information may be found in the online version of this article:

**Appendix S1.** Supplementary methods and results.

## Appendix S1: Supplementary methods and results

## 1. Classification of the SEEG contacts

In this work we used the historical definition of EZ^1-3^, which is the site of the beginning and of the primary organization of the seizure. Thus, the historical definition of the EZ used in the current study is different from the one of Lüders et al.^4,5^: "the minimum amount of cortex that must be resected (inactivated or completely disconnected) to produce seizure freedom." Specifically, the classification of the SEEG contacts in EZ, propagation zone (PZ) and non-involved zone (NIZ) was made according to the sites of seizure initiation (SOZ) based on visual analysis and on the Epileptogenicity Index (EI).^6-8^ Channels exhibiting EI values above 0.1 to 0.2 were considered to pertain to the PZ. Besides, channels exhibiting EI values above 0.3 to 0.4 were considered to be epileptogenic. By binarizing the EI using these thresholds we obtained an estimation of the extent of the PZ and EZ. Then, the final classification was obtained by combining the EI information with the visually defined SOZ, which includes the identification of the contacts where the earlier and delayed ical activity were seen. Further details about the method used here for classifying the SEEG channels have can be found in previous works^7-10^.

## 2. Detection and clustering of polymorphic events

In this section, we start by describing the NODE (Nested Outlier Detection) algorithm and how it defines the interictal events. Then, we define the cluster, comparison and labeling constraints associated with the semi-supervised constrained clustering approach used to group the polymorphic events in clusters. Illustrative examples to help the conceptual interpretation of the labels assigned to the interictal events by the NODE algorithm, are presented in the main text (see Methods, subsection "Data and statistical analysis").

### The NODE algorithm

We defined interictal events as anomalies (i.e. outliers of amplitude) in the LFP time series using the Nested Outlier Detection (NODE) algorithm. Figure 1A schematize the main processing steps associated with the NODE algorithm. Briefly, on each SEEG channel we applied the following series of steps. In step 1, we band-pass filtered the whole raw LFP time series in the frequency bands of interest. In step 2, we use the Local False Discovery Rate (LFDR) method^11,12^ to detect outliers present in the distribution of amplitude values within each frequency band in a controlled manner.

Specifically, the LFDR method allows to define amplitude thresholds for detecting outliers of amplitude while controlling for the proportion of false positives. In this regard, the NODE algorithm take as an input multiple thresholds to detect amplitude outliers which are not absolute values of amplitude, but rather rates of false discovery (LFDR threshold). From these LFDR threshold values the corresponding amplitude thresholds are computed through the LFDR technique by processing the whole distribution of amplitudes of the band-pass filtered time series under analysis. An outlier of amplitude is detected each time the amplitude envelope of the time series band-pass filtered around the frequency band of interest exceeds one of the amplitude thresholds defined by the LFDR method (see the small black triangles in Figure 1A). Once the amplitude outliers are identified in each frequency band, in step 3 the NODE algorithm merges in a single event the amplitude outliers which co-occur across the frequency bands within a time window of 200 ms. Note that this time window is used to merge the outliers, hence, imposing a lower limit on how close two adjacent events can be to each other. However, this time window is not involved in the step 1 of the NODE algorithm which would produce significant edge effects associated with the band-pass filtering. In step 4, a *Nfb* digits label is assigned to each event, where *Nfb* is the number of frequency bands analyzed by the NODE algorithm. The *Nfb* digits of the label are computed as 1 - *Tfb*, where *Tfb* is the lower LFDR threshold value crossed by the amplitude outlier within each frequency band. Thus, *Tfb* represents the proportion of the detected anomalies that can be expected to be false positives, and the other 1 - *Tfb* fraction being genuine true discoveries.^11,12^ As a result, each one of the *Nfb* digits of the label can be interpreted as the proportion of the detected anomalies that can be expected to be true outliers in the corresponding frequency band. That is, the *Nfb* digits of the label can be interpreted using a positive logic in which the higher the value of the digit the higher the proportion of true outliers associated with the corresponding frequency band. In this work we use four frequency bands (*Nfb* = 4) and two LFDR thresholds {0.5, 0.1}, resulting in 4-digits labels where each digit can adopt the values 0, 0.5 or 0.9 corresponding to amplitude outliers crossing none (0), the **higher** (1 - *Tfb =* 1 - **0.5** = 0.5) and the **lower** (1 - *Tfb =* 1 - **0.1** = 0.9) LFDR threshold value (*Tfb*), respectively (see Figure 1A). Finally, in step 5 the events were grouped in clusters based on the assigned labels. For *Nfb* = 4 frequency bands and two LFDR thresholds we have a total of 80 possible clusters each one characterized by a 4-digits label.

### Constrained clustering

From a machine learning point of view, the NODE algorithm can be used in an unconstrained manner by using fine-grained bins of frequency and amplitude in order to find the clusters naturally emerging from the data, however, for our clinical application we decided to follow a more computationally efficient strategy. That is, the particular implementation of the NODE algorithm used in this work is based on a semi-supervised approach for clustering data while incorporating domain knowledge in the form of constraints, known as constrained clustering.^13^

### Cluster constraints

We included two cluster constraints in this particular implementation of the NODE algorithm. The first constraint refers to the frequency bands of interest which were defined based on the *a priori* information about the transient waveform shape of interest. Specifically, the low [1 Hz - 10 Hz] and high [150 Hz - 255 Hz] frequency bands were included as features characterizing the waveform shapes associated with sharp spikes and spike-wave complexes. Besides, two medium frequency bands [8 Hz - 32 Hz], [30 Hz - 155 Hz] were included with the aim to differentiate subtypes of spikes (e.g. epileptiform sharp spikes from smoother transients). The second constraint refers to the number of LFDR thresholds (i.e. bins of amplitude). In this case we consider that the relevant feature is the occurrence of outliers across the frequency bands, and that the particular amplitude of the outliers do not carry relevant information. Accordingly, we followed a coarse-grained approach by defining only two LFDR thresholds {0.5, 0.1} in each frequency band, representing a proportion of {50%, 90%} of the detected anomalies that can be expected to be true outliers in the corresponding frequency band, respectively. In general, the introduction of constraints restrict the diversity of the resulting clusters in favour of algorithmic efficiency. The two clusters constraints described above restrict the diversity to 80 possible clusters each one characterized by a 4-digits label. Importantly, we explored a wide range of parameters of the NODE algorithm and verified that these cluster constraints do not significantly affect the spontaneous dynamics of polymorphic events observed over fast-ultradian time scales (see Figures S4 to S6). We found that the spontaneous dynamics of the interictal events rate results highly robust with respect the relaxation of the clustering constraints associated with the number and range of the frequency bands. In particular, the rhythmic dynamics of the polymorphic events rate is clearly distinguishable even using a very aggressive constraint in the number of frequency bands. Figure S6 shows that only in the case where a very high number of noisy anomalies are allowed (e.g. > 90% of LFDR), the rhythmic behavior of the events rate dynamics becomes almost indistinguishable from the random fluctuations (see Figures S6E and S6F).

### Comparison constraints

Since the labels assigned by the NODE algorithm are constituted by numerical digits, it is possible to compute a variety of distance measures to quantify the difference between the polymorphic events. In this work, we used a simple similarity criteria to conform the clusters consisting in grouping the events with identical 4-digits label (i.e. each cluster is defined by all the events producing zero euclidean distance between their labels). Importantly, we found that changing the parameters of the NODE algorithm, and thus the way in which the events are grouped in clusters, does not significantly affect the fluctuations of the rate of events observed over the fast-ultradian time scales (see Figures S4 to S6).

### Labeling constraints

We followed a epileptogenicity-agnostic approach for clustering, that is, no *a priori* information about the epileptogenic or physiological nature of the labels assigned to each type of event was introduced during the clustering processes (i.e. no labeling constraints). Note that this strategy is essentially different from the commonly used approach based on a binary classification (epileptic vs non-epileptic events) in which the events not meeting certain particular epileptogenicity criteria are excluded from any further analysis. In this study we used a different strategy based on epileptogenicity-agnostic approach to detect and cluster the events. Subsequently, the epileptogenicity of the detected events was assessed using two quantitative strategies, 1) ordering the NODE clusters according to their power to segregate the EZ and NIZ channels across all the patients (see Figure S1), and 2) computing the fraction of epileptiform discharges as visually identified by an epileptologist (FBo) captured by each NODE cluster (see Figure S3). See also the discussion in the section "Detection and clustering of interictal events" of the main text, in connection with the clusters 0_09_09_05, 09_09_09_05 and 09_0_0_0 shown in the Figure 1B to 1G. Importantly, regardless of their epileptogenicity, all the events subtypes corresponding to the 80 NODE clusters were included in our analysis. The latter, paves the way to unveil a novel and counter-intuitive link between the dynamics of the overall rate of polymorphic events and the rate of specific subtypes of epileptiform spikes which was exploited here to improve the EZ localization (see Figures 5, S7, S12 and the discussion in the section "Predicting the spontaneous fast-ultradian dynamics to improve the epileptogenic zone localization" of the main text). Of note, this correlation between the temporal patterns associated with different events subtypes captured by the NODE clusters would be very difficult to unveil by solely considering a limited subtype of events (e.g. visually marked epileptic spikes).

As a result, the two stages associated with the **semi-supervised constrained clustering** method used in this study can be described as follows:

A. **Unsupervised** epileptogenicity-agnostic clustering based on **no labeling constraints** and zero euclidean distance between the labels as a comparison constraint.
B. The clusters obtained in A) are reviewed (i.e. **supervised**) by the epileptologists in order to assess their epileptogenicity. In this regard, two quantitative strategies were also implemented, 1) ordering the NODE clusters according to their power to segregate the EZ and NIZ channels across all the patients (see Figure S1), and 2) computing the fraction of epileptiform discharges as visually identified by an epileptologist (FBo) captured by each NODE cluster (see Figure S3).

## 3. Time-frequency analysis

Time-frequency maps of the polymorphic events were computed as scalograms using Morlet wavelets including spectral whitening by Z_H0_-score normalization of each frequency bin across time samples.^14^ Specifically, for each SEEG channel we first computed the complex time-frequency map for the whole time series. Then, the resulting time-frequency map was whitened by Z_H0_-score normalization of each frequency bin. From the whole time-frequency map we extracted the time interval (200 ms) centered around each event of interest. The final time-frequency representation is obtained by computing the average of the time-frequency maps of power, i.e. incoherent averaging, corresponding to all the events of interest.

## 4. Precision and recall analysis

To quantify the capability of the subtypes of events identified by the NODE algorithm in segregating the SEEG channels involved in the epileptogenic zone (EZ) from those not involved (NIZ), we implemented a precision and recall analysis which is a suitable tool for imbalanced classification problems (in general Number NIZ channels >> Number of EZ channels). Once the clusters were computed using the NODE algorithm, we then implemented a standard precision and recall analysis. The precision and recall curve was computed as a function of a moving ER threshold for events pertaining to each NODE cluster of interest (CoI), and the area under the precision and recall curve (AUPREC) was computed to quantitatively summarize the capacity of EZ identification of each CoI. The chance level in each patient was computed as the ratio between the number of channels pertaining to EZ and the total number of channels. For the precision and recall analysis, the ground truth was given by the classification of the SEEG channels in the epileptogenic (EZ), propagation (PZ) and non-involved (NIZ) zones defined preoperatively by the epileptologists in all the analyzed patients (Engel I). This classification was made according to the sites of seizure initiation (SOZ) based on visual analysis and the Epileptogenicity Index (EI)^6-8^ (see Section 1 of this Appendix). The precision and recall analysis was also applied to assess the capacity of the subtypes of events identified by the NODE algorithm to map the resected zone (RZ) defined postoperatively by the epileptologists.

## 5. Characterization of the fast-ultradian dynamics

The fast-ultradian dynamics of the rate of the interictal events was analyzed with a fine-grained temporal resolution given by the time window of 200 ms used in the NODE algorithm. For this, we plotted the cumulative count of all the detected events (CE) as a function of the time for each SEEG channel (bipolar derivation). In these plots, the slope of each rectilinear segment correspond to the mean rate of events in that particular bipolar channel (see Figures 2A and 2B). In order to analyze the fluctuations of the events rate around its mean value, we subtracted a fitted straight line from the CE to obtain the residual of CE for each bipolar channel. The resulting detrended count of events (DCE) revealed in each SEEG channel the spontaneous fluctuations of the rate of interictal events over sub-hour time scales (see Figures 2C to 2F). In addition, time series of the event rate (ER) in each SEEG channel (bipolar derivation) and the AUPREC for EZ were constructed by computing the mean value of ER and the AUPREC for EZ value at each time position of a sliding epoch scanning the whole interictal SEEG recording available in each patient (see Figures 2G to 2J and Table 1). Different lengths of the sliding epoch were explored within the range 1 - 10 min. In each case, the length of the sliding epoch was kept unchanged to scan the whole interictal SEEG recording available in each patient. The overlap between successive time positions of the sliding epoch was 90% in all cases, with the exception of Figures 3E to 3H in which we use 1 min incremental step for all the epoch lengths. The resulting ER and AUPREC for EZ time series were z-score normalized before assessing their temporal correlation through the Pearson coefficient and linear regression (see Figure 4).

While the DCE consistently reproduce the slow fluctuations of the rate of events (ER) with a high temporal resolution (only limited by the 200 ms time window used in the NODE algorithm for detecting the events. See Figures 2C to 2F), it is essential to note that in order to compute the ER we must define a sliding time window in which the rate can be determined as ER = Number of events within the time window / Length of the time window. As a consequence, by using a time window of a finite length (e.g. 1 min, 5 min, 10 min) we are effectively imposing a constraint on the time resolution of the resulting ER time series. This is an important limitation, in particular for the patients disclosing rhythmic bursts of events with short time periods within the ultradian time scales (compare the Figure 2F with the Figure 2H).

## 6. Definition of the Spike-like group

To define the Spike-like cluster group we selected the first four NODE clusters capturing most of the IEDs as visually identified by an epileptologist (First four clusters in Figure S3: 09_09_09_09, 0_09_09_0, 09_09_09_0, 0_09_09_09). Then, for each one of these four NODE clusters we add the associated clusters corresponding to all the possible combinations of digits. For instance, for the cluster 0_09_09_0, we add the other three possible combinations of digits: 0_09_05_0, 0_05_09_0, 0_05_05_0. As a result, the Spike-like group includes the epileptiform discharges pertaining to the clusters 09_09_09_05, 0_09_09_05 having a high power for EZ localization (see Figure 1), and also events from other 34 clusters, some of them being less specific to the EZ like the cluster 09_09_09_0 shown in Figure S1.

## 7. Spontaneous fast-ultradian dynamics of the rate of interictal events

We investigated the dependence of the observed fluctuations of the AUPREC for EZ on the type of events and on the specificity of the detector of interictal discharges. For the same time resolution than that used in Figures 2I and 2J (sliding epoch of 5 min in length and 90% overlap), the Figures 3A and 3B show the temporal dynamics of AUPREC for EZ for three type of events, 1) epileptiform discharges pertaining to a single cluster 0_09_09_05 (see Figure 1C and 1F), 2) Spike-like events resulting from the combination of 36 clusters (see Figure S1C) and 3) Events pertaining to the cluster 09_09_0_0, associated with amplitude outliers in the low frequency bands (1 Hz - 30 Hz). The Spike-like category can be thought as the result of a detector of interictal discharges with low specificity when compared to the manual marking produced by an epileptologist. For a detailed definition of the Spike-like group the reader is referred to Section 6 of this Appendix. Figures 2I, 2J, 3A and 3B show that the events pertaining to the clusters 09_09_09_05, 0_09_09_05 and the Spike-like group produce similar temporal dynamics of the AUPREC for EZ. These results reveal that the temporal fluctuations of the events rate during interictal periods are present across different subtypes of epileptiform discharges and effectively entrain the precision to localize the EZ. Moreover, we quantified the magnitude of the excursions of the AUPREC for EZ time series observed during the interictal periods by computing the relative difference (RD) between the extreme AUPREC for EZ values with respect to its maximum value ((max - min) / max) for three sliding epoch lengths. Figures 3C and 3D show that the RD of AUPREC for EZ decreases with the epoch length (for the epileptiform discharges 09_09_09_05, the median of the RD of AUPREC for EZ across all the patients is 0.67 (IQR = 0.43 - 0.75), 0.43 (IQR = 0.34 - 0.65) and 0.27 (IQR = 0.20 - 0.47) for epoch lengths of 1 min, 5 min and 10 min, respectively). However, we found no significant difference between the RD of AUPREC for EZ values at the population level for epileptiform discharges (Clusters: 09_09_09_05, 0_09_09_05) and the Spike-like group (green and red violin plots shown in Figures 3C and 3D). This result suggests that the magnitude of the temporal fluctuations of the AUPREC for EZ computed using IEDs (e.g. Clusters: 09_09_09_05, 0_09_09_05) do not significantly diminishes when other non-epileptiform events besides the IEDs are included in the analysis (e.g. Spike-like group).

## 8. Attenuation of the interictal fluctuations as a function of the epoch length

In order to gain insight about the nature of the temporal dynamics of different types of interictal events, we then characterize the magnitude of the excursions of the AUPREC for EZ as a function of the sliding epoch length. It is worth noting that the fluctuations of AUPREC for EZ are mainly produced by the relative change of the event rate in EZ with respect to the rest of the SEEG channels (PZ and NIZ). Figures 3A and 3B show that, as expected, non-epileptic events (blue dots) produce very low values of AUPREC for EZ when compared to the epileptiform discharges (red dots), however, fluctuations of the AUPREC for EZ values are observed in both types of events. Figures 3E and 3F show the absolute difference (AD) between the extreme values (max - min) of the AUPREC for EZ time series based on the rate of IEDs, as a function of the sliding epoch length. These log-linear plots show a linear trend suggesting an exponential dependence *y ∝ e^-x/Tao^* where *y* represents the AD of AUPREC for EZ being proportional to a decaying exponential function of the epoch length *x*, with a characteristic time scale *Tao* in the range 11.7 min - 12.7 min (CoIs: 0_09_09_05 and 09_09_09_05). On the other hand, Figures 3G and 3H show the dependence of the AD of AUPREC for EZ values based on the rate of non-epileptic events, as a function of the sliding epoch length. In this case, we found a piecewise linear trend characterized by two linear segments with different slope in a log-log plot, suggesting a scale-free behavior. Interestingly, the AD of AUPREC for EZ values based on the Spike-like group showed a dependence with the sliding epoch length in between the exponential and scale-free behavior (data not shown). As a conclusion, these results strongly suggest that the temporal fluctuations of the AUPREC for EZ values based on the rate of IEDs is characterized by a scale-rich process showing an exponentially decay as a function of the epoch length with a characteristic time scale *Tao*. This scale-rich behavior is essentially different from the temporal dynamics of the AD of AUPREC for EZ based on the non-epileptic interictal events showing a scale-free trend (see Figures 3G and 3H and Figure S2). Figures 3E and 3F show the absolute difference (AD) between the extreme values (max - min) of the AUPREC for EZ time series based on the rate of IEDs, as a function of the sliding epoch length. These log-linear plots show a linear trend suggesting an exponential dependence *y ∝ e^-x/Tao^* where *y* represents the AD of AUPREC for EZ being proportional to a decaying exponential function of the epoch length *x*, with a characteristic time scale *Tao* in the range 11.7 min - 12.7 min (CoIs: 0_09_09_05 and 09_09_09_05). Let us consider the relative attenuation *A* obtained for two epoch lengths *x_1_*and *x_2_*, *A* = (*y1 - y2*) / *y1*

By substituting in this equation the exponential dependences *y1* ∝ *e^-x1/Tao^*and *y2* ∝ *e^-x2/Tao^*and operating algebraically we obtain the following expression, *x2 = x1 - Tao Ln*(1 - *A*) where *Ln*() stands for natural logarithm. Thus, considering a characteristic time constant of *Tao* = 12.2 min and *x1* = 5 min, the required epoch length *x2* for a 90% attenuation of *y2* with respect to *y1* results *x2* = 5 min - 12.2 min *Ln*(1 - 0.9) = 33.1 min.

## 9. Putative mechanisms linked to the fast-ultradian dynamics of the rate of interictal events

The temporal fluctuations of the IEDs rate and its potential to confound EZ localization have been reported in the context of circadian and multidien time scales in relation to wakefulness, sleep, seizure occurrence, post-ictal state and antiepileptic drug withdrawal.^15-19^ Besides, fluctuations of the spike rate over sub-hour time scales have been reported during attention and memory tasks.^20-23^ Also, hour-to-hour changes in the spatial distribution of the epileptic spikes, explained in part by sleep and in part by seizures, have been previously reported.^24^ In contrast, the temporal dynamics observed in our study appear to emerge spontaneously in the interictal SEEG traces of the 35 patients with Engel I seizure outcome included in the analysis, that is, not directly related to any of the previously reported mechanisms mentioned above. It is important to note that the results shown in the Figures 4A and 4B assess the propagation mechanism involving the occurrence of the same type of IEDs (CoI: 09_09_09_05) in EZ and PZ, which could effectively confound the identification of the EZ channels and have an impact on the dynamics of AUPREC for EZ. On the other hand, there is evidence showing that sharp IEDs in EZ in general propagate through the network changing their waveform shape, emerging in PZ as smoother spikes,^25,26^ see also Figure 2A in Tomlinson et al.^27^ In our analysis, the Spike-like group could capture this effect since it is constituted by a variety of IEDs subtypes (see Figure S1C), including sharp spikes (e.g. 09_09_09_05) and smoother waveform shapes (e.g. 09_09_09_0). However, Figure 4C shows that even for the Spike-like group the propagation mechanism could have a dominant role in explaining the interictal fluctuations of the AUPREC for EZ time series in a limited fraction of the analyzed patients (upper bound of approx. 20%). This result constitutes additional evidence supporting the hypothesis of importance of the local intrinsic excitability of the epileptogenic tissue in connection with fast-ultradian time scales.

## 10. Predicting the spontaneous fast-ultradian dynamics to improve the epileptogenic zone localization

We found that the time series corresponding to 1) the mean rate of events including all the clusters and averaged across all the channels (ER of all Clust in all Chan) and 2) the mean rate of epileptiform spikes (ER of CoI in all Chan, with CoI: 0_09_09_05 or 09_09_09_05) both negatively correlates with the AUPREC for EZ time series computed using these IEDs subtypes (e.g. clusters 0_09_09_05 and 09_09_09_05 associated by the epileptologists with interictal epileptiform spikes and spike-wave complexes respectively). Note that the overall rate of interictal events "ER of all Clust in all Chan" and the rate of epileptiform spikes "ER of CoI in all Chan" across all the SEEG channels, are suitable measures for prospective analysis since they do not depend on the SEEG channels classification in EZ, PZ, NIZ. Importantly, the AUPREC for EZ time series computed for these IEDs subtypes produced a negative correlation with the "ER of all Clust in all Chan" time series of higher magnitude than that observed with the "ER of CoI in all Chan" time series for CoI: 0_09_09_05 or 09_09_09_05 (data not shown). In the case of the cluster 09_09_09_05, Figures 5B and 5C show that the correlation between the time series AUPREC for EZ and "ER of all Clust in all Chan", is negative and statistically significant in 63% (22/35) of the patients. Of note, the slope of the linear regression between AUPREC for EZ and "ER of all Clust in all Chan" time series in the case of the IEDs subtype 09_09_09_05 is −0.334 (see Figures 5C and S8) which is the double of the negative slope obtained using the cluster 0_09_09_05 (see Figure S8). As a consequence, the overall rate of interictal events (epileptic and non-epileptic) across all the SEEG channels can better predict the interictal dynamics of the spike-wave complexes (CoI: 09_09_09_05) with respect to the epileptiform spikes (CoI: 0_09_09_05). Figure S7 illustrates the temporal correlation among the time series AUPREC for EZ, "ER of IEDs (CoI: 09_09_09_05) in EZ" and "ER of all Clust in all Chan" observed in four patients. This feature can be understood by considering three facts, 1) in most patients, the NIZ channels are more numerous than the EZ channels, 2) the temporal dynamics of the ER corresponding to a) the interictal events in NIZ and b) the IEDs in EZ present a significant degree of dissociation (see Figures 2E and 2F), 3) IEDs occur predominantly in EZ, and are also observed in a minor proportion in PZ and NIZ. The latter represents a confounding factor for EZ localization. Thus, a decrease of "ER of all Clust in all Chan" will produce a reduction of the ER of all the interictal events, including the IEDs, in NIZ (most numerous channels). This decrease will to some extent be dissociated from the dynamics of the ER of IEDs in EZ, which in turn will likely result in an increase of the AUPREC for EZ value.

Importantly, the magnitude of the negative correlation between the overall rate of interictal events (epileptic and non-epileptic) across all the SEEG channels and the precision to localize the EZ varies across the subtypes of IEDs used to compute the AUPREC for EZ. Figure S8 shows the slope of the linear regression between AUPREC for EZ and "ER of all Clust in all Chan" time series for all the NODE clusters. Figure S9 shows the predictive performance for near-optimal EZ localization of all the NODE clusters. In Figure S9, the AUPREC for EZ values were computed in each patient from the 5 min epoch associated with the minimum value of "ER of all Clust in all Chan" (i.e. estimated best 5 min epoch for near-optimal EZ localization). Of note, while both IEDs subtypes corresponding to the clusters 09_09_09_05 (spike-wave complexes) and 0_09_09_05 (epileptiform spikes) have been found to be more abundant in EZ than in NIZ (see panels C and D in Figure S1) and both produce good EZ localization on average across the 35 patients (see panel A in Figure S9), only the cluster 09_09_09_05 (spike-wave complexes) disclose a strong negative correlation between the AUPREC for EZ and "ER of all Clust in all Chan" time series resulting in a statistically significant predictive value in estimating the best 5 min epoch for near-optimal EZ localization (compare the clusters 09_09_09_05 and 0_09_09_05 in Figures S1, S8 and S9). These conclusions also hold true for the RZ localization (see Figures S10 and S11).

## 11. Comparing the fast-ultradian dynamics between awake and non-REM sleep states

In this section we discuss the results related to the comparison between two states of the patients: 1) awake at rest and 2) non-REM sleep (see Methods and Table S1). The comparison was made by quantifying the magnitude of the fast-ultradian fluctuations of the precision to localize EZ based on the rate of interictal events. For this, time series of the event rate (ER) in each SEEG channel (bipolar derivation) and the AUPREC for EZ quantifying the goodness of EZ localization based on the rate of interictal events were constructed by computing the mean value of ER and the AUPREC for EZ value at each time position of a sliding epoch scanning the whole interictal SEEG recording (see Sections 4 and 5 of this Appendix). In this case, we used a sliding epoch of 5 min in length and 90% overlap to scan the whole SEEG time series (bipolar derivations) available in each patient (see Table S1). Figures S13 to S16 show for 4 patients the resulting ER and AUPREC for EZ time series together with the DCE (see Sections 5 of this Appendix) for the awake and non-REM sleep states and computed using the interictal events pertaining to the cluster 09_09_09_05. Figure S17 summarizes the results for the IEDs (clusters 09_09_09_05, 0_09_09_05) and non-epileptic events (cluster 09_0_0_0) across the 27 patients included in this analysis (see Table S1). These 27 patients are a subset of the main group of 35 patients listed in Table 1 and S1 and they were selected according to the following two criteria: A) SEEG recordings were available for both awake at rest and non-REM sleep states and B) in each patient the difference in the time length of the SEEG recordings between the two states were not greater than 100%. The median time-length and range of the interictal SEEG traces among the subset of 27 patients for the awake and non-REM sleep states were 28.5 min (range = 25.8 - 31.5 min) and 28.2 min (range = 25.2 - 32.5 min), respectively. Figure S17 shows the relative difference (RD) between the extreme values ((max - min) / max) of the AUPREC for EZ time series as a measure quantifying the magnitude of the fast-ultradian fluctuations of the precision to localize EZ based on the rate of interictal events. Importantly, we found no significant differences at the group level (N=27) between the awake at rest and non-REM sleep states in terms of the RD of AUPREC for EZ for the IEDs and non-epileptic events shown in Figure S17 (P = 1, Wilcoxon signed rank test with the P values Bonferroni-adjusted to correct for multiple comparisons across the 80 NODE clusters). Figure S18 summarizes the comparison between awake at rest and non-REM sleep states in terms of the RD of AUPREC for EZ for the 80 NODE clusters. The number of days between the two SEEG recordings sessions included in the analysis has a median of 1 days (range = 0 - 3 days). Taken together, these results suggest that the temporal fluctuations of the rate of different subtypes of interictal events entraining the precision to localize the EZ over fast-ultradian time scales occurs spontaneously during both the awake and the non-REM sleep states of the analyzed patients. Importantly, we found that the magnitude of the temporal fluctuations of the AUPREC for EZ presents no significant differences between the awake at rest and non-REM sleep states of the analyzed patients.

**Table S1:** Days between the two recording sessions and time of day (or night) when the SEEG recordings were made for the 35 patients reported in Table 1 of the main text. To ensure the anonymity, serial numbers attributed randomly to each patient are used as patients ID. The resulting ID numbers have no correlation with any clinical information of the patients.

| Patient ID | Awake SEEG recordings<br>(Time [hh:mm],<br>Recording length [min]) | Asleep SEEG recordings<br>(Time [hh:mm],<br>Recording length [min]) | Number of days between the<br>SEEG recording sessions |
| --- | --- | --- | --- |
| 1 | 19:04, 22.9 | 03:36, 20.8 | 3 |
| 2 | 08:52, 27.5 | N/A | N/A |
| 3 | 18:47, 30.3 | 02:17, 32 | 4 |
| 4 | 12:37, 27.3 | 01:47, 25 | 1 |
| 5 | 11:22, 25.4 | 01:23, 27.5 | 1 |
| 6 | 11:17, 14.7 | N/A | N/A |
| 7 | 15:27, 60 | N/A | N/A |
| 8 | 14:35, 26 | N/A | N/A |
| 9 | 08:51, 60 | N/A | N/A |
| 10 | 10:47, 20.4 | N/A | N/A |
| 11 | 10:03, 60 | 02:07, 60 | 1 |
| 12 | 10:07, 27.8 | 03:50, 28.2 | 1 |
| 13 | 15:09, 30.6 | N/A | N/A |
| 14 | 15:05, 11.6 | 03:22, 22.6 | 3 |
| 15 | 14:42, 29 | 01:40, 27.7 | 0 |
| 16 | 21:37, 14 | 23:52, 22.6 | 0 |
| 17 | 16:04, 43.4 | 02:09, 60 | 1 |
| 18 | 17:53, 31.9 | 02:32, 25.8 | 1 |
| 19 | 10:20, 34.5 | 02:10, 26.2 | 0 |
| 20 | 10:20, 16.3 | 04:32, 32.3 | 0 |
| 21 | 09:40, 34.7 | 00:21, 30.8 | 4 |
| 22 | 17:45, 28.8 | 02:00, 20 | 1 |
| 23 | 09:57, 22.5 | 01:15, 29.7 | 0 |
| 24 | 14:09, 30.2 | 06:15, 60 | 6 |
| 25 | 18:42, 28.7 | 02:22, 29.4 | 0 |
| 26 | 16:07, 15.5 | 02:30, 25 | 0 |
| 27 | 16:35, 28.2 | 01:55, 32.3 | 1 |
| 28 | 11:52, 12.5 | N/A | N/A |
| 29 | 09:55, 27.3 | 03:50, 19.2 | 1 |
| 30 | 10:37, 28.4 | 02:3, 25.7 | 4 |
| 31 | 11:20, 27.3 | 02:20, 26 | 0 |
| 32 | 07:09, 30.4 | 02:11, 34 | 0 |
| 33 | 09:19, 60 | 00:16, 60 | 0 |
| 34 | 12:48, 26.8 | 20:19, 32.5 | 4 |
| 35 | 15:46, 60 | 01:01, 60 | 4 |

Symbols and abbreviations: N/A, Not available.

## 12. Comparing the fast-ultradian dynamics between two SEEG recording sessions in awake state

In this section we present the results related to the comparison between two SEEG recording sessions, referred as Awake 1 and Awake 2, taken at different days and time of day for the same patient state (awake at rest, see Tables S2 and S3). The comparison was made by quantifying the magnitude of the fast-ultradian fluctuations of the precision to localize EZ based on the rate of interictal events. For this, time series of the event rate (ER) in each SEEG channel (bipolar derivation) and the AUPREC for EZ quantifying the goodness of EZ localization based on the rate of interictal events were constructed by computing the mean value of ER and the AUPREC for EZ value at each time position of a sliding epoch scanning the whole interictal SEEG recording (see Sections 4 and 5 of this Appendix). In this case, we used a sliding epoch of 5 min in length and 90% overlap to scan the whole SEEG time series (bipolar derivations) available in each patient (see Table S3). Figures S19 to S22 show for 4 patients the resulting ER and AUPREC for EZ time series together with the DCE (see Sections 5 of this Appendix) for the Awake 1 and Awake 2 states and computed using the interictal events pertaining to the cluster 09_09_09_05. Figure S23 summarizes the results for the IEDs (clusters 09_09_09_05, 0_09_09_05) and non-epileptic events (cluster 09_0_0_0) across the 12 patients included in this analysis (see Tables S2 and S3). In each patient the difference in the time length of the SEEG recordings between the two states were not greater than 100%. The median time-length and range of the interictal SEEG traces among the subset of 12 patients for the Awake 1 and Awake 2 states were 30.1 min (range = 27.5 - 31.4 min) and 31.4 min (range = 28.1 - 33.4 min), respectively. Figure S23 shows the relative difference (RD) between the extreme values ((max - min) / max) of the AUPREC for EZ time series as a measure quantifying the magnitude of the fast-ultradian fluctuations of the precision to localize EZ based on the rate of interictal events. Importantly, we found no significant differences at the group level (N=12) between the Awake 1 and Awake 2 states in terms of the RD of AUPREC for EZ for the IEDs and non-epileptic events shown in Figure S23 (P = 1, Wilcoxon signed rank test with the P values Bonferroni-adjusted to correct for multiple comparisons across the 80 NODE clusters). Figure S24 summarizes the comparison between Awake 1 and Awake 2 states in terms of the RD of AUPREC for EZ for the 80 NODE clusters. The number of days between the two SEEG recordings sessions included in the analysis has a median of 5.5 days (range = 4 - 7 days). Taken together, these results suggest that the magnitude of the temporal fluctuations of the AUPREC for EZ assessed for the same patient state (awake at rest) over fast-ultradian time scales, presents no significant differences across several days.

**Table S2:**
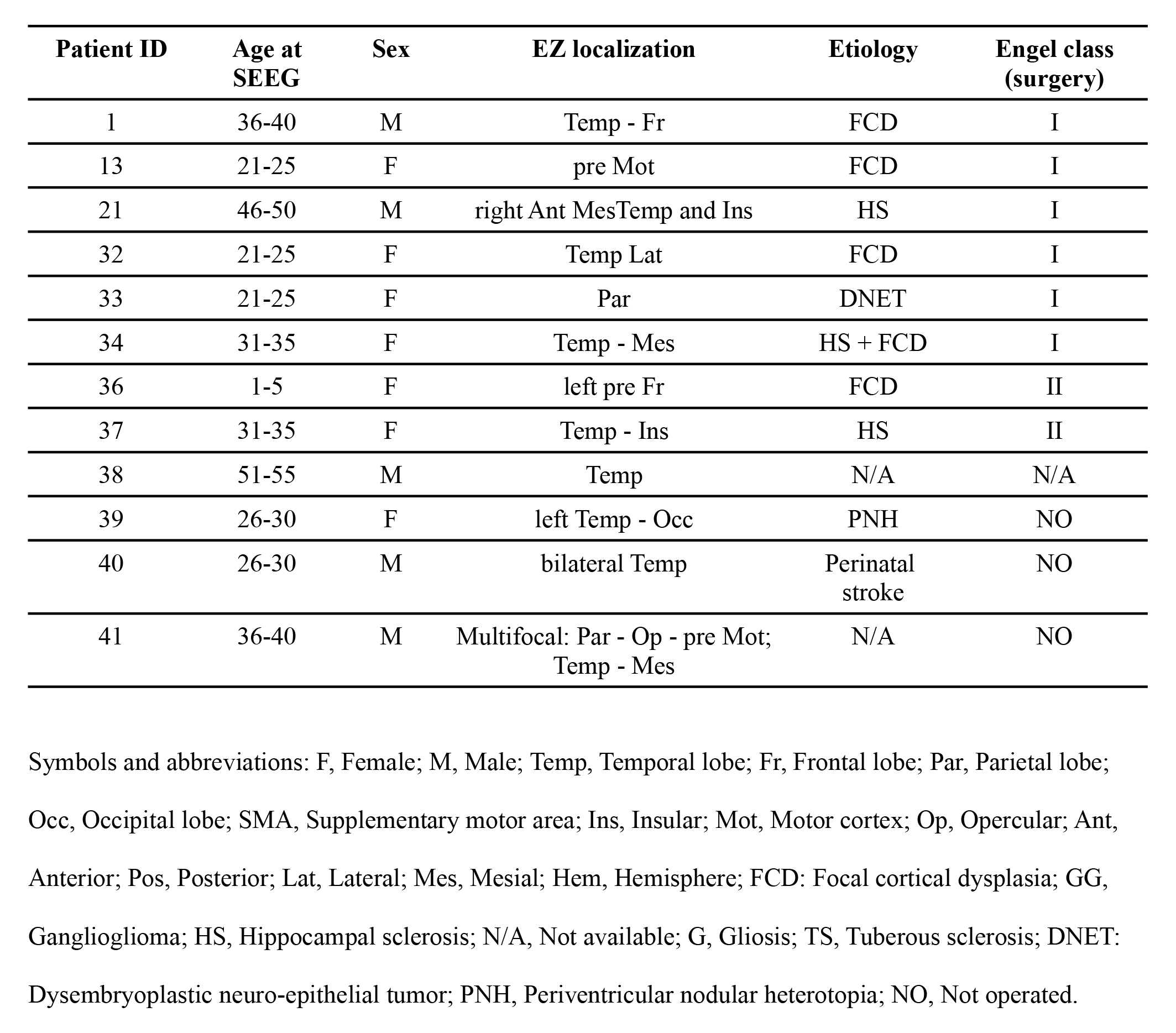
Patients clinical information. The length of the interictal SEEG recordings corresponding to these patients can be found in the Table S3. To ensure the anonymity, serial numbers attributed randomly to each patient are used as patients ID. The resulting ID numbers have no correlation with any clinical information of the patients.

**Table S3:** Days between the two recording sessions and time of day when the SEEG recordings were made for the 12 patients reported in Table S2. To ensure the anonymity, serial numbers attributed randomly to each patient are used as patients ID. The resulting ID numbers have no correlation with any clinical information of the patients.

| Patient ID | Awake 1 SEEG recordings<br>(Time [hh:mm],<br>Recording length [min]) | Awake 2 SEEG recordings<br>(Time [hh:mm],<br>Recording length [min]) | Number of days between the<br>SEEG recording sessions |
| --- | --- | --- | --- |
| 1 | 19:04, 22.9 | 13:57, 27.5 | 9 |
| 13 | 15:09, 30.6 | 10:26, 33.5 | 6 |
| 21 | 09:40, 34.7 | 08:55, 31.7 | 4 |
| 32 | 07:09, 30.4 | 09:09, 30.7 | 7 |
| 33 | 09:19, 60 | 13:04, 60 | 1 |
| 34 | 12:48, 26.8 | 08:30, 32 | 5 |
| 36 | 17:10, 32.2 | 10:19, 35.3 | 2 |
| 37 | 21:26, 28.8 | 08:00, 31 | 4 |
| 38 | 09:31, 19.3 | 06:44, 11.3 | 6 |
| 39 | 17:47, 28.8 | 08:57, 27.1 | 10 |
| 40 | 16:30, 29.8 | 12:20, 28.7 | 4 |
| 41 | 16:16, 30.7 | 09:36, 33.4 | 7 |

---

**Figure S1:**
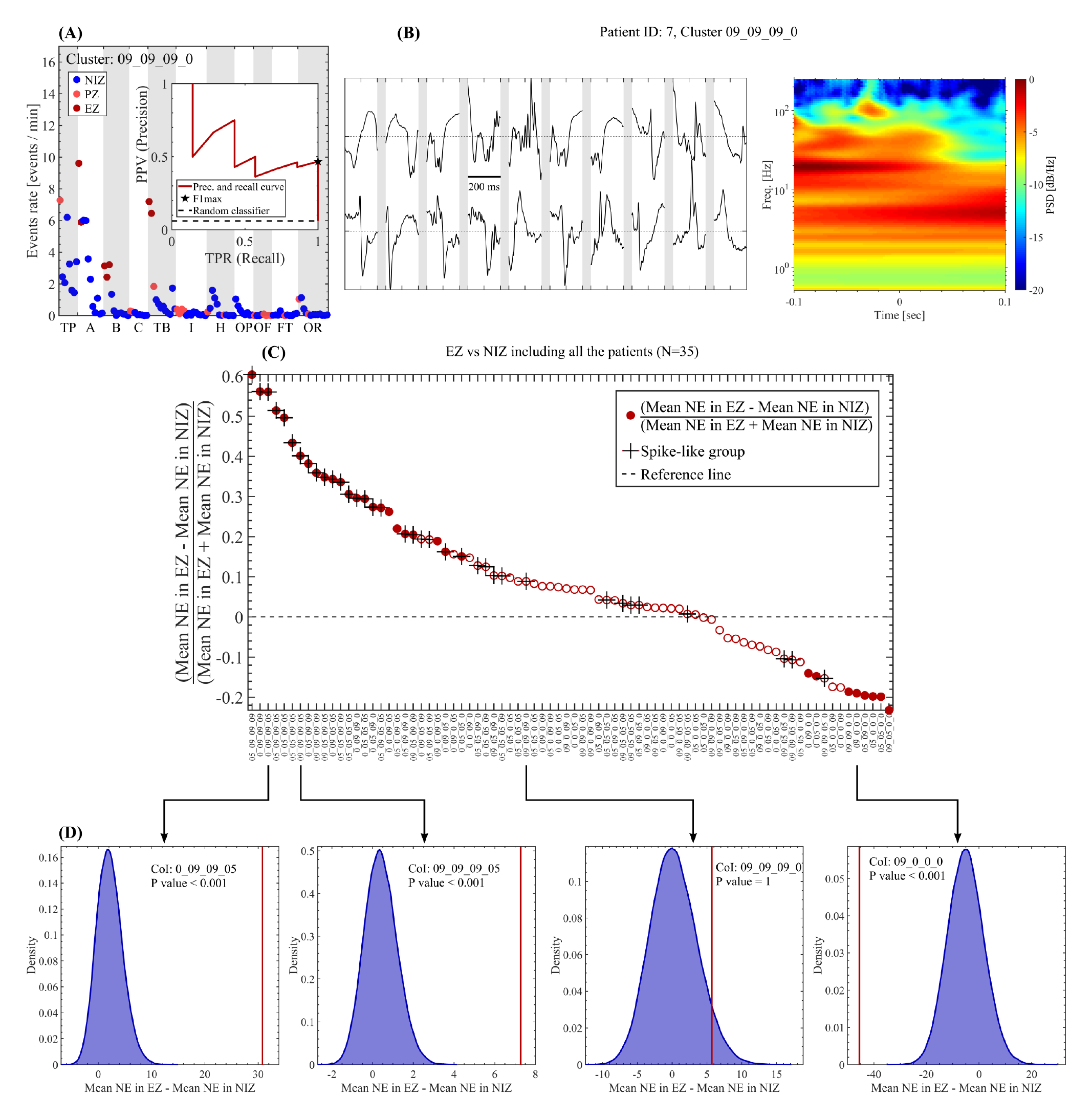
Characterization of the NODE clusters. **(A)** Mean rate of events for the CoI: 09_09_09_0 in each bipolar channel obtained from the whole interictal SEEG recording available for the patient 7 (60 min). The inset shows the results of the precision and recall analysis for the sharp events pertaining to the clusters 09_09_09_0 (F1 max = 0.64). **(B)** Raw time series and whitened time-frequency map (scalogram using Morlet wavelets) for the events pertaining to the clusters 09_09_09_0 detected in the EZ (B: Hippocampus cephalis) of the patient 7. **(C)** Relative difference between the mean values of the two distributions: 1) NE in each channel pertaining to the EZ and 2) NE in each channel pertaining to the NIZ. First, the channels were segregated in the EZ and NIZ groups across all the patients included in Table 1 (N=35). Then, the NE of a given CoI was evaluated in each channel using the complete time series length available in each patient. Finally, the relative difference between the mean values of NE computed as (Mean NE in EZ - Mean NE in NIZ) / (Mean NE in EZ + Mean NE in NIZ) is shown as a function of the clusters. Filled and empty circles indicate clusters producing significant (Bonferroni-adjusted P value < 0.05) and non-significant (Bonferroni-adjusted P value ≥ 0.05) difference between the mean NE values of the EZ and NIZ distributions, respectively (non-parametric permutation test). The cross markers highlight the clusters included in the Spike-like group. **(D)** Histograms showing the difference between the mean NE values of the surrogate EZ and NIZ distributions including all the patients of Table 1 (N=35). The histograms were computed via random sampling without replacement (10^5^ permutations). The red vertical solid line shown in the histograms indicates the difference between the mean NE values of the actual EZ and NIZ distributions (Mean NE in EZ - Mean NE in NIZ). The reported P values resulting from the non-parametric permutation test were Bonferroni-adjusted to correct for multiple comparisons across the 80 clusters. Symbols and abbreviations: PSD, power spectral density; EZ, epileptogenic zone; PZ, propagation zone; NIZ, non-involved zone; PPV, positive predictive value; TPR, true positive rate; AUPREC, area under the precision and recall curve; NE, Number of Events; TP, Temporal pole; A, Amygdala complex; B, Hippocampus cephalis; C, Hippocampus caudalis; TB, Temporal basalis; I, Insula; H, Gyrus temporal transverse Heschl; OP, Opercule parietal; OF, Opercule frontal; FT, Front Triangularis; OR, Fronto Orbitaire oblique.

**Figure S2:**
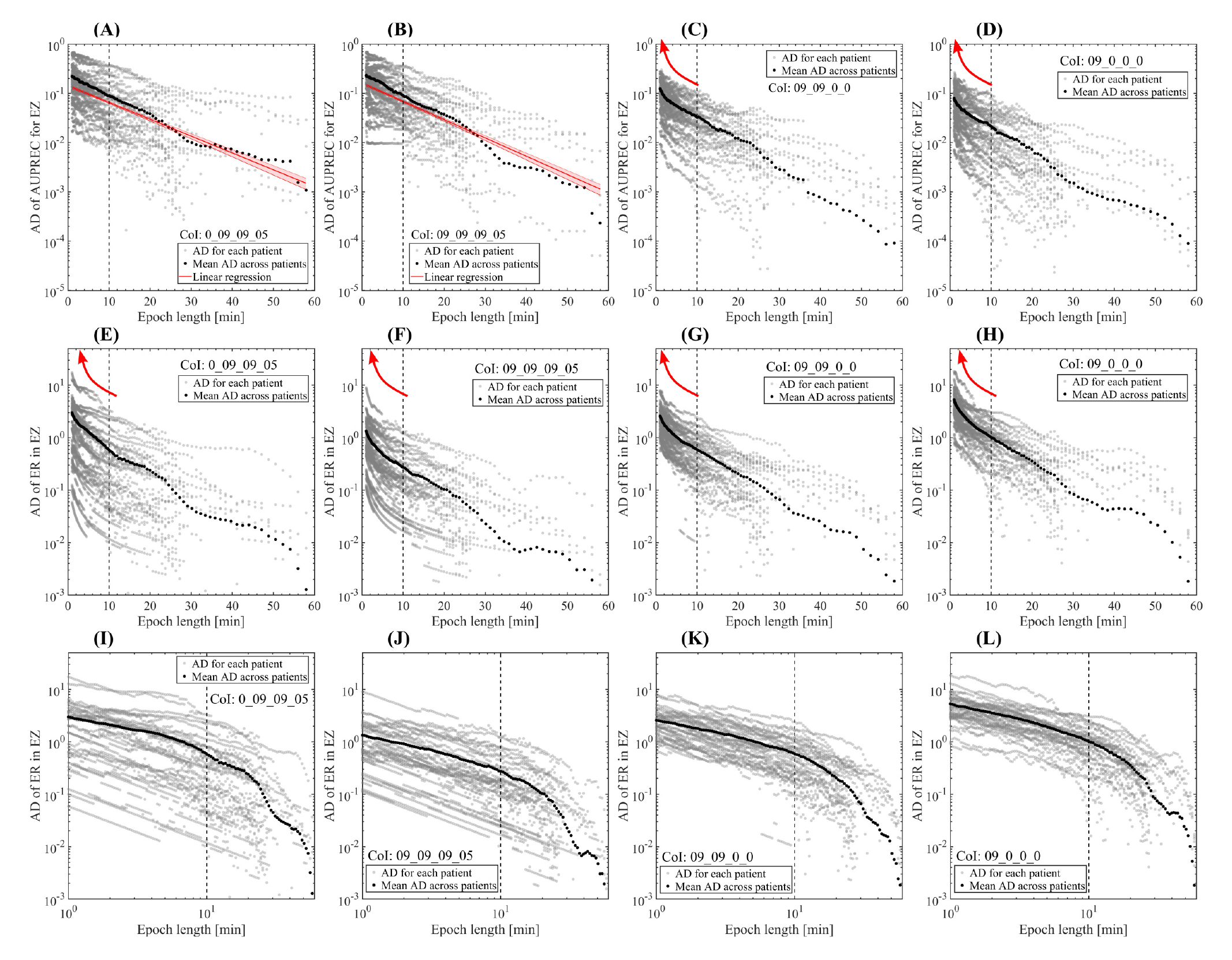
Plots showing the absolute difference (AD) between the extreme values (max - min) of the ER in EZ and AUPREC for EZ as a function of the sliding epoch length. **(A - D)** Log-linear plots showing the absolute difference (AD) between the extreme values (max - min) of the AUPREC for EZ as a function of the sliding epoch length. **(E - H)** Log-linear plots showing the absolute difference (AD) between the extreme values (max - min) of the ER in EZ as a function of the sliding epoch length. **(I - J)** Log-log plots showing the absolute difference (AD) between the extreme values (max - min) of the ER in EZ as a function of the sliding epoch length. In all the panels, the gray dots associated with the AUPREC for EZ and ER in EZ measures correspond to the mean value in each position of the sliding epoch for the interictal events pertaining to the cluster of interest (CoI). 1 min incremental step for all the epoch lengths was used to scan the whole time series available in each patient. In the case of IEDs (clusters 0_09_09_05 and 09_09_09_05), panels A and B show a linear trend of the AD of AUPREC for EZ values indicating an exponential dependence, with a characteristic time scale *Tao* (linear trend in a log-linear plot), disclosed by the fluctuations of the AUPREC for EZ as a function of the epoch length (see also Figures 3E and 3F). This scale-rich behavior is essentially different from the dependence of the AD of AUPREC for EZ values based on the non-epileptic interictal events (clusters 09_09_0_0 and 09_0_0_0) shown in panels C and D (see red arrows), which were found to disclose a scale-free trend (i.e. linear trend in a log-log plot, see Figures 3G and 3H). Regarding the fluctuations of the ER in EZ measure shown in panels E to L, it was found a scale-free like dependence of the AD of ER in EZ as a function of the epoch length for both epileptic and non-epileptic events (see the linear trend in the log-log plots corresponding to panels I to L). Taken together, these results suggest that in the case of IEDs (e.g. clusters 0_09_09_05 and 09_09_09_05), the observed ultradian fluctuations of the AUPREC for EZ of can not be completely explained by considering it simply as a function of the IED rate fluctuations within the EZ. Symbols and abbreviations: CoI, cluster of interest; EZ, epileptogenic zone; AD, absolute difference; ER, events rate, IEDs, interictal epileptogenic discharges; AUPREC, area under the precision and recall curve.

**Figure S3:**
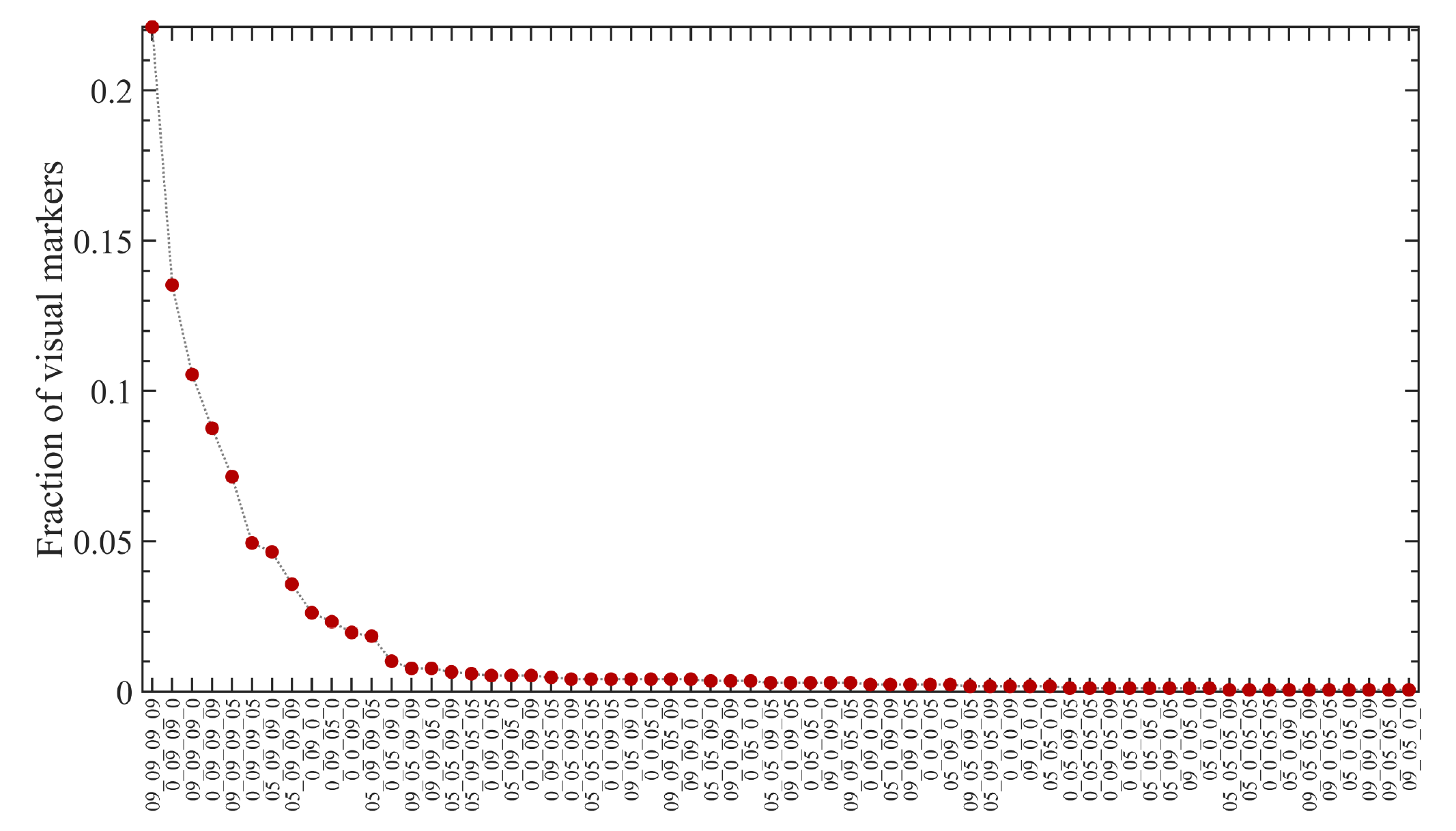
Fraction of the visual markers captured by the NODE clusters. The 1678 epileptiform discharges included in this plot were visually identified by an epileptologist (F. Bonini) from the SEEG traces of three patients included in the Table 1: 4, 6 and 7. The analysis included a 5 min epoch in each patient and a total of 12 bipolar SEEG channels (2 NIZ, 2 PZ and 8 EZ). The NODE algorithm detected the 99% of the events corresponding to the visual markers and segregated them in the 64 clusters shown in the abscissa axis. Most of the visually marked IEDs were grouped into the first NODE clusters shown on the left side of the plot. These NODE clusters were found to produce significant difference between the mean NE values of the EZ and NIZ distributions (see Figure S1). Symbols and abbreviations: EZ, epileptogenic zone; PZ, propagation zone; NIZ, non-involved zone; NE, Number of Events; IEDs, interictal epileptogenic discharges; NODE, Nested Outlier Detection.

**Figure S4:**
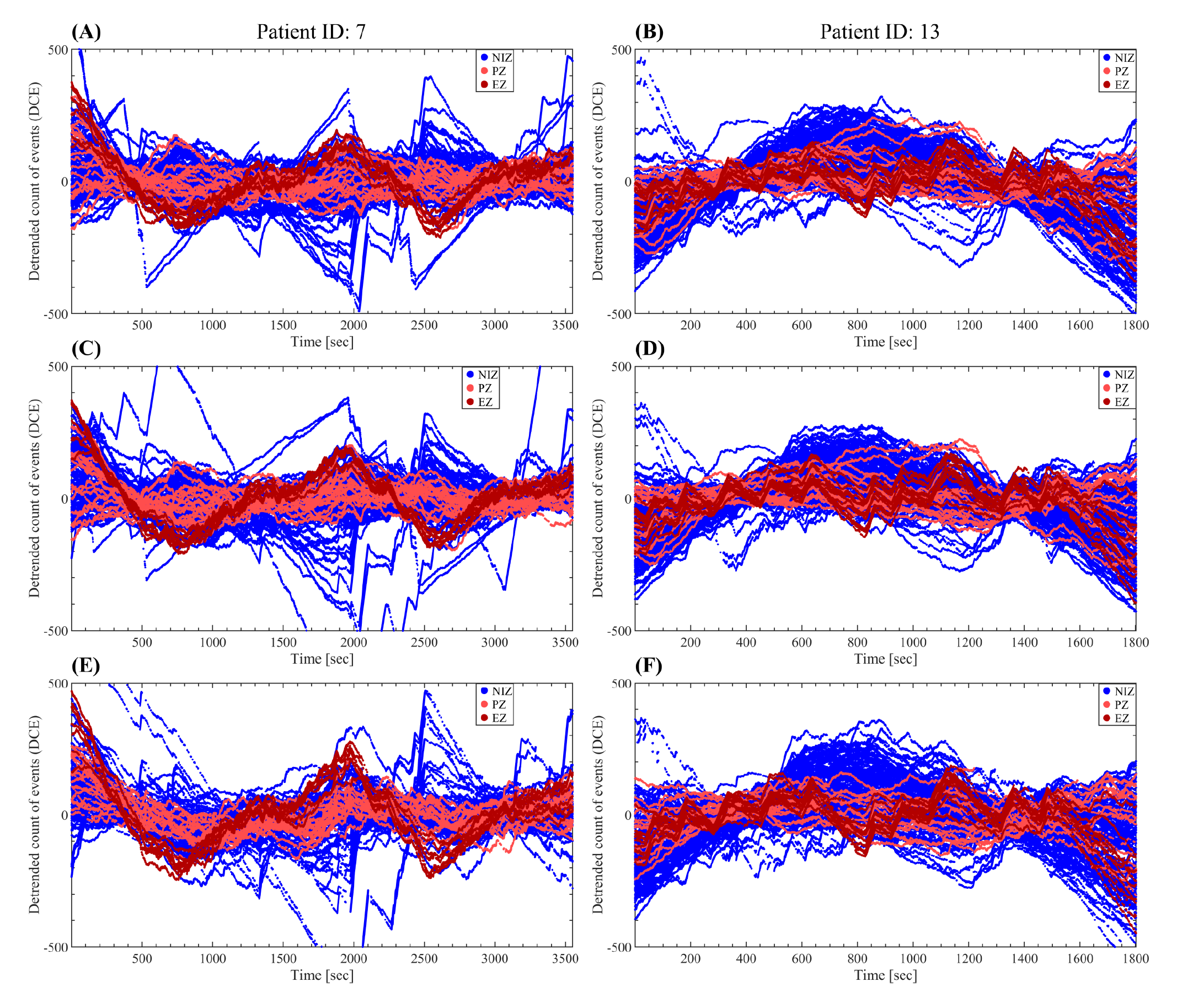
The spontaneous interictal dynamics of the events rate is highly independent of the frequency bands used in the NODE algorithm. The parameters of the NODE algorithm were configured as: 200 milliseconds time window length associated with each event and LFDR thresholds = {0.5, 0.1}. **(A, B)** 4 Frequency bands: [1 Hz - 10 Hz], [8 Hz - 32 Hz], [30 Hz - 155 Hz], [150 Hz - 255 Hz]. **(C, D)** 7 Frequency bands: [0.1 Hz - 4.9 Hz], [4 Hz - 9 Hz], [8 Hz - 14 Hz], [13 Hz - 31 Hz], [30 Hz - 85 Hz], [80 Hz - 155 Hz], [150 Hz - 255 Hz]. **(E, F)** 2 Frequency bands: [0.1 Hz - 60.9 Hz], [60 Hz - 255 Hz]. Symbols and abbreviations: NODE, Nested Outlier Detection; EZ, epileptogenic zone; PZ, propagation zone; NIZ, non-involved zone; LFDR, local false discovery rate.

**Figure S5:**
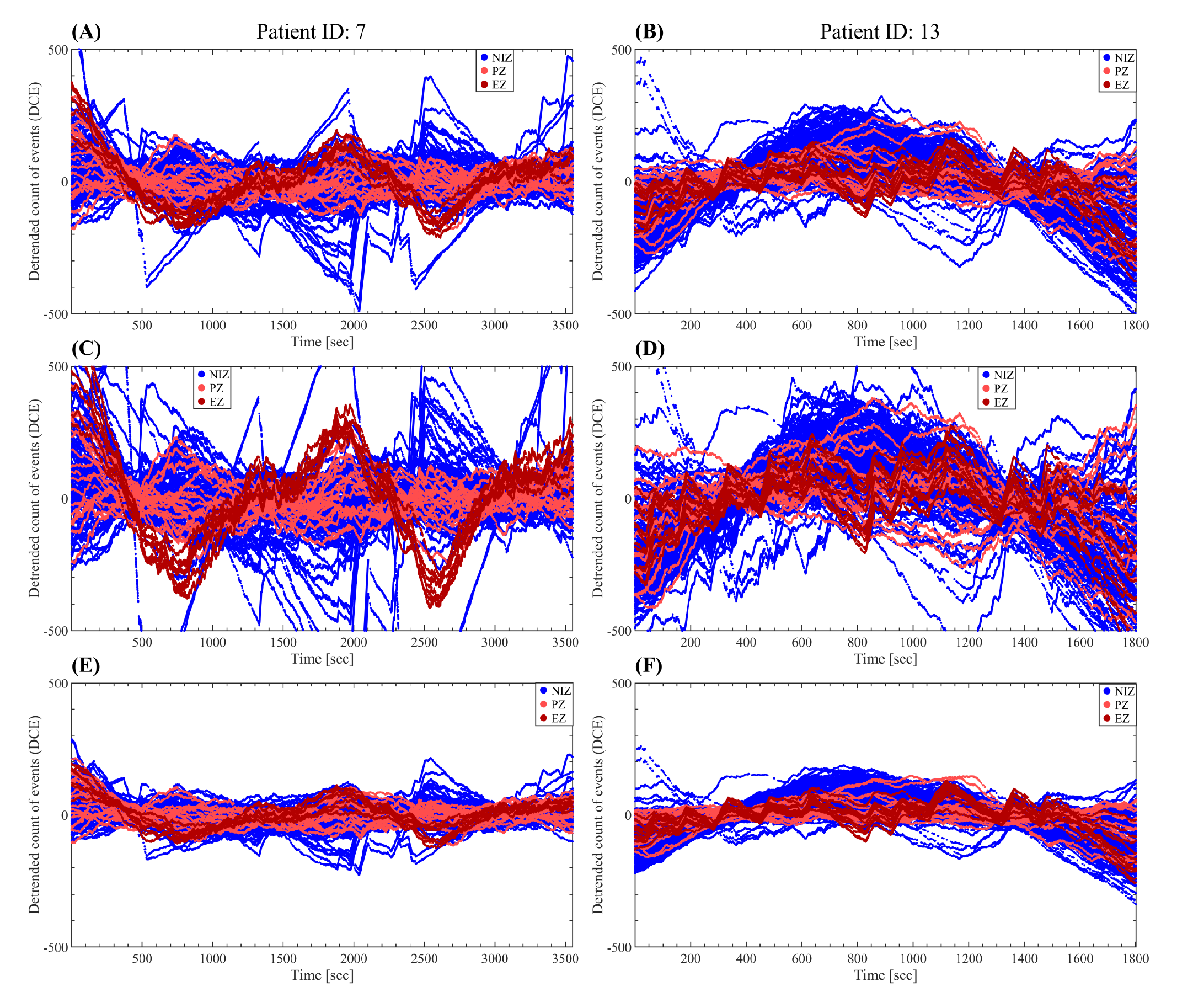
The spontaneous interictal dynamics of the events rate is highly independent of the time window length used to define the events. The parameters of the NODE algorithm were configured as: LFDR thresholds = {0.5, 0.1}, 4 frequency bands: [1 Hz - 10 Hz], [8 Hz - 32 Hz], [30 Hz - 155 Hz], [150 Hz - 255 Hz]. **(A, B)** 200 milliseconds time window length associated with each event. **(C, D)** 100 milliseconds time window length associated with each event. **(E, F)** 400 milliseconds time window length associated with each event. Symbols and abbreviations: NODE, Nested Outlier Detection; EZ, epileptogenic zone; PZ, propagation zone; NIZ, non-involved zone; LFDR, local false discovery rate.

**Figure S6:**
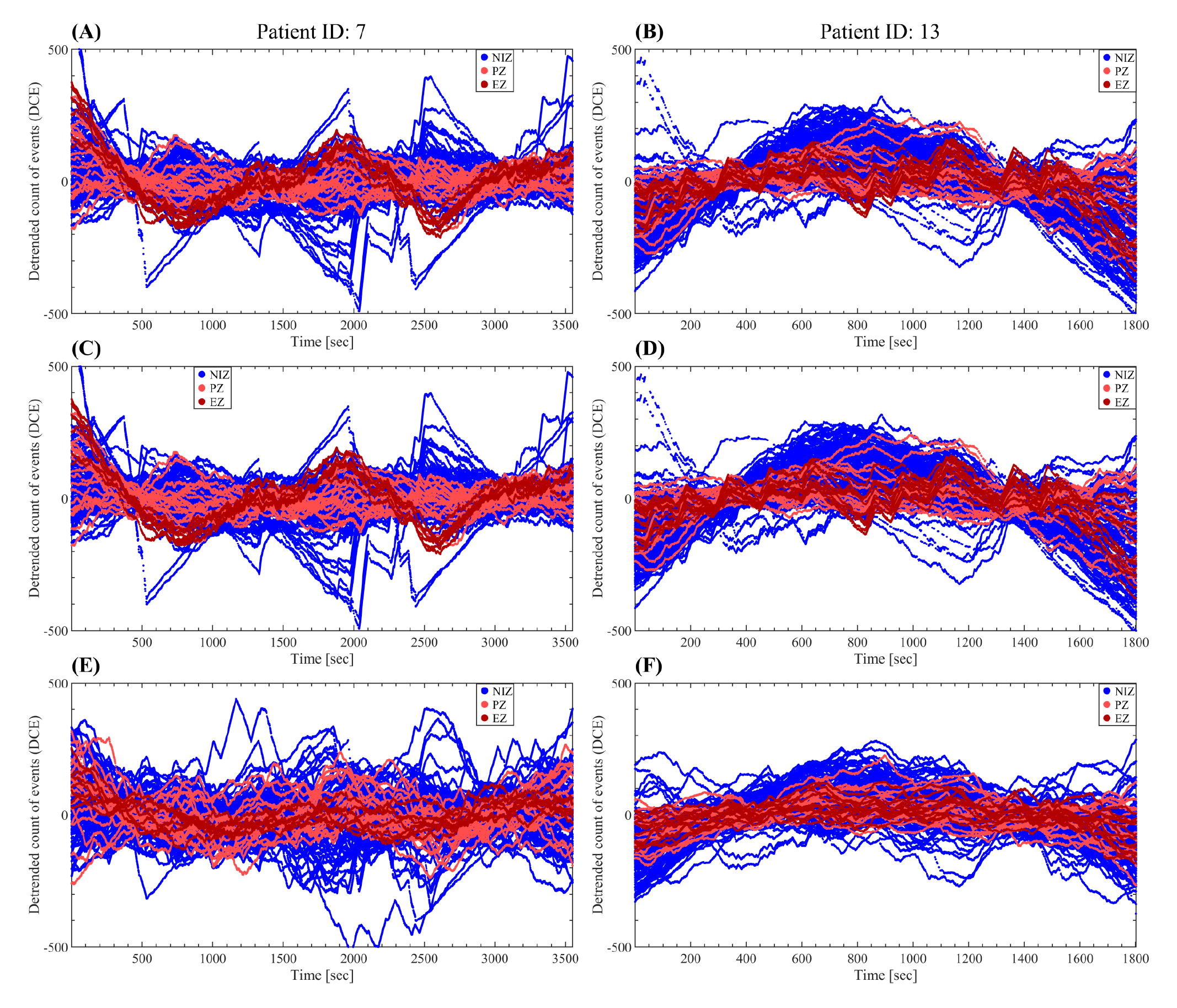
The spontaneous interictal dynamics of the events rate is attenuated as more noise is allowed by increasing the higher LFDR threshold used in the NODE algorithm. Note that the rhythmic behavior of the events rate dynamics becomes almost indistinguishable from the random fluctuations only in the case where a very high number of noisy anomalies are allowed, i.e. very high values of the higher LFDR threshold (local false discovery rate > 90%, see panels E and F corresponding to LFDR thresholds = {higher threshold = 0.9>>0.5, lower threshold = 0.1}). The parameters of the NODE algorithm were configured as: 200 milliseconds time window length associated with each event, 4 frequency bands: [1 Hz - 10 Hz], [8 Hz - 32 Hz], [30 Hz - 155 Hz], [150 Hz - 255 Hz]. **(A, B)** LFDR thresholds = {0.5, 0.1} **(C, D)** LFDR thresholds = {0.5, 0.05}. **(E, F)** LFDR thresholds = {0.9, 0.1}. Symbols and abbreviations: NODE, Nested Outlier Detection; EZ, epileptogenic zone; PZ, propagation zone; NIZ, non-involved zone; LFDR, local false discovery rate.

**Figure S7:**
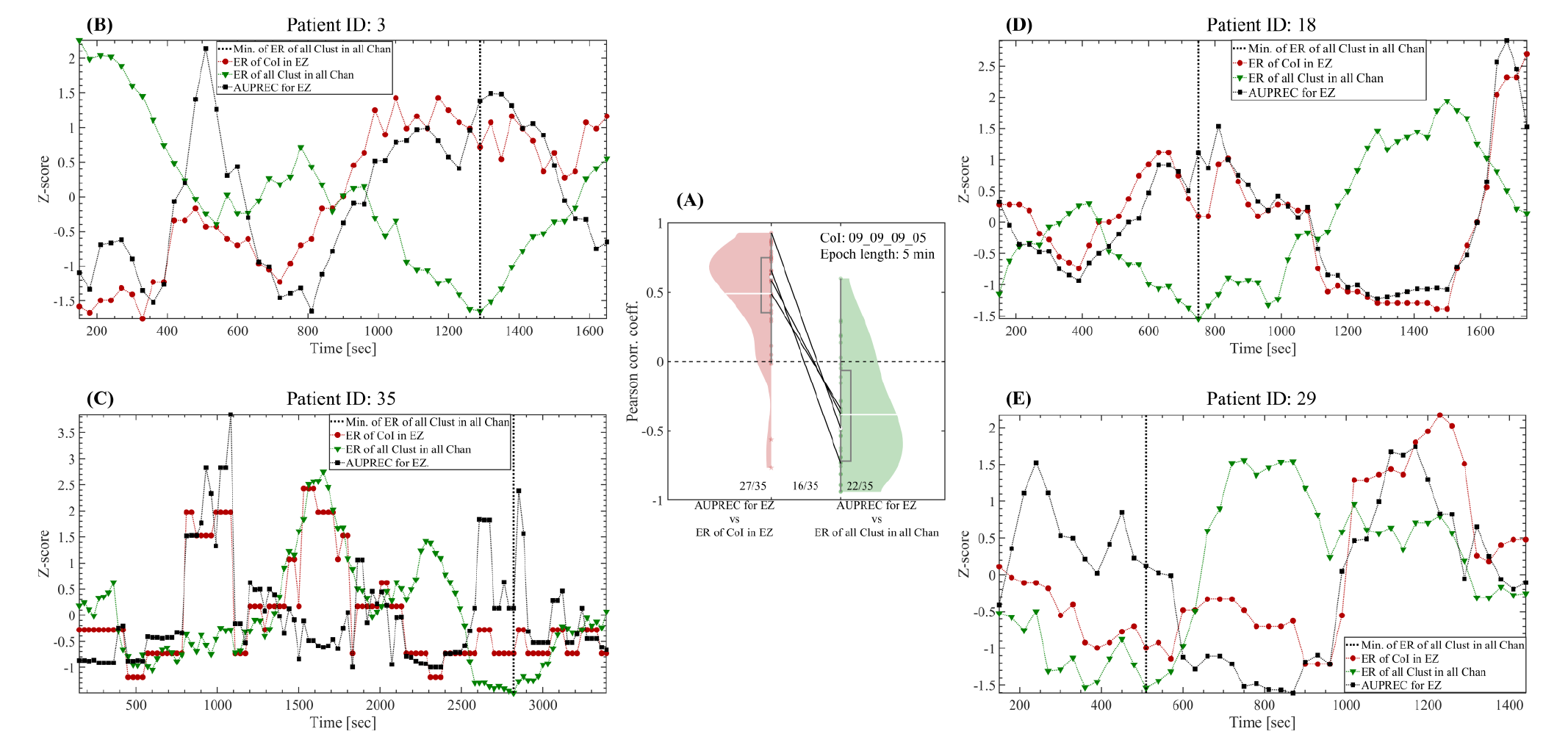
Estimating the best interictal 5 min-epoch for near-optimal EZ localization. In the time series AUPREC for EZ, ER of CoI for EZ and ER of all Clust in all Chan, each point corresponds to the mean value of these quantities in a sliding epoch of 5 min in length and 90% overlap. **(A)** Violin plots showing all the patients paired across the two correlations, 1) AUPREC for EZ vs ER of CoI in EZ, and 2) AUPREC for EZ vs ER of all Clust in all Chan. Solid black lines correspond to the four patients 3, 18, 29 and 35, in which the two correlations are significant and have opposite sign. The three fractional numbers accompanying the paired violin plots indicate the fraction of patients presenting significant correlations values. The statistical significance of the correlations (P < 0.05) was assessed by using the Student’s t distributions of the two-tailed hypothesis test under the null hypothesis that the correlation is zero. **(B, C, D, E)** Time series corresponding to the four patients indicated by the solid black lines y panel **(A)**. Note the out-of-phase oscillations of the negative correlated time series AUPREC for EZ and ER of all Clust in all Chan. In each patient, the best interictal 5 min-epoch for near-optimal EZ localization (maximum of the AUPREC for EZ time series) occurs approximately at the minimum of the overall rate of the interictal epileptic and non-epileptic events (see the vertical dotted line marking the minimum of the ER of all Clust in all Chan time series).

**Figure S8:**
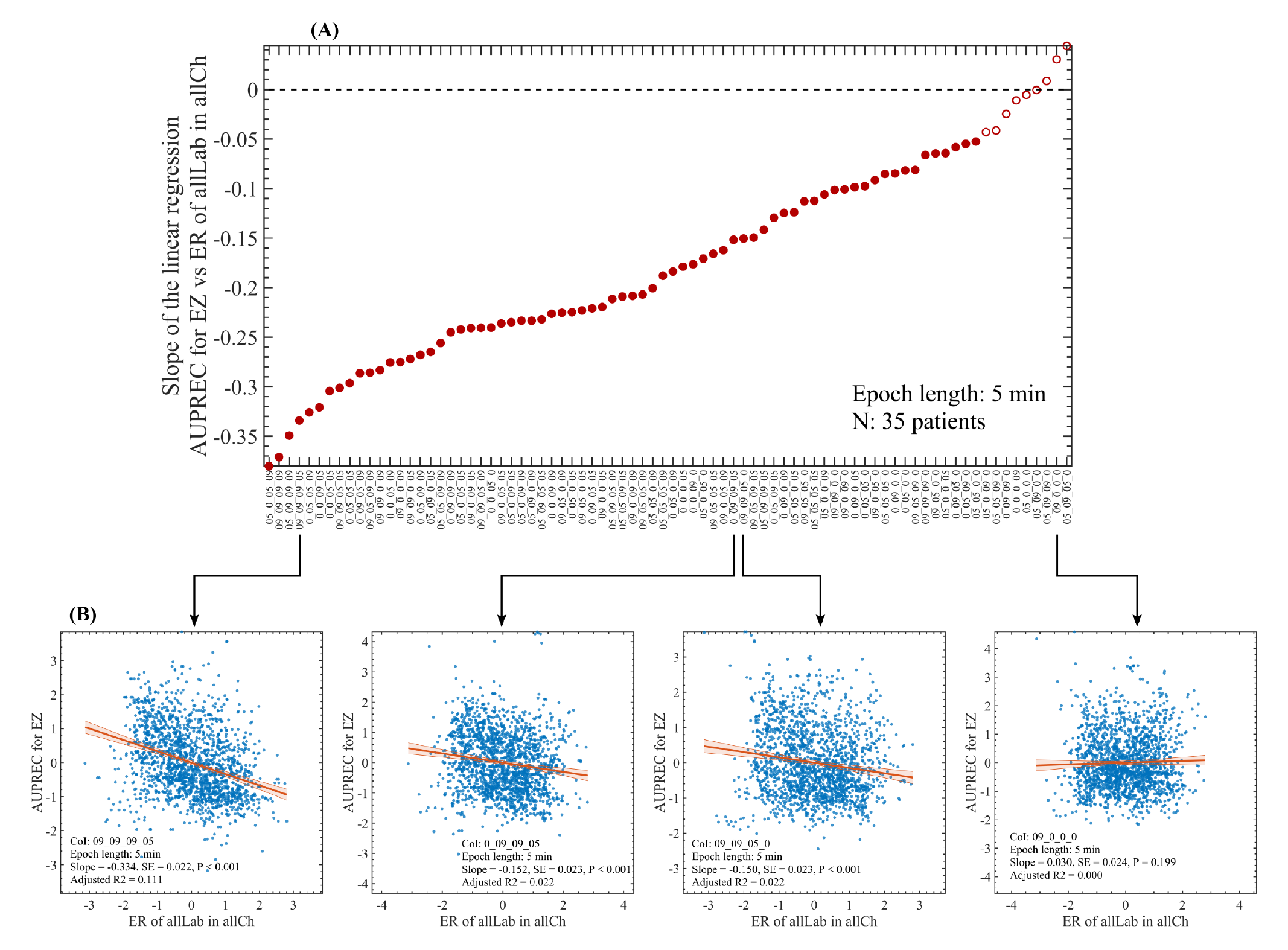
Correlation between the time series AUPREC for EZ and ER of all Clust in all Chan. **(A)** Slope of the linear regression between the AUPREC for EZ and ER of all Clust in all Chan values as a function of the NODE clusters. The two measures were computed for the CoI: 09_09_09_05 and each dot correspond to the measure value in a particular time position of the sliding epoch of 5 min in length and 90% overlap, covering the whole interictal SEEG time series available in each patient. The red line and red shaded error bars represent the linear regression and the 95% confidence interval, respectively. **(B)** Scatter plots showing the correlation between the time series AUPREC for EZ and ER of all Clust in all Chan for 4 NODE clusters. All the reported P values correspond to the t-statistic of the two-sided hypothesis test (no Bonferroni correction was implemented). Symbols and abbreviations: CoI, cluster of interest; EZ, epileptogenic zone; AUPREC, area under the precision and recall curve. CL, chance level; ER of all Clust in all Chan, mean rate of events including all the clusters and averaged across all the channels.

**Figure S9:**
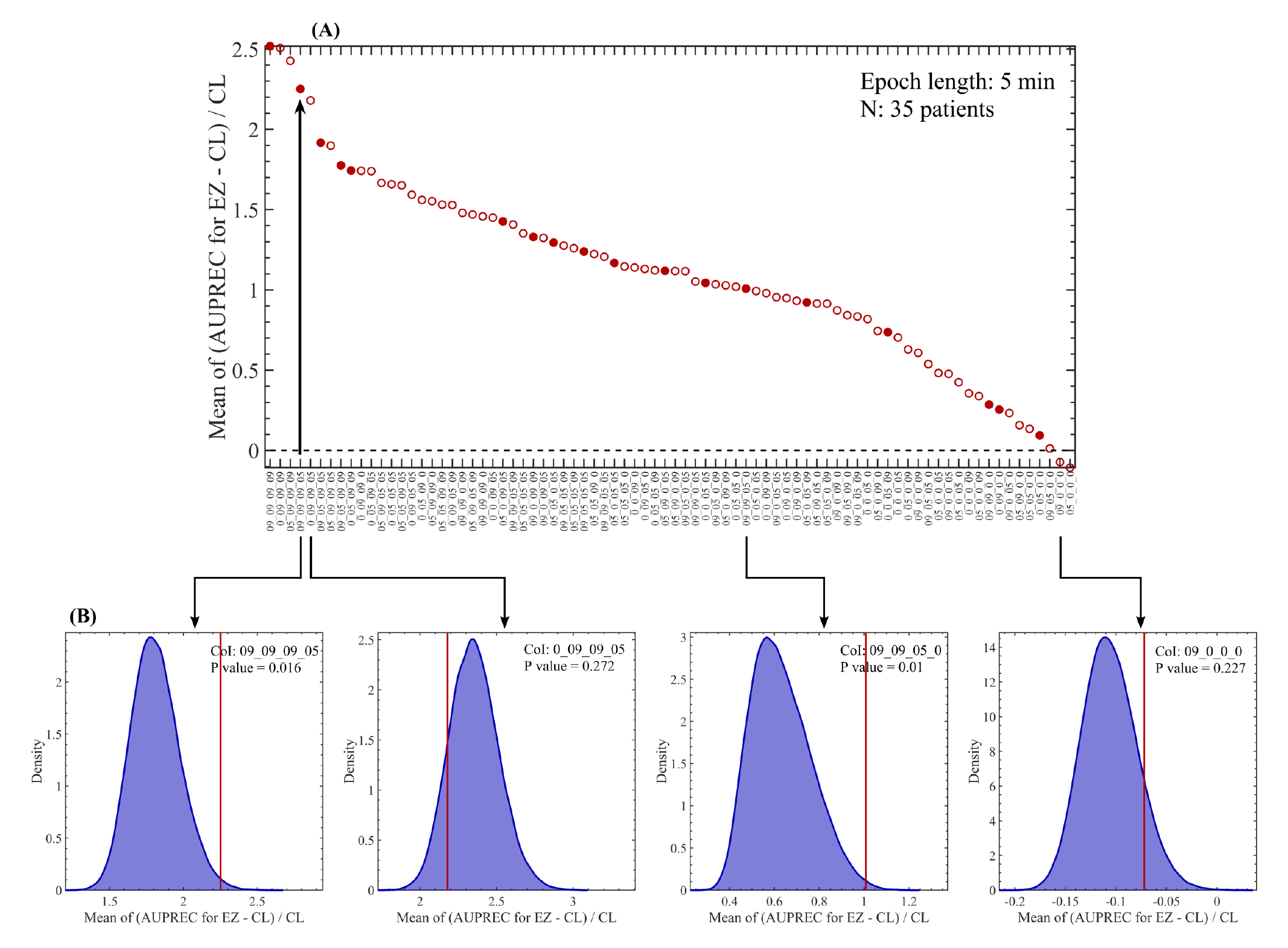
Predictive performance of the NODE clusters. **(A)** Mean relative difference of the AUPREC for EZ with respect to the chance level (CL) as a function of the NODE clusters. First, the AUPREC for EZ value was computed in each patient from the 5 min epoch associated with the minimum value of ER of all Clust in all Chan (i.e. estimated best 5 min epoch for near-optimal EZ localization). Then, the relative difference of the AUPREC for EZ with respect to the chance level (CL) was computed. In each patient, the CL was obtained as the ratio between the number of channels pertaining to EZ and the total number of channels. Finally, the relative difference was averaged across the 35 patients listed in Table 1. Filled and empty circles indicate clusters producing significant (P value < 0.05) and non-significant (P value ≥ 0.05) difference between the estimated best 5 min epoch for near-optimal EZ localization and the distribution of values produced by 5 min epochs randomly sampled from the interictal SEEG recordings of each patient (10^5^ random samplings). **(B)** Histograms showing the distribution of values produced by 5 min epochs randomly sampled from the interictal SEEG recordings of each patient. The histograms were computed via random sampling without replacement (10^5^ permutations). The red vertical solid line shown in the histograms indicates the mean relative difference produced by the best 5 min epoch for near-optimal EZ localization. The reported P values were obtained from the non-parametric permutation test (no Bonferroni correction was implemented). Symbols and abbreviations: CoI, cluster of interest; EZ, epileptogenic zone; AUPREC, area under the precision and recall curve. CL, chance level; ER of all Clust in all Chan, mean rate of events including all the clusters and averaged across all the channels.

**Figure S10:**
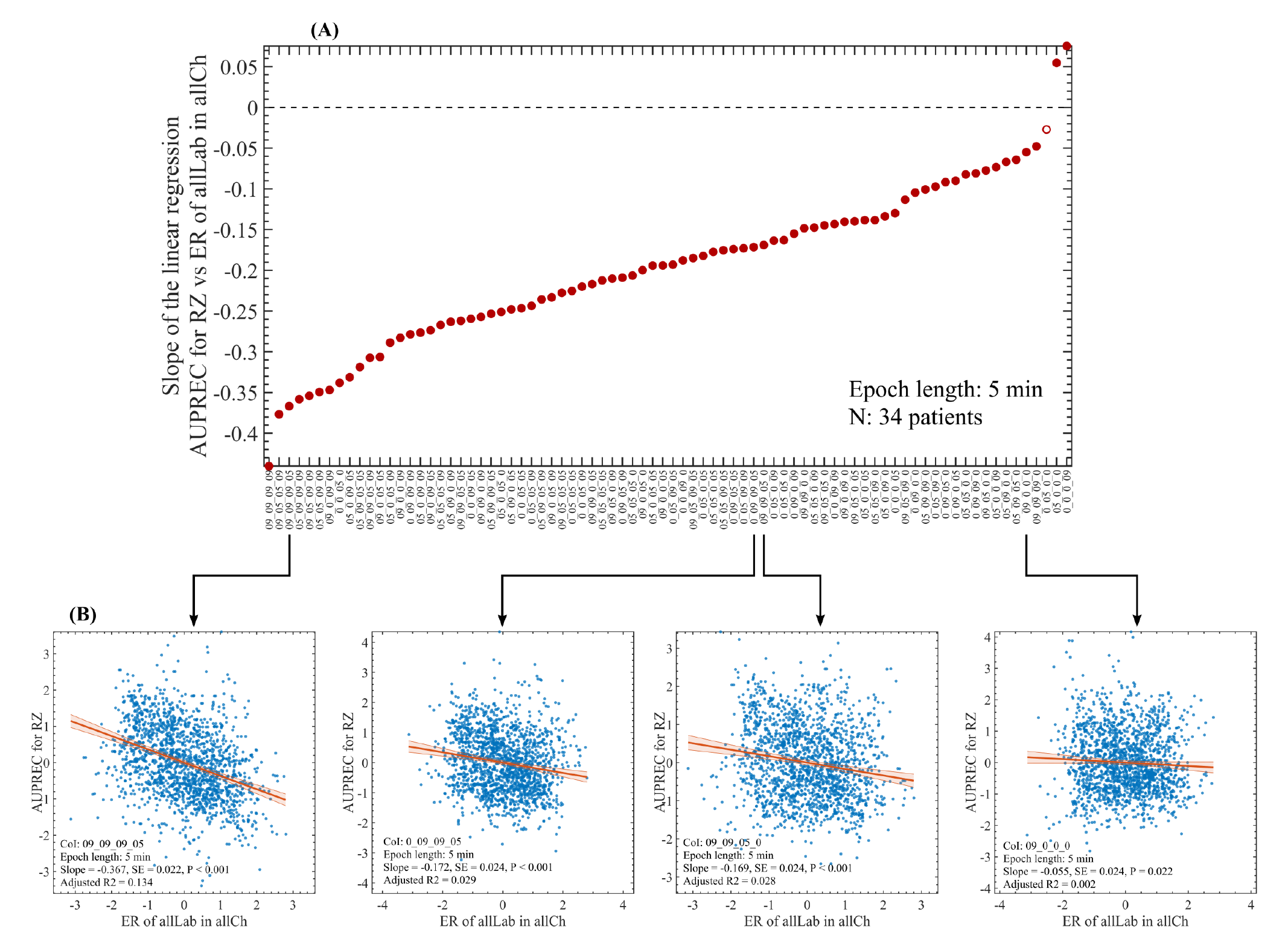
Correlation between the time series AUPREC for RZ and ER of all Clust in all Chan. **(A)** Slope of the linear regression between the AUPREC for RZ and ER of all Clust in all Chan values as a function of the NODE clusters. The two measures were computed for the CoI: 09_09_09_05 and each dot correspond to the measure value in a particular time position of the sliding epoch of 5 min in length and 90% overlap, covering the whole interictal SEEG time series available in each patient. The red line and red shaded error bars represent the linear regression and the 95% confidence interval, respectively. **(B)** Scatter plots showing the correlation between the time series AUPREC for RZ and ER of all Clust in all Chan for 4 NODE clusters. All the reported P values correspond to the t-statistic of the two-sided hypothesis test (no Bonferroni correction was implemented). Symbols and abbreviations: CoI, cluster of interest; RZ, resected zone; AUPREC, area under the precision and recall curve. CL, chance level; ER of all Clust in all Chan, mean rate of events including all the clusters and averaged across all the channels.

**Figure S11:**
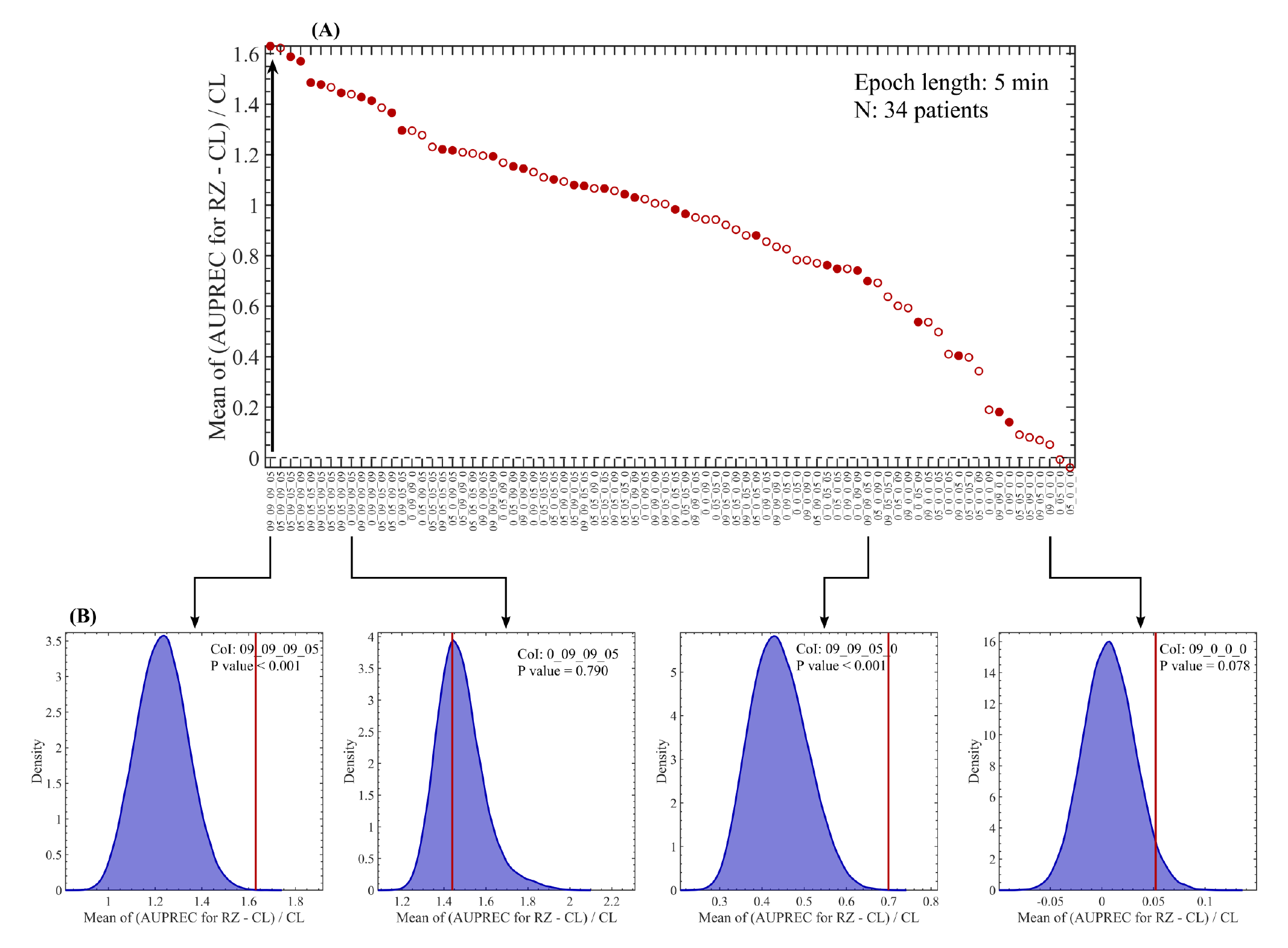
Predictive performance of the NODE clusters. **(A)** Mean relative difference of the AUPREC for RZ with respect to the chance level (CL) as a function of the NODE clusters. First, the AUPREC for RZ value was computed in each patient from the 5 min epoch associated with the minimum value of ER of all Clust in all Chan (i.e. estimated best 5 min epoch for near-optimal RZ localization). Then, the relative difference of the AUPREC for RZ with respect to the chance level (CL) was computed. In each patient, the CL was obtained as the ratio between the number of channels pertaining to RZ and the total number of channels. Finally, the relative difference was averaged across the 35 patients listed in Table 1. Filled and empty circles indicate clusters producing significant (P value < 0.05) and non-significant (P value ≥ 0.05) difference between the estimated best 5 min epoch for near-optimal RZ localization and the distribution of values produced by 5 min epochs randomly sampled from the interictal SEEG recordings of each patient (10^5^ random samplings). **(B)** Histograms showing the distribution of values produced by 5 min epochs randomly sampled from the interictal SEEG recordings of each patient. The histograms were computed via random sampling without replacement (10^5^ permutations). The red vertical solid line shown in the histograms indicates the mean relative difference produced by the best 5 min epoch for near-optimal RZ localization. The reported P values were obtained from the non-parametric permutation test (no Bonferroni correction was implemented). Symbols and abbreviations: CoI, cluster of interest; RZ, resected zone; AUPREC, area under the precision and recall curve. CL, chance level; ER of all Clust in all Chan, mean rate of events including all the clusters and averaged across all the channels.

**Figure S12:**
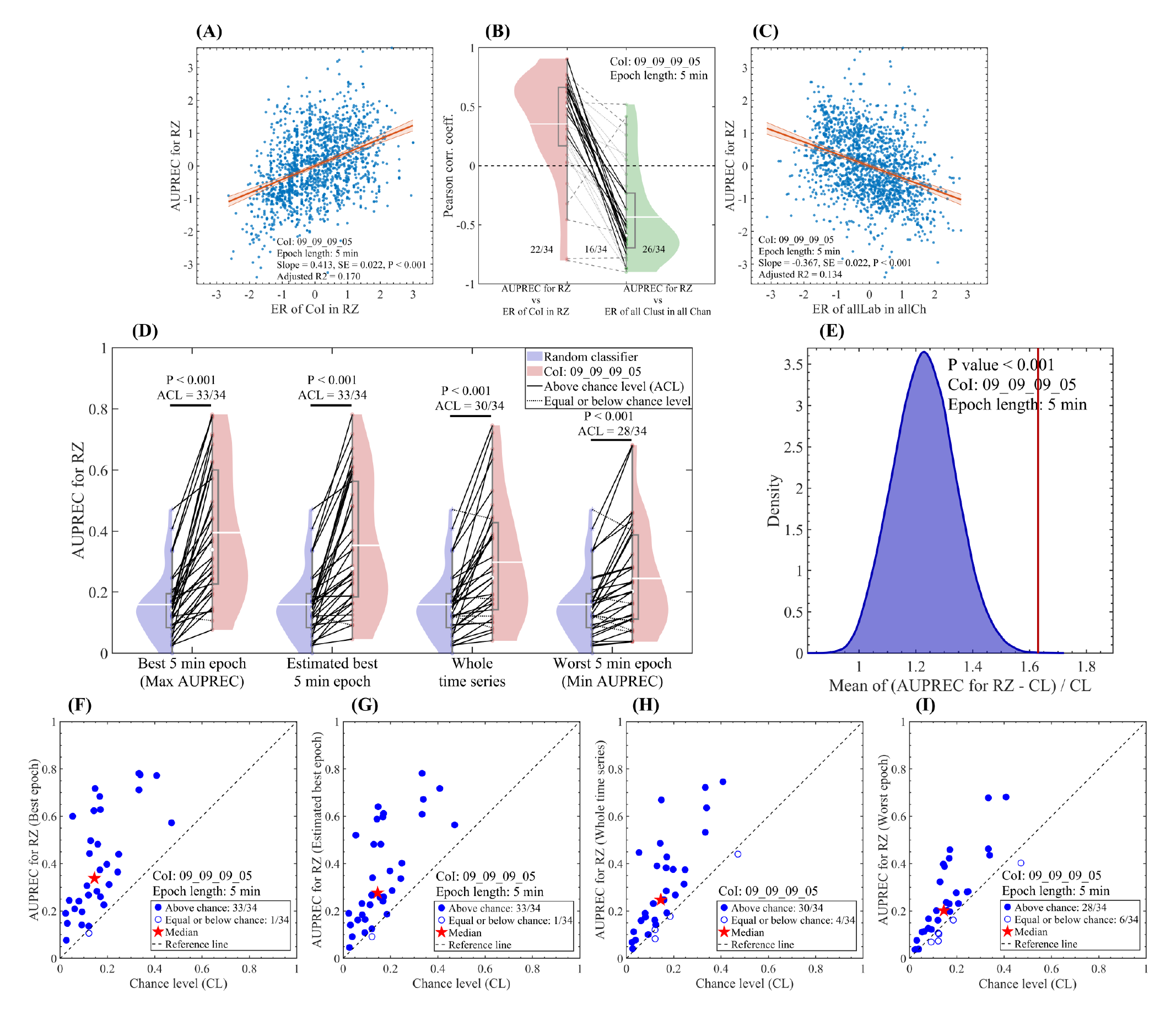
Estimating the best interictal 5 min-epoch for RZ localization. **(A)** Scatter plot showing the correlation between the time series AUPREC for RZ and ER of CoI in RZ. **(C)** Scatter plot showing the correlation between the time series AUPREC for RZ and ER of all Clust in all Chan. In panels **(A)** and **(C)**, all the measures were computed for the CoI: 09_09_09_05 and each dot correspond to the measure value in a particular time position of the sliding epoch of 5 min in length and 90% overlap, covering the whole interictal SEEG time series available in each patient. The red line and red shaded error bars represent the linear regression and the 95% confidence interval, respectively. For panel **(A)** we obtained: Slope = +0.430 [1/min], SE = 0.022 [1/min], P < 0.001, t-statistic of the two-sided hypothesis test. For panel **(C)** we obtained: Slope = −0.367 [1/min], SE = 0.022 [1/min], P < 0.001, t-statistic of the two-sided hypothesis test. **(B)** Violin plots showing all the patients paired across the two correlations, 1) AUPREC for RZ vs ER of CoI in RZ, and 2) AUPREC for RZ vs ER of all Clust in all Chan. Dotted gray lines correspond to patients in which at least one of the two correlations is no significant. Dashed gray lines correspond to patients in which the two correlations are significant and, negative in the red violin plot and positive in the green violin plot, or have the same sign. Solid black lines correspond to patients in which the two correlations are significant and, positive in the red violin plot and negative in the green violin plot. The three fractional numbers accompanying the paired violin plots indicate the fraction of patients presenting significant value for the left-hand, both and right-hand correlations. The statistical significance of the correlations (P < 0.05) was assessed by using the Student’s t distributions of the two-tailed hypothesis test under the null hypothesis that the correlation is zero. **(D)** Violin plots showing all the patients paired across the values of AUPREC for RZ based on a random classifier, computed as the ratio between the number of channels pertaining to RZ and the total number of channels in each patient (blue violin plot), and the AUPREC for RZ based on the rate of events pertaining to the cluster 09_09_09_05 (red violin plot). In an intra-group paired analysis (Wilcoxon signed rank test), the reported P values indicate significant differences between the two distributions of AUPREC values in all the four cases shown. In an inter-group paired analysis, we found significant differences between all the four paired groups (P < 0.001, Wilcoxon signed rank test with the P values Bonferroni-adjusted to correct for multiple comparisons across the 4 cases). **(E)** Histogram showing the distribution of the relative difference of the AUPREC for RZ with respect to the chance level (CL) for a 5 min epoch randomly sampled from the interictal SEEG recordings of each patient (10^5^ random samplings). The CL was computed as the ratio between the number of channels pertaining to EZ and the total number of channels in each patient. The red vertical solid line shown in the histogram indicates the relative difference value corresponding to the estimated best 5 min epoch for near-optimal RZ localization (second case from the left in panel **D**). **(F, G, H, I)** Scatter plots corresponding to the four cases shown in panel **(D)**. In panels D, E and G, the estimated best 5 min epoch for near-optimal EZ localization corresponds to the interictal epoch producing the minimum value of ER of all Clust in all Chan. Symbols and abbreviations: CoI, cluster of interest; RZ, resected zone; ER, events rate; AUPREC, area under the precision and recall curve. ACL, above chance level; ER of CoI in RZ, mean rate of events pertaining to the CoI averaged over the RZ channels; ER of all Clust in all Chan, mean rate of events including all the clusters and averaged across all the channels.

**Figure S13:**
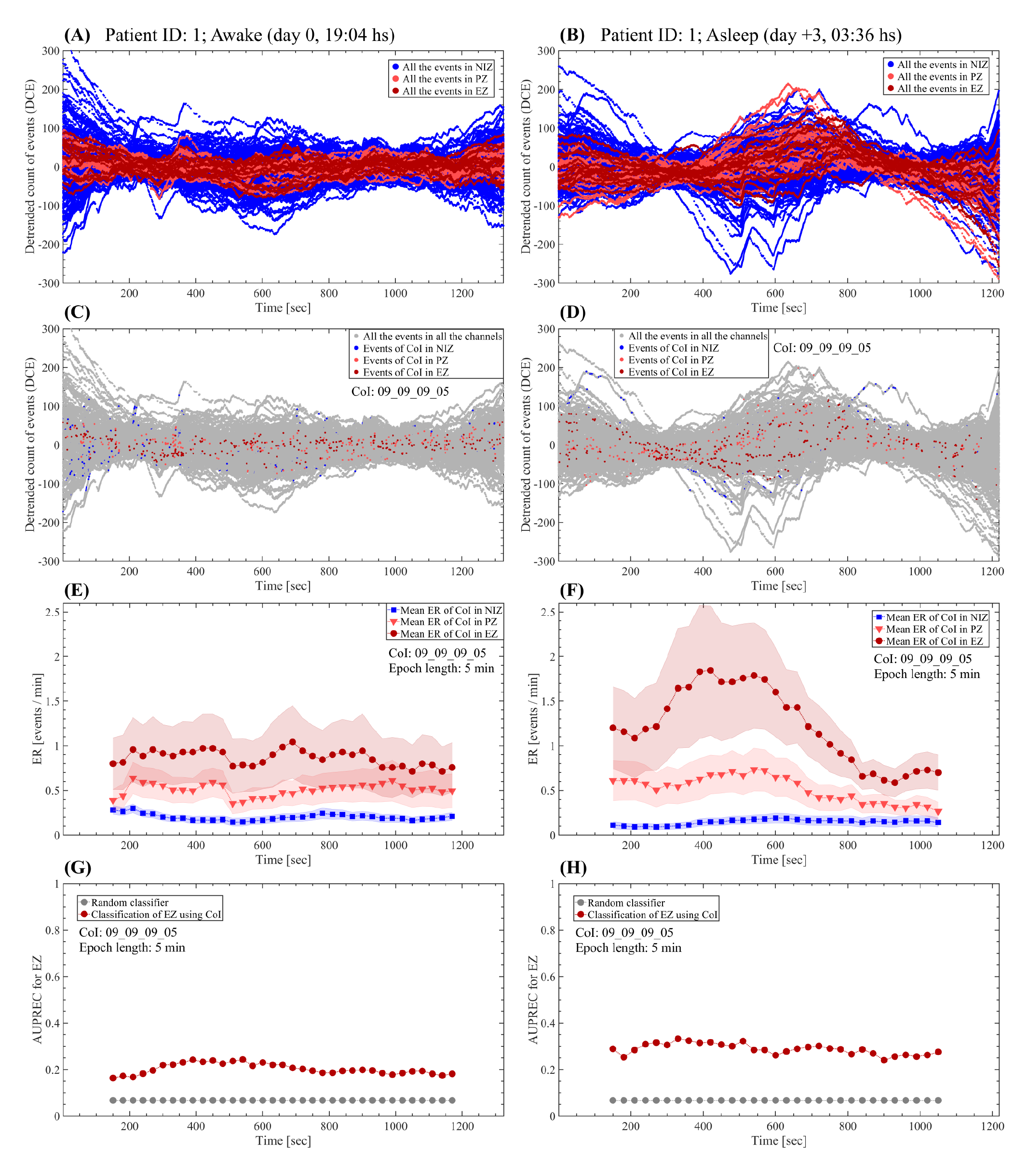
Comparing the fast-ultradian dynamics between awake and non-REM sleep states. **(A, B)** Detrended cumulative count of events (cumulative residual) including all events subtypes (epileptic and non-epileptic) detected by the NODE algorithm (80 clusters). **(C, D)** Cumulative residual showing all the detected events (gay dots) and the discharges pertaining to the cluster 09_09_09_05 in color (NIZ: blue dots, PZ: light red dots, EZ: dark red dots). **(E, F)** Mean rate of events pertaining to the cluster 09_09_09_05. The dots correspond to the mean value of the events rate at each time position of the sliding epoch of 5 min in length and 90% overlap. The shaded error bars correspond to the standard error. **(G, H)** AUPREC for EZ localization based on the rate of events pertaining to the cluster 09_09_09_05 at each time position of the 5 min length sliding epoch. Symbols and abbreviations: CoI, cluster of interest; EZ, epileptogenic zone; PZ, propagation zone; NIZ, non-involved zone; ER, events rate; AUPREC, area under the precision and recall curve.

**Figure S14:**
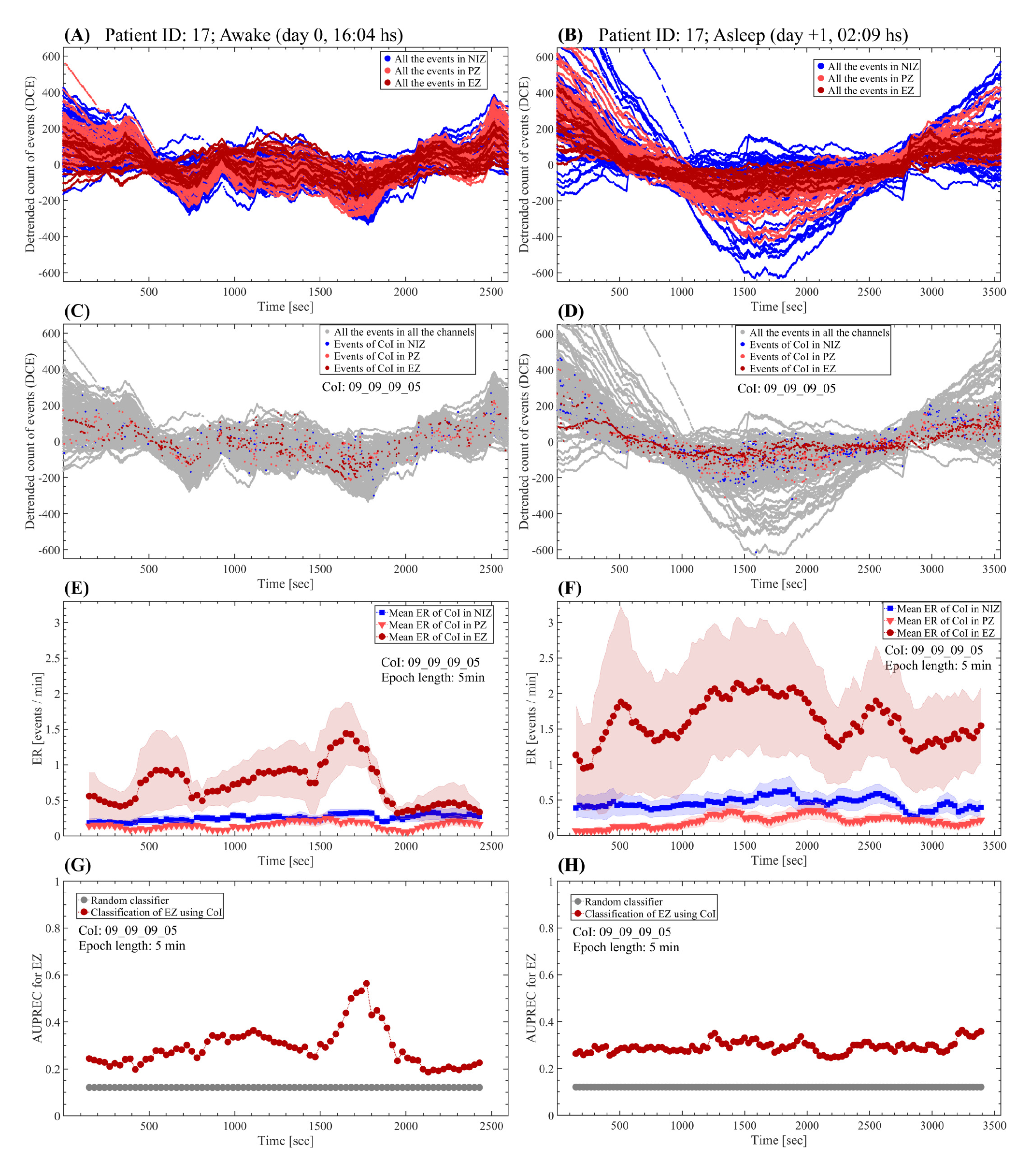
Comparing the fast-ultradian dynamics between awake and non-REM sleep states. **(A, B)** Detrended cumulative count of events (cumulative residual) including all events subtypes (epileptic and non-epileptic) detected by the NODE algorithm (80 clusters). **(C, D)** Cumulative residual showing all the detected events (gay dots) and the discharges pertaining to the cluster 09_09_09_05 in color (NIZ: blue dots, PZ: light red dots, EZ: dark red dots). **(E, F)** Mean rate of events pertaining to the cluster 09_09_09_05. The dots correspond to the mean value of the events rate at each time position of the sliding epoch of 5 min in length and 90% overlap. The shaded error bars correspond to the standard error. **(G, H)** AUPREC for EZ localization based on the rate of events pertaining to the cluster 09_09_09_05 at each time position of the 5 min length sliding epoch. Symbols and abbreviations: CoI, cluster of interest; EZ, epileptogenic zone; PZ, propagation zone; NIZ, non-involved zone; ER, events rate; AUPREC, area under the precision and recall curve.

**Figure S15:**
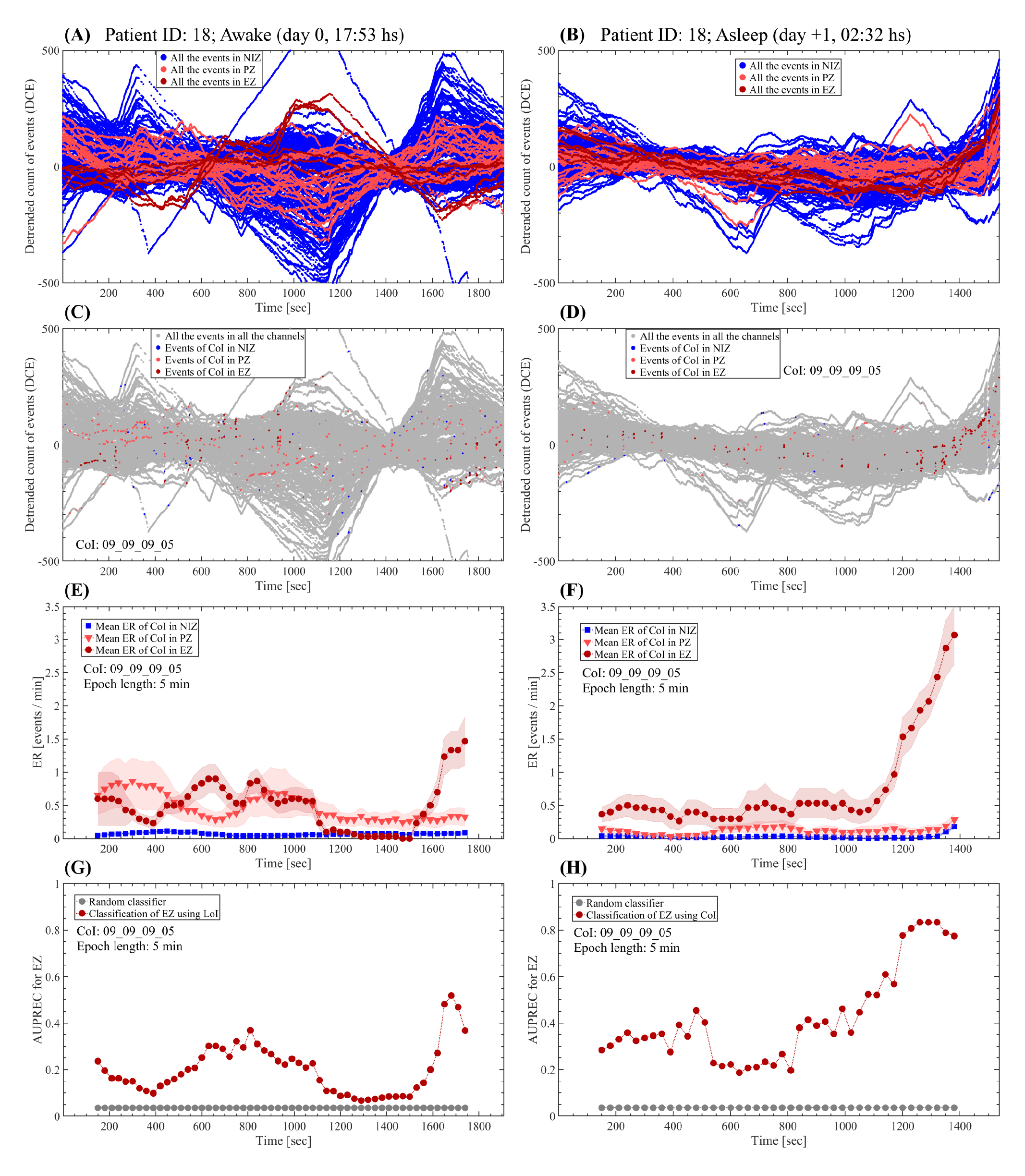
Comparing the fast-ultradian dynamics between awake and non-REM sleep states. **(A, B)** Detrended cumulative count of events (cumulative residual) including all events subtypes (epileptic and non-epileptic) detected by the NODE algorithm (80 clusters). **(C, D)** Cumulative residual showing all the detected events (gay dots) and the discharges pertaining to the cluster 09_09_09_05 in color (NIZ: blue dots, PZ: light red dots, EZ: dark red dots). **(E, F)** Mean rate of events pertaining to the cluster 09_09_09_05. The dots correspond to the mean value of the events rate at each time position of the sliding epoch of 5 min in length and 90% overlap. The shaded error bars correspond to the standard error. **(G, H)** AUPREC for EZ localization based on the rate of events pertaining to the cluster 09_09_09_05 at each time position of the 5 min length sliding epoch. Symbols and abbreviations: CoI, cluster of interest; EZ, epileptogenic zone; PZ, propagation zone; NIZ, non-involved zone; ER, events rate; AUPREC, area under the precision and recall curve.

**Figure S16:**
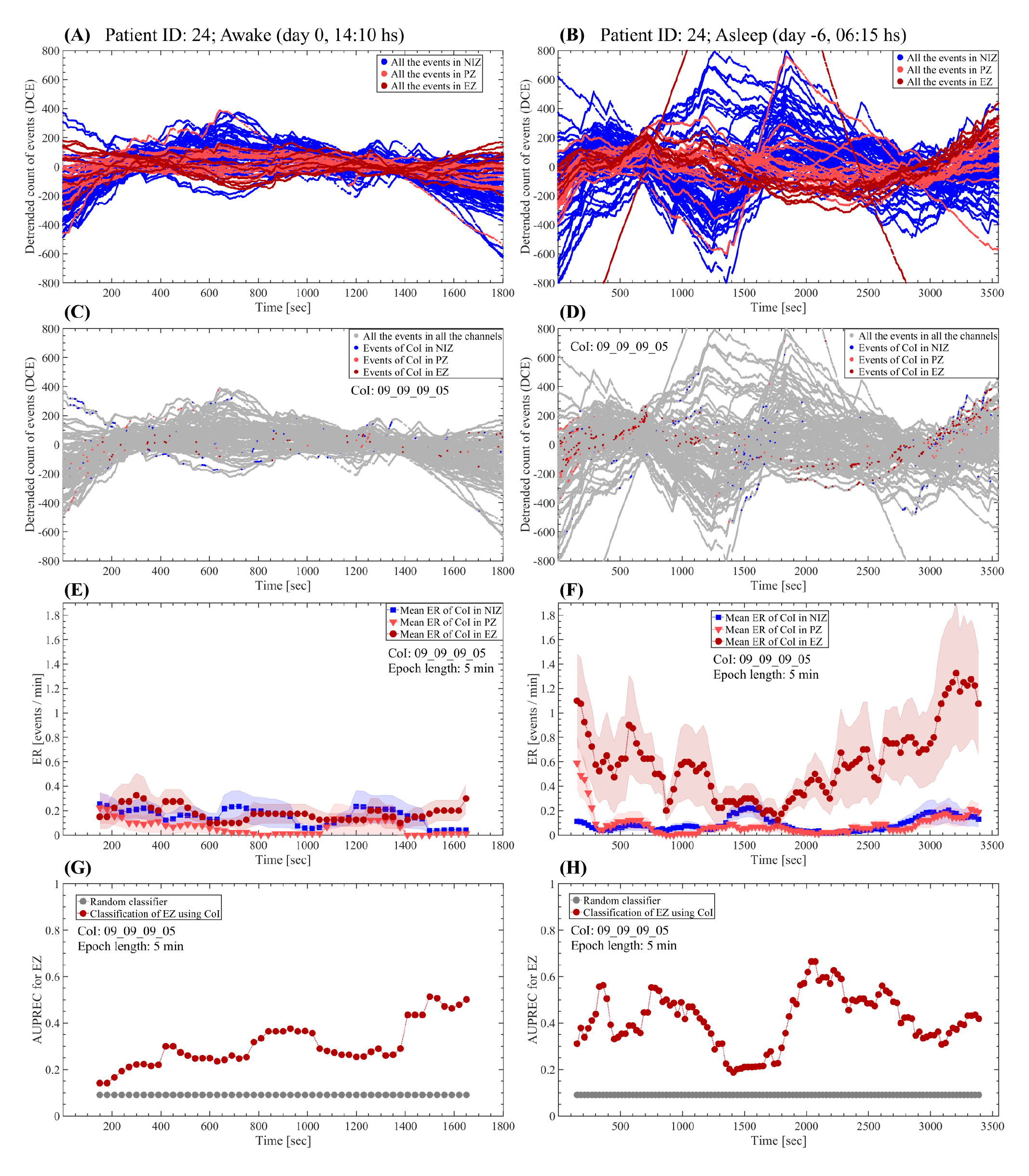
Comparing the fast-ultradian dynamics between awake and non-REM sleep states. **(A, B)** Detrended cumulative count of events (cumulative residual) including all events subtypes (epileptic and non-epileptic) detected by the NODE algorithm (80 clusters). **(C, D)** Cumulative residual showing all the detected events (gay dots) and the discharges pertaining to the cluster 09_09_09_05 in color (NIZ: blue dots, PZ: light red dots, EZ: dark red dots). **(E, F)** Mean rate of events pertaining to the cluster 09_09_09_05. The dots correspond to the mean value of the events rate at each time position of the sliding epoch of 5 min in length and 90% overlap. The shaded error bars correspond to the standard error. **(G, H)** AUPREC for EZ localization based on the rate of events pertaining to the cluster 09_09_09_05 at each time position of the 5 min length sliding epoch. Symbols and abbreviations: CoI, cluster of interest; EZ, epileptogenic zone; PZ, propagation zone; NIZ, non-involved zone; ER, events rate; AUPREC, area under the precision and recall curve.

**Figure S17:**
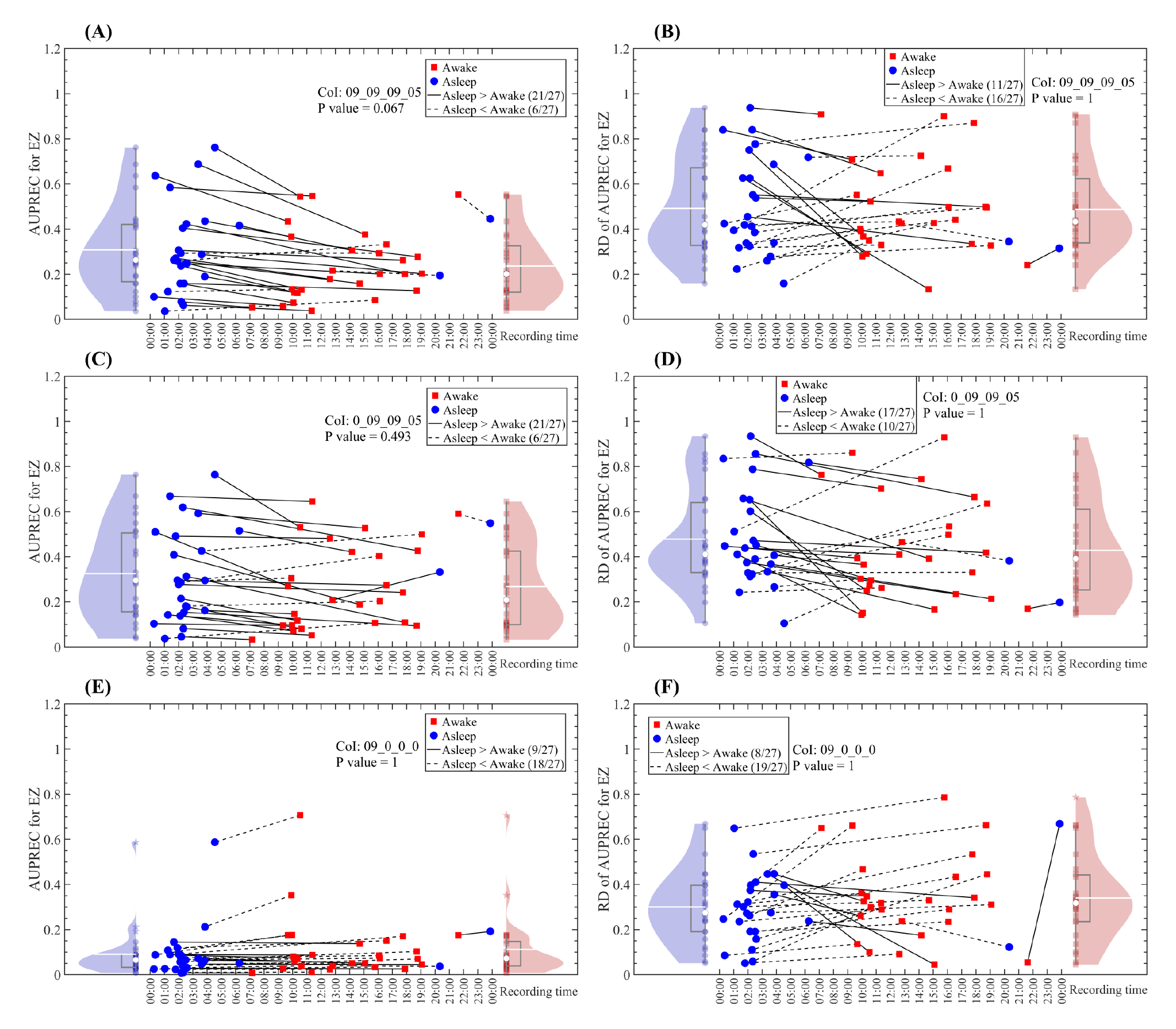
Comparing the fast-ultradian dynamics between awake and non-REM sleep states. **(A, C, E)** AUPREC for EZ values as a function of the time of day (or night) when the SEEG recordings were made. The values shown correspond to the mean value of the AUPREC for EZ time series computed across all the 5 min length epochs covering the whole SEEG time series (bipolar derivations) available in each patient (see Table S1). **(B, D, F)** RD of AUPREC for EZ values as a function of the time of day (or night) when the SEEG recordings were made. The relative difference (RD) was computed between the extreme values ((max - min) / max) of the AUPREC for EZ time series of each patient. The panels (A, B), (C, D) and (E, F) show the mean and RD of AUPREC for EZ values computed based on the rate of interictal events pertaining to the clusters 09_09_09_05 (epileptiform spikes), 0_09_09_05 (spike-wave complexes) and 09_0_0_0 (non-epileptiform events), respectively. Blue and red dots (and violin plots) correspond to the awake at rest and non-REM sleep patients states, respectively. All the reported P values correspond to a paired analysis based on a Wilcoxon signed rank test with the P values Bonferroni-adjusted to correct for multiple comparisons across the 80 NODE clusters.

**Figure S18:**
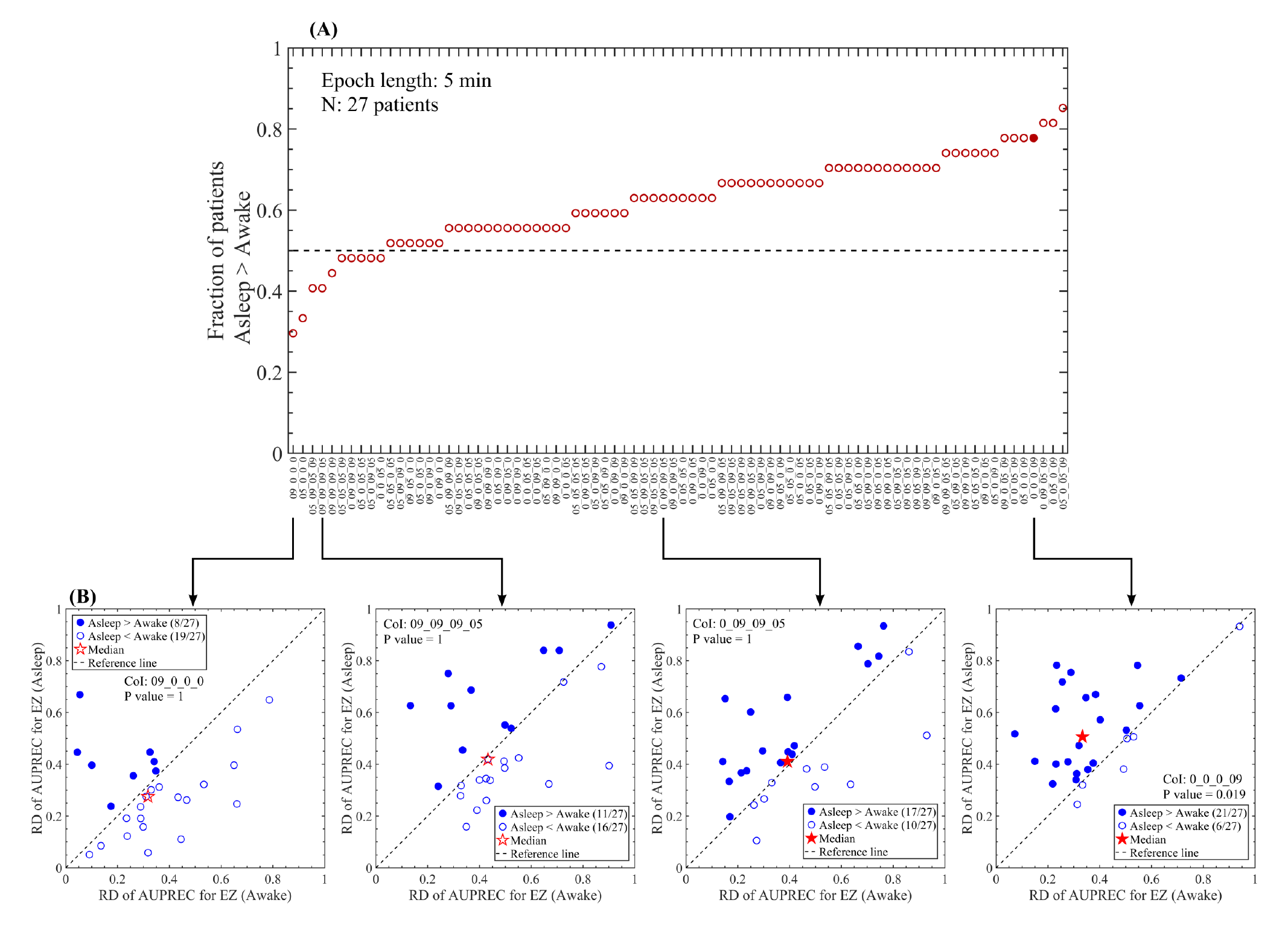
Comparing the fast-ultradian dynamics between awake and non-REM sleep states. **(A)** Fraction of patients disclosing RD of AUPREC for EZ values in the non-REM sleep state greater than those in the awake at rest state, as a function of the NODE clusters. The relative difference (RD) was computed between the extreme values ((max - min) / max) of the AUPREC for EZ time series of each patient. Filled and empty circles indicate clusters producing significant (Bonferroni-adjusted P value < 0.05) and non-significant (Bonferroni-adjusted P value ≥ 0.05) difference between the awake at rest and non-REM sleep states in terms of the RD of AUPREC for EZ (Wilcoxon signed rank test with the P values Bonferroni-adjusted to correct for multiple comparisons across the 80 NODE clusters). **(B)** Scatter plots corresponding to four NODE clusters shown in panel A. All the reported P values correspond to a paired analysis based on a Wilcoxon signed rank test with the P values Bonferroni-adjusted to correct for multiple comparisons across the 80 NODE clusters.

**Figure S19:**
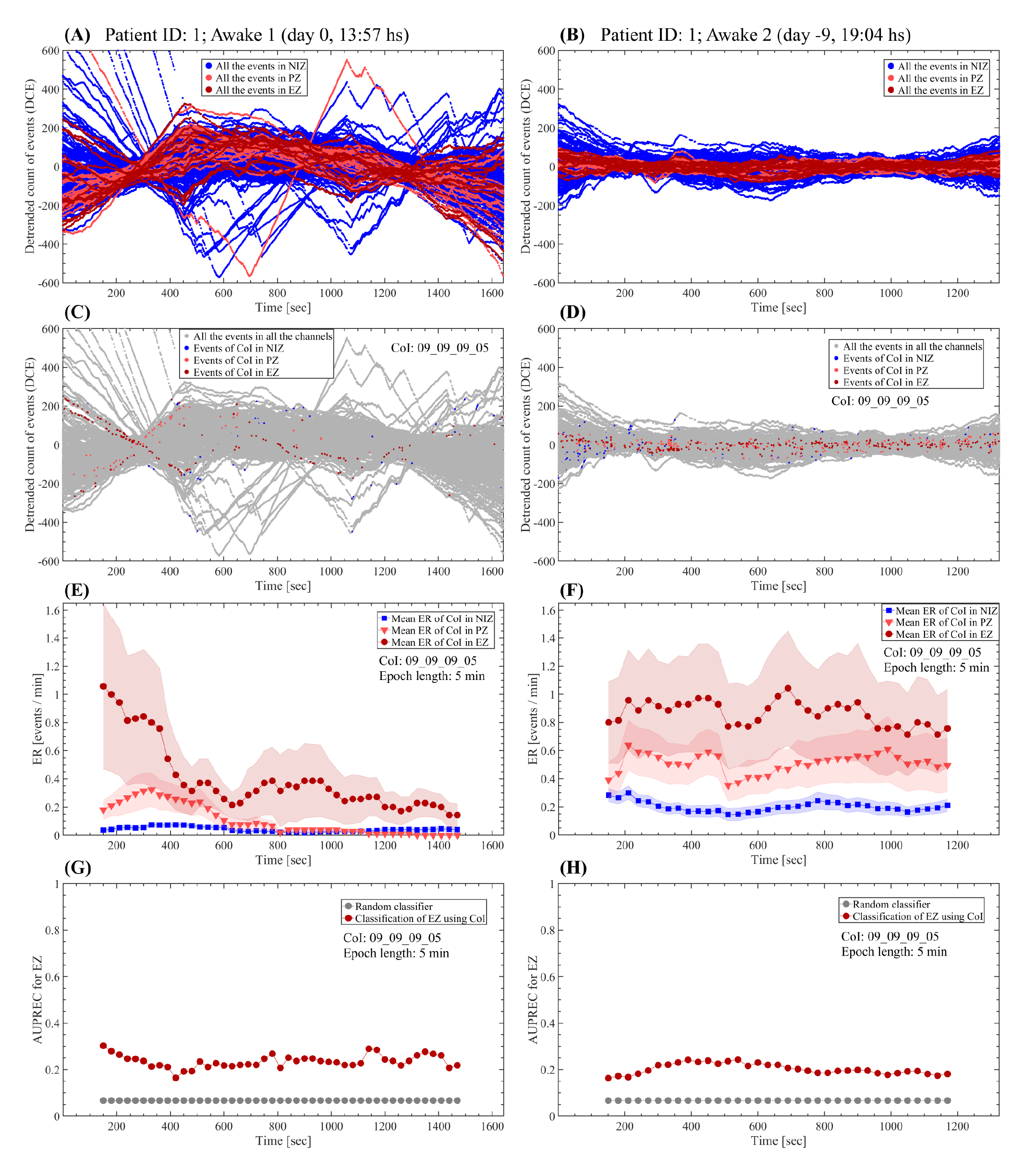
Comparing the fast-ultradian dynamics between two SEEG recording sessions in awake state. **(A, B)** Detrended cumulative count of events (cumulative residual) including all events subtypes (epileptic and non-epileptic) detected by the NODE algorithm (80 clusters). **(C, D)** Cumulative residual showing all the detected events (gay dots) and the discharges pertaining to the cluster 09_09_09_05 in color (NIZ: blue dots, PZ: light red dots, EZ: dark red dots). **(E, F)** Mean rate of events pertaining to the cluster 09_09_09_05. The dots correspond to the mean value of the events rate at each time position of the sliding epoch of 5 min in length and 90% overlap. The shaded error bars correspond to the standard error. **(G, H)** AUPREC for EZ localization based on the rate of events pertaining to the cluster 09_09_09_05 at each time position of the 5 min length sliding epoch. Symbols and abbreviations: CoI, cluster of interest; EZ, epileptogenic zone; PZ, propagation zone; NIZ, non-involved zone; ER, events rate; AUPREC, area under the precision and recall curve.

**Figure S20:**
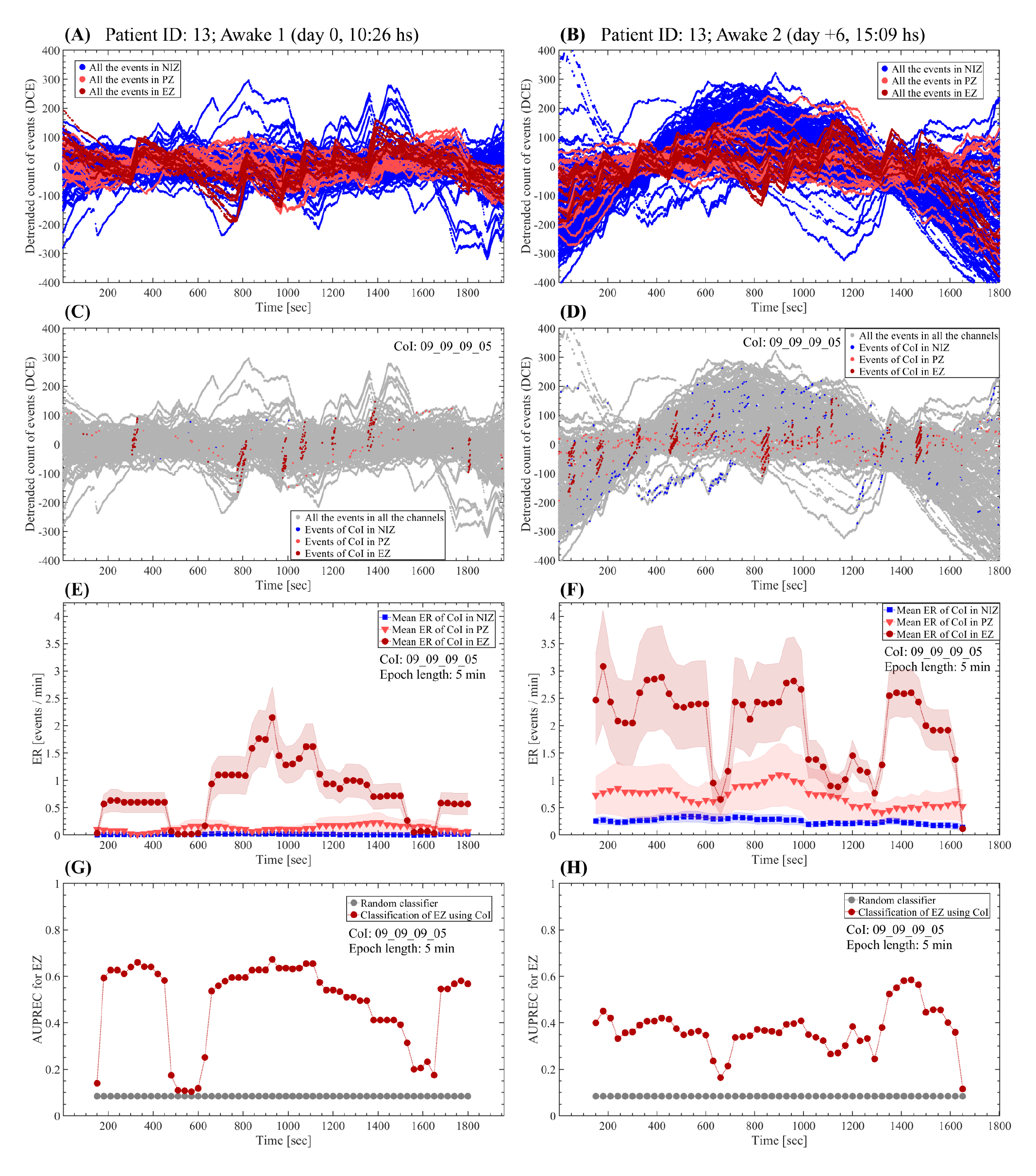
Comparing the fast-ultradian dynamics between two SEEG recording sessions in awake state. **(A, B)** Detrended cumulative count of events (cumulative residual) including all events subtypes (epileptic and non-epileptic) detected by the NODE algorithm (80 clusters). **(C, D)** Cumulative residual showing all the detected events (gay dots) and the discharges pertaining to the cluster 09_09_09_05 in color (NIZ: blue dots, PZ: light red dots, EZ: dark red dots). **(E, F)** Mean rate of events pertaining to the cluster 09_09_09_05. The dots correspond to the mean value of the events rate at each time position of the sliding epoch of 5 min in length and 90% overlap. The shaded error bars correspond to the standard error. **(G, H)** AUPREC for EZ localization based on the rate of events pertaining to the cluster 09_09_09_05 at each time position of the 5 min length sliding epoch. Symbols and abbreviations: CoI, cluster of interest; EZ, epileptogenic zone; PZ, propagation zone; NIZ, non-involved zone; ER, events rate; AUPREC, area under the precision and recall curve.

**Figure S21:**
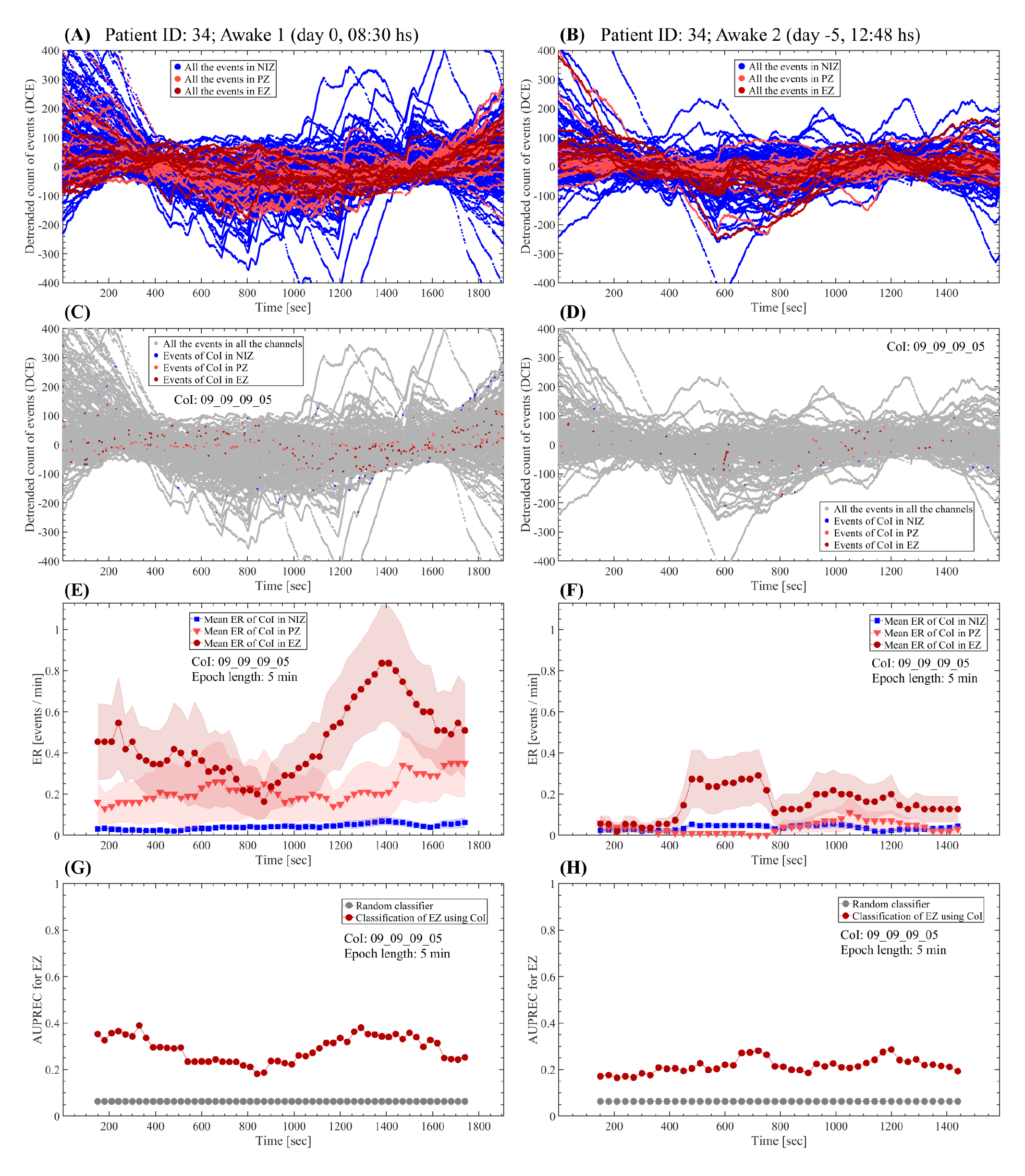
Comparing the fast-ultradian dynamics between two SEEG recording sessions in awake state. **(A, B)** Detrended cumulative count of events (cumulative residual) including all events subtypes (epileptic and non-epileptic) detected by the NODE algorithm (80 clusters). **(C, D)** Cumulative residual showing all the detected events (gay dots) and the discharges pertaining to the cluster 09_09_09_05 in color (NIZ: blue dots, PZ: light red dots, EZ: dark red dots). **(E, F)** Mean rate of events pertaining to the cluster 09_09_09_05. The dots correspond to the mean value of the events rate at each time position of the sliding epoch of 5 min in length and 90% overlap. The shaded error bars correspond to the standard error. **(G, H)** AUPREC for EZ localization based on the rate of events pertaining to the cluster 09_09_09_05 at each time position of the 5 min length sliding epoch. Symbols and abbreviations: CoI, cluster of interest; EZ, epileptogenic zone; PZ, propagation zone; NIZ, non-involved zone; ER, events rate; AUPREC, area under the precision and recall curve.

**Figure S22:**
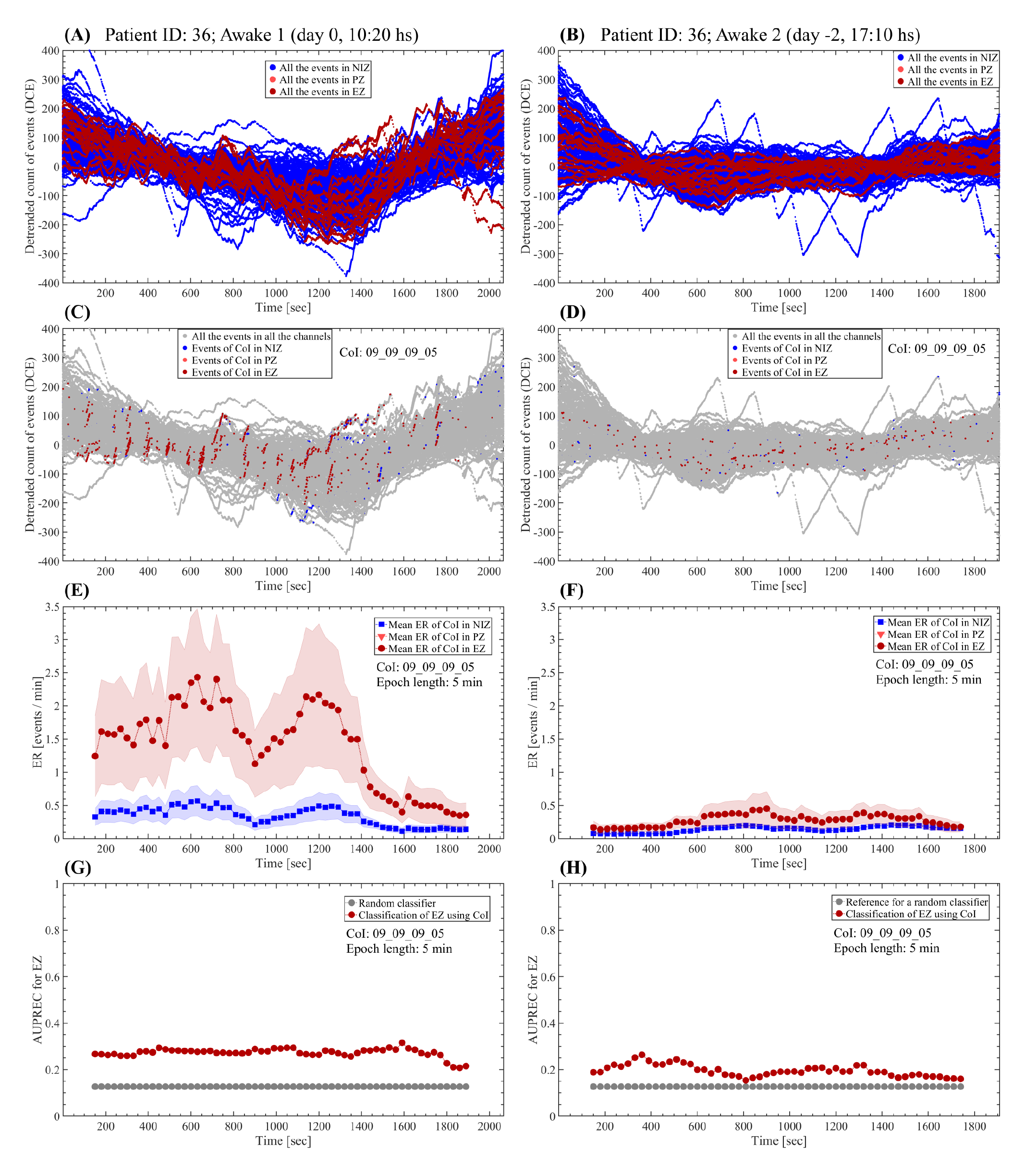
Comparing the fast-ultradian dynamics between two SEEG recording sessions in awake state. **(A, B)** Detrended cumulative count of events (cumulative residual) including all events subtypes (epileptic and non-epileptic) detected by the NODE algorithm (80 clusters). **(C, D)** Cumulative residual showing all the detected events (gay dots) and the discharges pertaining to the cluster 09_09_09_05 in color (NIZ: blue dots, PZ: light red dots, EZ: dark red dots). **(E, F)** Mean rate of events pertaining to the cluster 09_09_09_05. The dots correspond to the mean value of the events rate at each time position of the sliding epoch of 5 min in length and 90% overlap. The shaded error bars correspond to the standard error. **(G, H)** AUPREC for EZ localization based on the rate of events pertaining to the cluster 09_09_09_05 at each time position of the 5 min length sliding epoch. Symbols and abbreviations: CoI, cluster of interest; EZ, epileptogenic zone; PZ, propagation zone; NIZ, non-involved zone; ER, events rate; AUPREC, area under the precision and recall curve.

**Figure S23:**
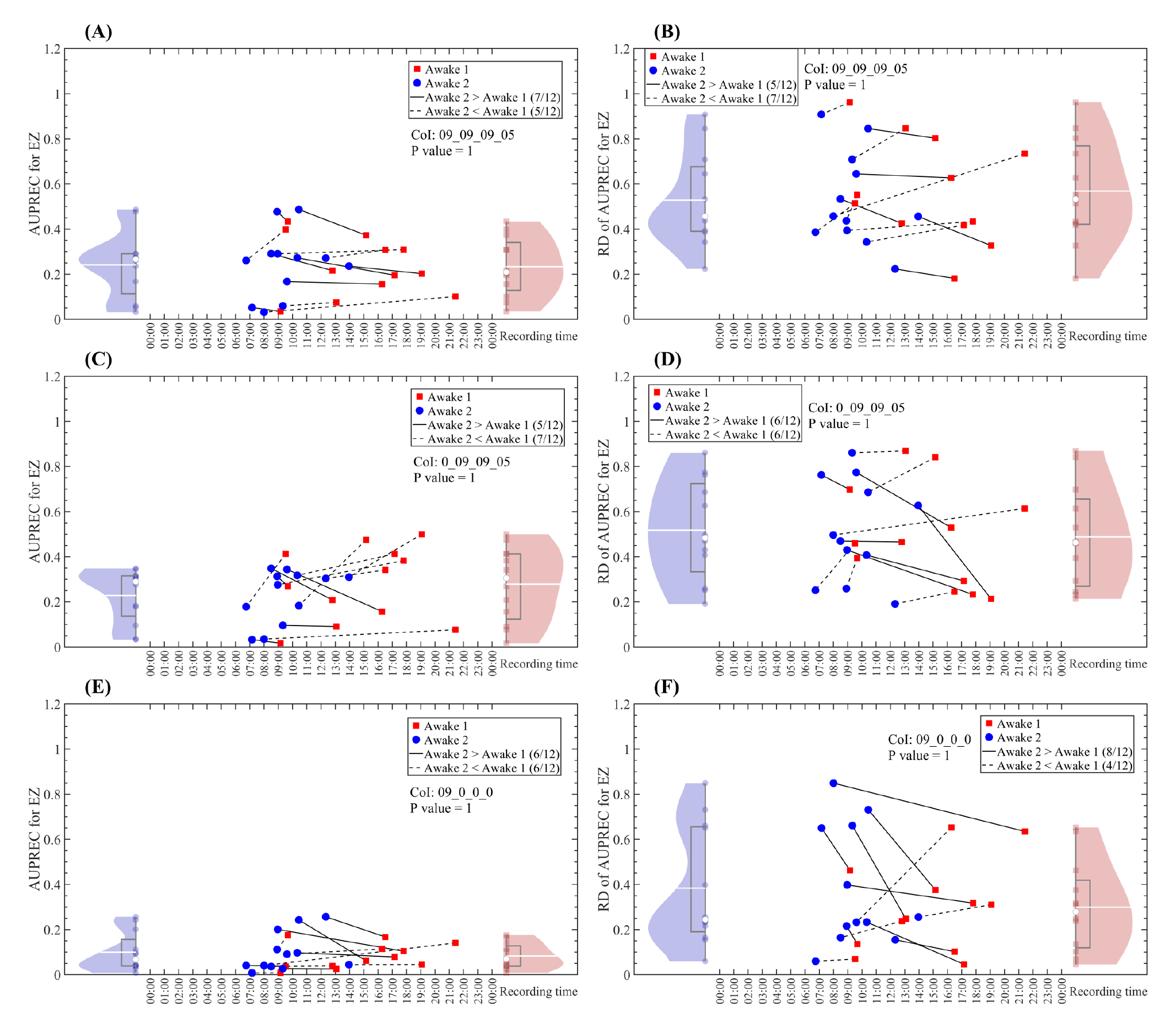
Comparing the fast-ultradian dynamics between two SEEG recording sessions in awake state. **(A, C, E)** AUPREC for EZ values as a function of the time of day when the SEEG recordings were made. The values shown correspond to the mean value of the AUPREC for EZ time series computed across all the 5 min length epochs covering the whole SEEG time series (bipolar derivations) available in each patient (see Table S1). **(B, D, F)** RD of AUPREC for EZ values as a function of the time of day (or night) when the SEEG recordings were made. The relative difference (RD) was computed between the extreme values ((max - min) / max) of the AUPREC for EZ time series of each patient. The panels (A, B), (C, D) and (E, F) show the mean and RD of AUPREC for EZ values computed based on the rate of interictal events pertaining to the clusters 09_09_09_05 (epileptiform spikes), 0_09_09_05 (spike-wave complexes) and 09_0_0_0 (non-epileptiform events), respectively. Blue and red dots (and violin plots) correspond to two SEEG recording sessions taken at different days and time of day for the same patient state (awake at rest, see Tables S2 and S3). All the reported P values correspond to a paired analysis based on a Wilcoxon signed rank test with the P values Bonferroni-adjusted to correct for multiple comparisons across the 80 NODE clusters.

**Figure S24:**
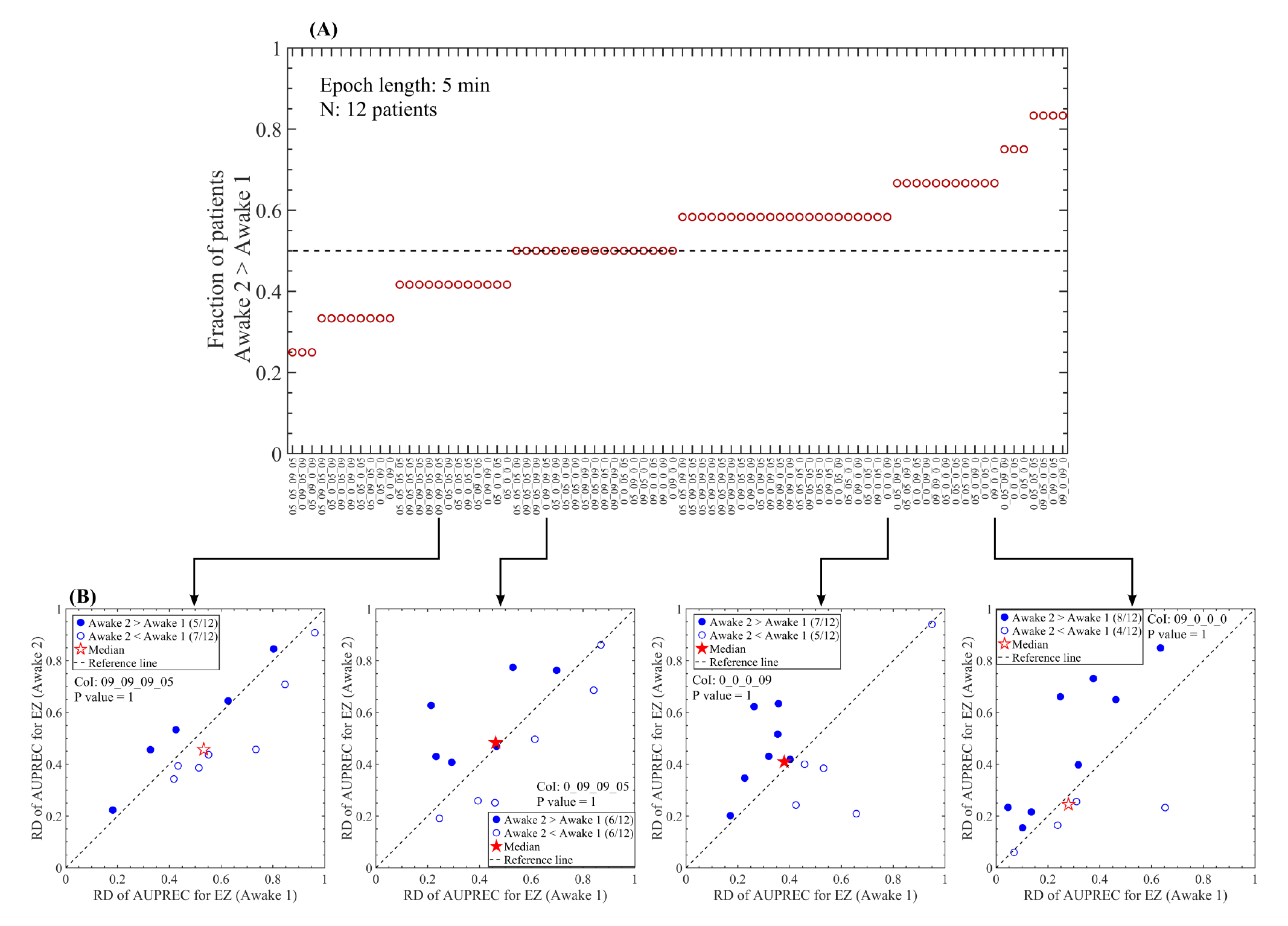
Comparing the fast-ultradian dynamics between two SEEG recording sessions in awake state. **(A)** Fraction of patients disclosing RD of AUPREC for EZ values in the Awake 2 state greater than those in the Awake 1 state, as a function of the NODE clusters. The relative difference (RD) was computed between the extreme values ((max - min) / max) of the AUPREC for EZ time series of each patient. Filled and empty circles indicate clusters producing significant (Bonferroni-adjusted P value < 0.05) and non-significant (Bonferroni-adjusted P value ≥ 0.05) difference between the Awake 1 and Awake 2 states in terms of the RD of AUPREC for EZ (Wilcoxon signed rank test with the P values Bonferroni-adjusted to correct for multiple comparisons across the 80 NODE clusters). **(B)** Scatter plots corresponding to four NODE clusters shown in panel A. All the reported P values correspond to a paired analysis based on a Wilcoxon signed rank test with the P values Bonferroni-adjusted to correct for multiple comparisons across the 80 NODE clusters.

